# Genome-wide Cross-multi-trait Analysis into Shared Genetic Architecture of Coronary Artery Disease and Aortic Aneurysms

**DOI:** 10.1101/2025.06.27.25330395

**Authors:** Jimi Virkkala, FINNGEN, Johannes Kettunen, Minna U. Kaikkonen

## Abstract

While affecting distinct vascular territories, coronary artery disease, abdominal aortic aneurysms and thoracic aortic aneurysms share similar risk factors and pathophysiological resemblance. To identify novel pleiotropic and heterogenous loci, we performed a joint meta-analysis leveraging the shared genetic architecture of thereof. Using summary statistics from three independent genome-wide association studies, we perform nine separate multivariate genome-wide scans together with cross-trait validation and identify 59 regions previously unreported as genome-wide significant for their respective traits. In our downstream investigations, we comprehensively map the functional characteristics of the loci, revealing common patterns of enriched cell types and tissues, prominent cellular pathways, expression quantitative loci and genetic correlation. Findings from this study support the presence of cardiovascular polygenicity, identify novel genome-wide significant variants for coronary artery disease and aortic aneurysms, and extend the previous knowledge on their genetic overlap and relationship. Taken together, these results show that joint analysis of related atherosclerotic disease traits allows for identification of unified biology that may offer the opportunity for future therapeutic manipulation.

## INTRODUCTION

Coronary artery disease (CAD), abdominal aortic aneurysm (AAA), and thoracic aortic aneurysm (TAA) remain major contributors to cardiovascular morbidity and mortality worldwide (Roth *et al*, 2020). While clinically distinct, cardiovascular diseases (CVDs) have high heritability (McPherson & Tybjaerg-Hansen, 2016; Wahlgren *et al*, 2010; Ostberg *et al*, 2020), can co-occur (Toghill *et al*, 2017) and have widely shared known risk factors (Vaduganathan *et al*, 2022), all indicative of common genetic determinants. Indeed, extensive efforts in genome-wide association studies (GWAS) have already identified numerous risk loci for CAD (Koyama *et al*, 2020; Tcheandjieu *et al*, 2022; Aragam *et al*, 2022), AAA (Zheng *et al*, 2024; Klarin *et al*, 2020) and TAA (Klarin *et al*, 2023), highlighting overlapping pathophysiological mechanisms related to apoptosis and phenotypic modulation of vascular smooth muscle cells (VSMCs) (Petsophonsakul *et al*, 2019; Yu *et al*, 2024), extracellular matrix (ECM) degradation and remodelling (Lin & Davis, 2023), chronic inflammation (Henein *et al*, 2022), and the loss of lipid metabolism homeostasis (Bhargava *et al*, 2022; Wazir *et al*, 2023), among others (Jebari-Benslaiman *et al*, 2022; Cho *et al*, 2023). Nevertheless, the current interpretation of possible genetic pleiotropy within the cardiovascular disease setting remains incomplete.

Multivariate GWAS have emerged as powerful extensions of their traditional univariate counterparts for modelling joint genetic effects in related complex traits (Porter & O’Reilly, 2017; Galesloot *et al*, 2014; Turchin & Stephens, 2019) and improving discovery of variants with modest effects from underpowered studies (23andMe Research Team *et al*, 2018). To this end, multi-trait methodologies typically leverage the inherent genetic correlation architecture over a range of phenotypes with linear mixed models, Bayesian statistics, or meta-analytic approaches that reduce the burden for multiple testing while accounting for population structures and cryptic relatedness (Lee *et al*, 2012; 23andMe Research Team *et al*, 2018; Ray & Boehnke, 2018; Li & Zhu, 2017; Fernandes *et al*, 2022; Qi *et al*, 2024; Grotzinger *et al*, 2019). The benefits of multivariate analyses in providing deeper insights into shared and disease-specific genetics have been extensively demonstrated across a broad spectrum of clinical disorders, both within and without the CVD milieu (Wu *et al*, 2020; Rosoff *et al*, 2023; Han *et al*, 2023; Zhu *et al*, 2024; Alemany *et al*, 2023).

Herein, we conducted multivariate GWAS meta-analyses of CAD, AAA and TAA across three non-overlapping cohorts to enhance the discovery of novel genetic risk factors of thereof. As a complementary sensitivity analysis, we further applied an independent cross-trait GWAS approach to validate the potential pleiotropy of our multi-trait associations. Finally, via series of comprehensive postGWAS (pGWAS) analyses, we systematically annotate our findings and inform our understanding on the shared and distinct pathways, cell types and tissues involved in CVD biology.

## METHODS

### Study populations

#### FinnGen

Launched in 2017, FinnGen is a public-private research project by Finnish biobanks, their background organisations and international pharmaceutical industry partners, aiming to better the understanding of disease mechanisms by combining genomic and healthcare data from up to 500,000 Finns. All FinnGen partners are listed at: https://www.finngen.fi/en/partners. In FinnGen, genotyping was carried out with ThermoFisher, Illumina and Affymetrix arrays. Following sample quality-control, genotypes were imputed using Finnish population-specific SisuV4 reference panel and variants with imputation INFO scores of <0.6 or MAF values of <0.0001 were excluded. Detailed genotyping, imputation and quality-control information are described in the original flagship publication (Kurki *et al*, 2023).

#### UK Biobank

The UK Biobank (UKBB) is a large-scale population-based cohort study with deep phenotypic and genetic data from half a million participants aged 40–69 yr from the United Kingdom since 2006. The resource is aimed at approved researchers from all types of academic, charity, government and commercial organisations to conduct studies improving public health. Briefly for genetic data, genotyping was performed on all participants using the UK Biobank Axiom Array. Approximately 850,000 markers were directly measured, with >90million variants imputed using the Haplotype Reference Consortium and UK10K + 1000 Genomes reference panels. The genotype platforms, genetic quality-controls and imputation procedures have been detailed elsewhere (Bycroft *et al*, 2018).

#### Million Veteran Program

The Million Veteran Program (MVP) is a national research initiative by the U.S. Department of Veterans Affairs (VA) aimed at understanding how genes, lifestyle, and military experiences affect health and disease. From its launch in 2011, the MVP cohort now consists of over one million veteran participants, making it one of the world’s largest genomic and health databases. By analysing genetic and medical data, researchers seek to advance precision medicine, improve treatments, and enhance healthcare for veterans and the general population (Gaziano *et al*, 2016). Genotyping in the cohort was performed using MVP 1.0 custom Axiom array, imputed to hybrid 1000 Genomes/African Genome Resource reference panel and TOPMed reference panel for approximately 650,000 individuals. Genotyping array design, imputation and quality-control have been previously described in detail (Hunter-Zinck *et al*, 2020).

### Study data

We obtained GWAS summary statistics for CAD, AAA and TAA from FinnGen, the Pan-UKBB project (preprint: Karczewski *et al*, 2024), and MVP. Specifically for Pan-UKBB and MVP, we used data from the European (EUR) ancestry subsets. In FinnGen phenotypes were characterised based on the 10th revision of the International Classification of Diseases (ICD-10) coding system and subsequent GWASs were performed with the Regenie software (Mbatchou *et al*, 2021), using an additive genetic model adjusting each phenotype for age, sex, the first 10 genetic principal components, and genotyping arrays. The Pan-UKBB project utilised a two-step GWAS approach using SAIGE (Zhou *et al*, 2018) and inverse-variance weighted meta-analyses to generate their within-ancestry, phecode-based phenotypes (preprint: Karczewski *et al*, 2024). Methodologies and phenotype definitions used in the MVP studies are detailed in the original publications (Tcheandjieu *et al*, 2022; Klarin *et al*, 2020, 2023).

The FinnGen and UKBB data were derived from the two-way meta-analysis of FinnGen Data Freeze 12 (DF12) and the Pan-UKBB study by the FinnGen analysis team to ensure uniform endpoint definitions. The custom workflow incorporates only endpoints with perfect ICD-10 to phecode overlap and redefines Pan-UKBB endpoints to match corresponding FinnGen endpoints where necessary.

Summary statistics from MVP studies of CAD, AAA and TAA were downloaded from the database of Genotypes and Phenotypes (dbGaP) analysis accessions pha005193.1, pha005146.1, pha005255.1, respectively, included under study accession number phs001672.v11.p1.

MVP summary statistics were converted from GRCh37 to GRCh38 genome build prior to analyses using the rtracklayer version 1.58.0 (Lawrence *et al*, 2009). Moreover, we harmonised summary statistics between cohorts with MungeSumstats version 1.10.1 (Murphy *et al*, 2021) with minimal variant filtering applied. Specifically, checked for SNV mapping to reference genome, allele flipping, missing or duplicated entries and unrealistic *P*-values. When not present, SNV effect estimate betas (BETA) were imputed from odds ratio (OR) as log(OR), and Z-scores from betas and associated standard errors (SE) as BETA/SE. For each summary statistic, we calculated the effective sample size (N_Eff_) with the LDSC method:

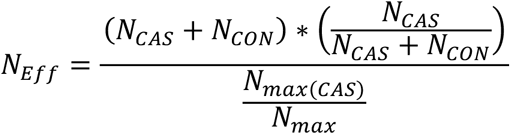

N_CAS_ is the number of GWAS cases, N_CON_ is the number of GWAS controls.

### Multi-trait meta-analyses of GWAS

We used MTAG beta version 0.9.0 (23andMe Research Team *et al*, 2018) as our primary approach to identify potentially pleiotropic loci shared between CAD, AAA and TAA. MTAG applies generalised inverse-variance-weighted (IVW) meta-analysis of multiple traits, accommodating for potential sample overlap of different GWAS experiments and leverages genetic correlation between analysis traits to enhance statistical power. Notably, MTAG assumes SNVs to share the same variance-covariance matrix of effect estimates across analysis traits (23andMe Research Team *et al*, 2018).

Here, we adopted a two-stage approach to perform meta-analyses and multi-trait analyses of GWAS summary statistics using MTAG.

First, harmonised CAD, AAA and TAA summary statistics from FinnGen, UKBB and MVP were separately meta-analysed via inclusion of MTAG parameters –*perfect_gencov* and –*equal_h2* to build trait-specific summary statistics. In total, our meta-analyses for (1) CAD included 202,505 cases and 983,067 controls, for (2) AAA 10,747 cases and 1,011,639 controls, and (3) TAA 13,058 cases and 1,191,932 controls. For each meta-analysis, MTAG computes a GWAS equivalent sample size that approximates the increase in power compared to a univariate GWAS. To avoid overestimation in the subsequent multi-trait analyses, we re-estimated trait-specific N_Eff_ by weighting the MTAG output N with corresponding prior meta-analysis N_Eff_, as per original author recommendation:

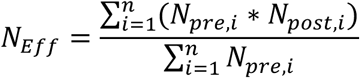

Where,

N_pre_ is the trait-specific effective sample size prior to MTAG analysis, N_post_ is the trait-specific GWAS equivalent sample size estimated in MTAG output.

In the second analysis stage, the trait-specific, meta-analysed outputs from the first stage were further included in a three-way MTAG analysis of CAD, AAA and TAA, as well as pair-wise multi-trait analyses of CAD with AAA, CAD with TAA, and AAA with TAA. SNVs with MAF<0.01 were discarded from the final output. We also enabled MTAG maximum FDR (maxFDR) calculation to observe the presence of potentially confounding inflation of trait-specific effect estimates. The final multi-trait N_MTAG_ in each trait-configuration was weighted with prior meta-analysis N_Eff_ as described above, for use in in applicable downstream pGWAS analyses.

### Cross-trait GWAS validation of novel loci

To control for violation of assumptions set by MTAG, we performed a further sensitivity analysis using cross-phenotype association analysis method CPASSOC version 1.01 (Li & Zhu, 2017) across our four multi-trait configurations. CPASSOC incorporates GWAS summary statistics from multiple traits to detect pleiotropic variants accounting for underlying population structure and cryptic relatedness (Li & Zhu, 2017). It implements two statistical tests S_hom_ and S_het_ to test variant effects either from homogenous or heterogenous studies. As S_hom_ represents the maximum weighted sum of trait-specific genetic effects under fixed-effects assumption, it generally lacks the necessary detection power under the presence of between-study heterogeneity, intrinsic to a multi-trait setting. We therefore chose to apply S_het_ for our analysis to better capture the between-trait variance from different phenotypes and GWAS study settings (Li & Zhu, 2017). As such, we ultimately defined significant pleiotropic variants as SNVs with *P*_MTAG_<5×10^-8^ and *P*_CPASSOC_<5×10^-8^.

### Definition of independent and novel loci

We defined a locus as a window of 2 MB (± 1 000 000 bases) containing at least one variant associated with a trait at *P*<5×10^-8^. Independent loci not within 2 MB (± 1 000 000 bases) of previously reported lead variants were considered novel. All genomic positions are reported using coordinates from the GRCh38 build of the human genome.

### Functional annotation of association signals

#### Candidate gene prioritisation

We employed the Open Targets Genetics variant-to-gene (V2G) pipeline to assign potential target genes to our previously unreported lead variants. Briefly, for each variant the V2G pipeline considers and integrates evidence from molecular QTL and chromatin interaction experiments, functional variant effect predictions (Ensembl VEP) and genomic distance from variant to canonical TSSs. Outputs are harmonised, filtered and weighted based on data source and aggregated into final scores for each candidate gene (Buniello *et al*, 2025; Ghoussaini *et al*, 2021; Mountjoy *et al*, 2021). For each of our lead variants, we extracted the top five predictions for each lead variant for assigning a candidate gene to a particular locus. Moreover, we applied ANNOVAR (Wang *et al*, 2010) as implemented in FUMA version 1.3.3d (Watanabe *et al*, 2017) and the ANNO tool developed by the FinnGen analysis team. Finally, we determined the potential candidate gene(s) with relevant biological function with help of extensive manual review of literature and Genbank (Clark *et al*, 2016), Uniprot (The UniProt Consortium *et al*, 2025) and GTEx (GTEx Consortium *et al*, 2017)databases.

#### Gene-based and pathway analyses

We applied Multi-marker Analysis of GenoMic Annotation (MAGMA) (De Leeuw *et al*, 2015) as implemented in FUMA version 1.3.3d (Watanabe *et al*, 2017) to identify trait-specific and pleiotropic genes and gene-sets. MAGMA uses multi-regression to account for between-marker LD patterns and detect effects associated with multiple markers (De Leeuw *et al*, 2015). The European ancestry subset of the 1000 Genomes Project Phase3 (1000 Genomes Project Consortium *et al*, 2015) panel was used as reference, and FDR correction applied for all output *P* values.

#### Phenome-wide association study (PheWAS)

As part of our annotation workflow, we screened the 4,835 phenotypes available at FinnGen (R6), UKBB and EMBL-EBI GWAS Catalog via the Open Targets Genetics platform for associations previously reported for our lead variants using the otargen R package version 1.1.5 (Feizi & Ray, 2023)

#### Drug target screening

To further inform our chosen annotations and to identify potential drug-repurposing options, we queried approximately 17 000 drug molecules available at the Open Targets platform for clinical precedence of drugs with investigational or approved indications targeting any of our assigned candidate causal genes.

### Pair-wise and genome-wide genetic correlation

We performed bi-variate linkage disequilibrium score regression (LDSC) (Schizophrenia Working Group of the Psychiatric Genomics Consortium *et al*, 2015; ReproGen Consortium *et al*, 2015a) to compute genetic correlation between our trait-specific meta-analysed endpoints. Moreover, we estimated the genetic correlation for the trait-specific and composite multi-trait endpoints across other complex traits in a comprehensive set of 772 phenotypes based on GWAS experiments from European populations provided by the Integrative Epidemiology Unit’s (IEU) OpenGWAS Project. We applied dataset-level FDR correction to assess significance of the observed correlations and considered *P_FDR_*< 0.05 as significant.

### Colocalisation of GWAS and eQTL signals

We used Bayesian divisive clustering algorithm as implemented by the R package HyPrColoc version 0.0.2 (Foley *et al*, 2021) to determine whether our GWAS signals colocalise with expression quantitative trait loci (eQTLs). We applied HyPrColoc at a window of 1 MB (± 500 000 bases) around each novel lead variant on *cis*-eQTL data from eQTL Catalog release version 7 to test for colocalisation. Based on prior knowledge on relevant tissues and cell types, the analysis was performed for aortic artery, coronary artery, tibial artery, heart atrial appendage, heart left ventricle, whole blood from Genotype-Tissue Expression project V8 (GTEx Consortium *et al*, 2017)as well as for cultured fibroblasts. A posterior probability (PP) of >0.75 for colocalisation was defined as significant.

### Cell type heritability enrichment

We applied stratified LD score regression (sLDSC) (Schizophrenia Working Group of the Psychiatric Genomics Consortium *et al*, 2015; ReproGen Consortium *et al*, 2015b) for testing heritability enrichment in 21 relevant cell types across our GWAS meta-analyses and each multi-trait configuration.

The cis-element atlas (CATLAS) version 1 was used to obtain ATAC-seq peaks called individually from 222 adult human cell types. Specifically, we used the merged and filtered peak sets with fixed width of 400 bp to ensure more standardised result (Zhang *et al*, 2021). Cell-type-specific annotations and per cell type LD scores were built by mapping the genotypes of European ancestry in the 1000 Genomes Project Phase3 (1000 Genomes Project Consortium *et al*, 2015). A superset of all merged cell types was used a background control together with the version 2.3 of the baseline LD model. We applied FDR correction for each dataset respectively to account for multiple testing and considered *P_FDR_*< 0.05 as significant.

### Research ethics

Our research complies with all relevant ethical regulations. The UKBB resource has been approved by the UKBB Research Ethics Committee. MVP has received study protocol approval from the VA Central Institutional Review Board. All patients and control subjects have provided written informed consent for biobank studies. Full FinnGen ethical statement is included as supplementary material (Supplementary Note 1.).

### Data availability

Summary level GWAS datasets will be made publicly available in the European Bioinformatics Institute’s (EMBL-EBI) GWAS Catalog upon publication under accession XXX.

### Code availability

Our analyses were performed with the following open-source software: MTAG (23andMe Research Team *et al*, 2018), CPASSOC (Li & Zhu, 2017), HyPrColoc (Foley *et al*, 2021), bedtools (Quinlan & Hall, 2010), LDSC (Schizophrenia Working Group of the Psychiatric Genomics Consortium *et al*, 2015; ReproGen Consortium *et al*, 2015a, 2015b), FUMA (Watanabe *et al*, 2017), MAGMA (De Leeuw *et al*, 2015), PLINK (Purcell *et al*, 2007), ANNOVAR (Wang *et al*, 2010), CADD (Kircher *et al*, 2014), otargen (Feizi & Ray, 2023), rtracklayer (Lawrence *et al*, 2009), MungeSumstats (Murphy *et al*, 2021), Tidyverse (Wickham *et al*, 2019), Python version 2.7. and 3.9 (Van Rossum & Drake, 1995, 2007), R versions 4.2.2 and 4.3.1 (R Core Team, 2023), RStudio (Posit Team, 2025). Analysis code, scripts and notebooks used for the generation of this manuscript are available from the corresponding author upon reasonable request.

## RESULTS

### Study and analysis design

We sourced European ancestry population GWAS summary statistics data for CAD, AAA and TAA from FinnGen, UKBB and MVP. Together, these GWASs provide information on up to 200,000 individuals with CAD, 10,000 with AAA and 13,000 with TAA, each with approximately one million control subjects (Supplementary Table 1.). Furthermore, the number of investigated genotypes ranged between 21–23 million, resulting in abundance of genome-wide significant SNVs (Supplementary Table 2.). Following uniform data quality-control procedures (see Methods), we first performed inverse-variance weighted GWAS meta-analyses for CAD, AAA and TAA, followed by a three-way and three pair-wise multi-trait GWAS meta-analyses of each composite trait combination (Figure 1.). The resulting 12 summary statistics from all meta-analyses were then interrogated within five analysis modules designed to elucidate the shared and distinct genetic architectures of CAD and AAs (Figure 1.).

**Figure 1.**
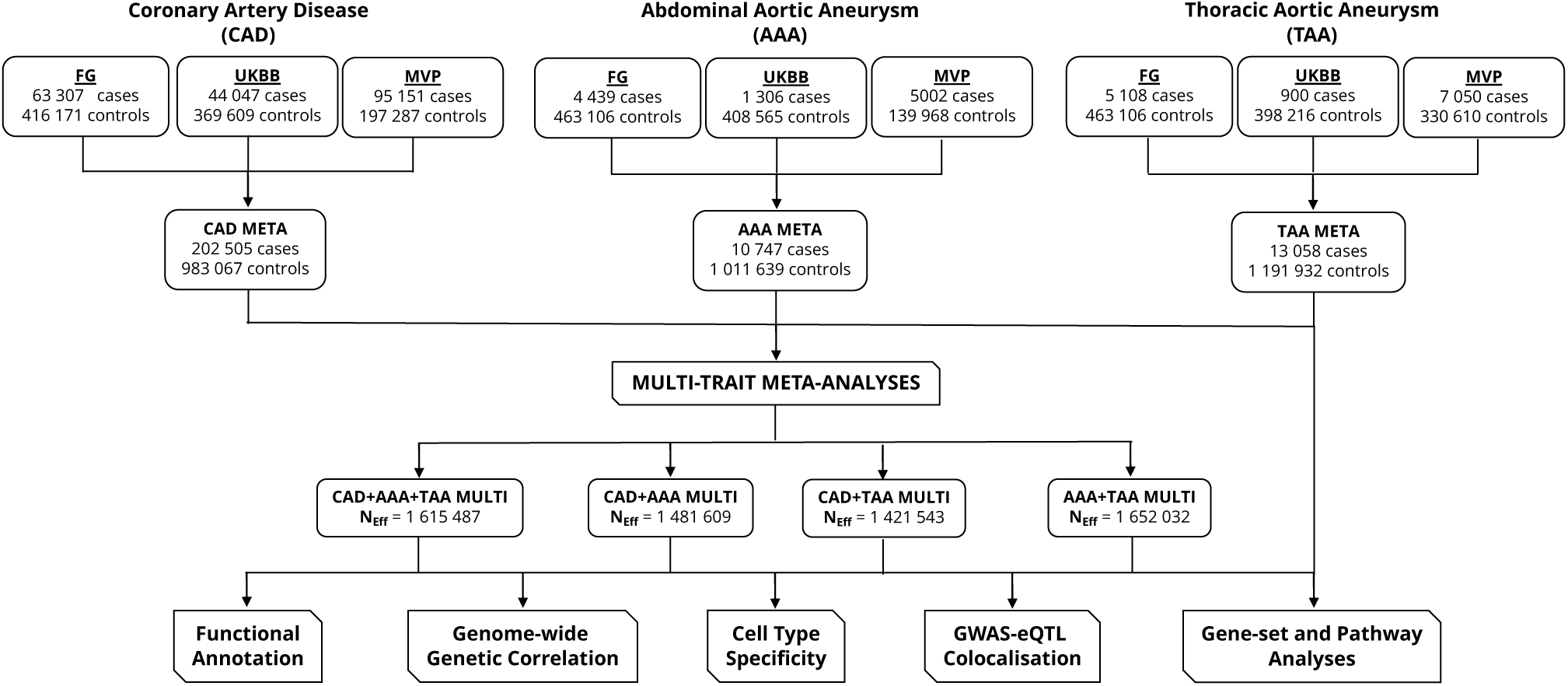
Schematic overview of study design. This study included GWAS summary statistics for CAD, AAA and TAA from FG, UKBB and MVP. The data were meta-analysed in two stages, first using inverse-variance weighting and then in four different multi-trait GWAS configurations. Subsequently, all meta-analyses were included in five distinct analysis modules comprising functional annotation, genome-wide genetic correlation, cell type specificity, GWAS-eQTL colocalisation and gene-set and pathway analyses. AAA, abdominal aortic aneurysm; CAD, coronary artery disease; eQTL, expression quantitative trait loci; FG, FinnGen; GWAS, genome-wide association study; META, meta-analysis; MULTI, multi-trait analysis; MVP, Million Veteran Program; NEff, effective sample size; TAA, thoracic aortic aneurysm; UKBB, United Kingdom Biobank.

### Cross-multi-trait GWAS discovery of 59 novel CAD, AAA and TAA loci

Across our 12 multi-trait GWAS meta-analyses with MTAG, we identified total of 59 previously unreported genomic regions, 17 for CAD, 15 for AAA and 27 for TAA, all located more than 1 MB apart from any previously reported lead variants (Supplementary Table 8.) (Figure 2.). MTAG estimated worst-case maximum false discovery rate (maxFDR) showed no evidence of inflation in estimates for CAD (maxFDR < 0.0003) or TAA (maxFDR < 0.04) in any multi-trait configuration. A degree of possible inflation was observed in our AAA multi-trait analyses (0.05 < maxFDR < 0.08), however, the mean chi-square values between AAA meta-analysis (χ^2^_AAA_ = 1.101) and MTAG analyses remained consistent, with the highest difference observed to the three-way multi-trait meta-analysis (χ^2^_CAD+AAA+TAA_ = 1.131). Moreover, our estimated genomic inflation factors for the three meta-analysed traits suggested inflation only for CAD (A = 1.304) but given the respective range of heritability intercept values remaining stable (0.9994 to 1.006), the observed inflation is likely influenced by true polygenic effects rather than confounding bias (Supplementary Table 4.). As a sensitivity analysis, we validated our MTAG results with a cross-trait GWAS meta-analyses using CPASSOC (see Methods). The CPASSOC analysis independently replicated the genome-wide significance of 8 out of 16 MTAG multi-trait associations for CAD, 12 of 15 for AAA, and 20 of 27 for TAA, further reinforcing the overall robustness of our findings (Figure 2.).

**Figure 2.**
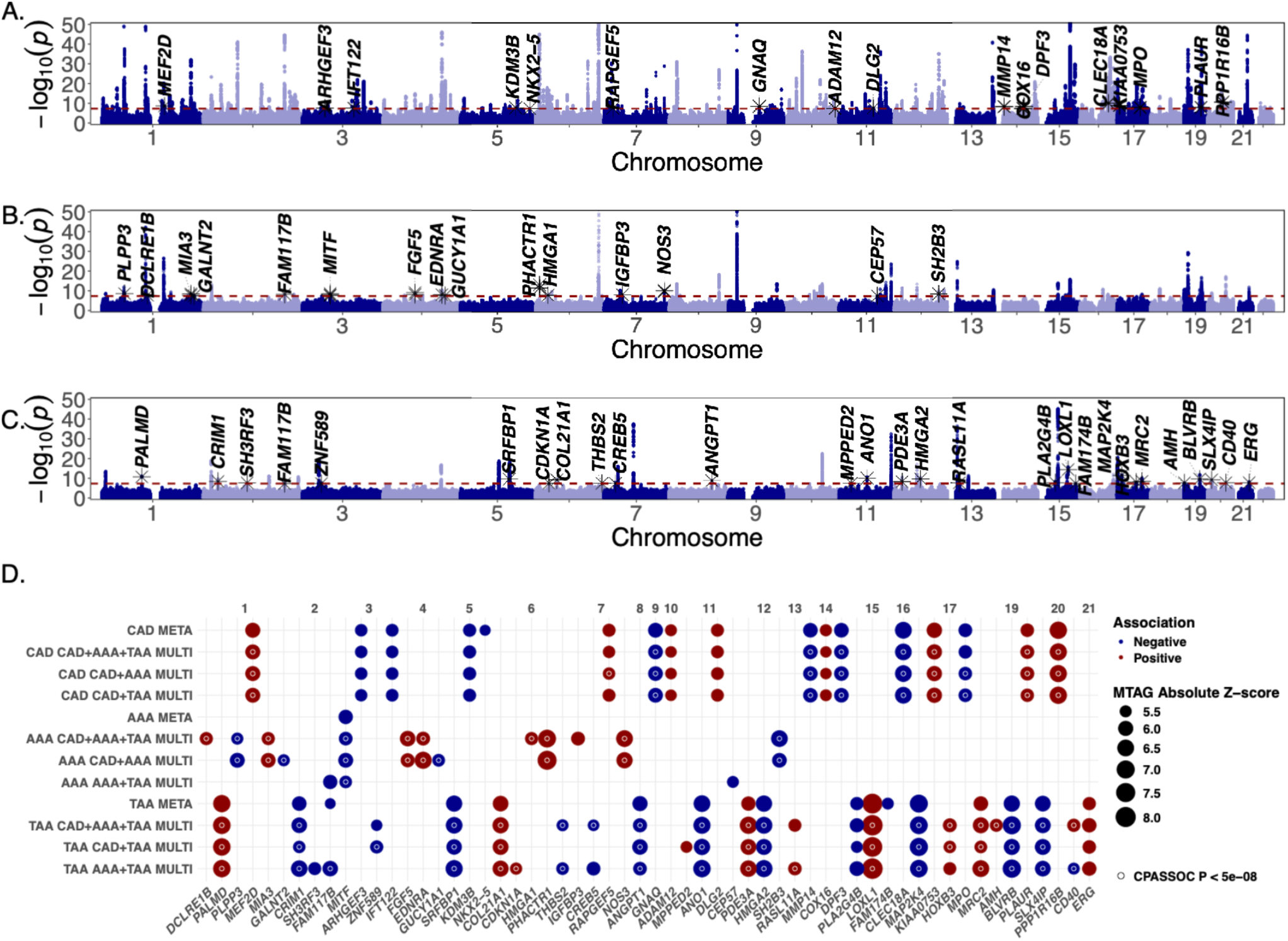
Summary of 59 novel loci identified in cross-multi-trait GWAS meta-analyses. (**A.**), (**B**.), (**C.**) Manhattan plots of previously unreported genome-wide significant (P < 5 x 10^-8^) regions associated with CAD, AAA and TAA, respectively. Individual single nucleotide variants are depicted as dots. Genomic loci are labelled with symbols for their annotated candidate gene. The X-axes represent genomic coordinates within the 22 autosomal chromosomes. The Y-axes display the strength of the association as -log10(P), restricted to maximum value of 50, with a dashed red line marking the threshold for genome-wide significance (P < 5 x 10^-8^). Alternating colour scale for easy viewing. (**D.**) A dot plot matrix representation of previously unreported genome-wide significant (P < 5 x 10^-8^) regions across all meta-analyses and multitrait configurations. Variants are grouped by chromosome on the top X-axis. The bottom X-axis lists the genomic loci labelled with symbols for their annotated candidate gene. Different trait configurations are shown on the Y-axis. Dot size corresponds to the strength of the association as represented by MTAG Z-score. Direction of the association denoted as red for positive and blue for negative. Associations reaching genome-wide significance (P < 5 x 10^-8^) in cross-trait CPASSOC sensitivity analysis are depicted with a white inner circle. AAA, abdominal aortic aneurysm; CAD, coronary artery disease; CPASSOC, cross-phenotype association analysis; GWAS, genome-wide association study; META, meta-analysis; MTAG, Multi-trait Analysis of GWAS, MULTI, multi-trait analysis; TAA, thoracic aortic aneurysm.

### Characterisation of novel association signals

We mapped our novel loci to likely causal genes by manually curating various platforms for genomic annotations (see Methods). In concordance with previous reports, our CAD loci converged on genes related to cardiac development, vascular tone and function, and inflammation (Supplementary Table 8.). For example, signalling mediators such as *ARHGEF3*, *RAPGEF5*, and *GNAQ* have been implicated for modulating vascular tone and endothelial integrity, potentially influencing atherosclerotic progression and post-injury repair (Serbanovic-Canic *et al*, 2011; Duan *et al*, 2025; Sasaki *et al*, 2022). *MMP14*, *ADAM12*, and *PLAUR* highlight the role of extracellular matrix remodelling in plaque destabilization and myocardial remodelling following ischemic injury, while *MPO*-driven inflammatory responses further exacerbate tissue damage and chronic vascular inflammation (Barnes *et al*, 2017; Dokun *et al*, 2015; Dai & Lin, 2023). Moreover, CAD signals observed near transcription factors *NKX2-5* and *MEF2D* are likely explained by links to cardiac morphogenesis and contractile function (Cao *et al*, 2023; Lu *et al*, 2021).

Candidate genes for AAA loci were most prominently related to lipid metabolisms, blood pressure and endothelial function. Genes such as *PLLP3* and *GALNT2* modulate lipid signalling, potentially impacting atherosclerotic processes (Mueller *et al*, 2019; Antonucci *et al*, 2022), whereas *HMGA1*, *IGFBP3*, and *MITF* all contribute to the fine-tuning of gene expression in response to metabolic stress (Arce-Cerezo *et al*, 2015; Yamada *et al*, 2010; Saha *et al*, 2006). *EDNRA*, *GUCY1A1*, and *NOS3* are pivotal in regulating vasodilatation and vasoconstriction (Gupta, 2023; Mauersberger *et al*, 2022; Pautz *et al*, 2021). Moreover, we identified *PHACTR1*, a well-known locus for CAD risk (Ma *et al*, 2022), as a potentially pleiotropic association for AAA.

Lastly for TAA we observed signals near *ANGPT1*, *THBS2*, *LOXL1*, and *COL21A1* that contribute to extracellular matrix remodelling, critical in maintaining vessel stability and influencing plaque composition (Brindle *et al*, 2006; Huang *et al*, 2023; Martínez-González *et al*, 2019; Chou & Li, 2002). Key cell cycle and stress regulators like *CDKN1A*, *MAP2K4*, and *HMGA2* mediate responses to hemodynamic stress and repair processes (Nayak *et al*, 2018; Yue & López, 2020; Wang *et al*, 2021), while transcription factors including *ERG* and *CREB5* orchestrate gene expression programs essential for endothelial cell function and vascular homeostasis (Birdsey *et al*, 2015; Jeon *et al*, 2007). Additionally, signalling modulators such as *SRFBP1*, *RASL11A*, and *PDE3A* integrate critical pathways that govern vascular tone and smooth muscle cell activity (Aberdeen *et al*, 2021; Martinez *et al*, 2022; Begum *et al*, 2011).

### Functional analyses

To explore functional pathway patterns associated with CAD, AAA and TAA, we performed MAGMA (De Leeuw *et al*, 2015) gene-set enrichment analysis using curated sets and Gene Ontology (GO) terms from MsigDB (Liberzon *et al*, 2011). Out of the 17 009 pathways tested, 1315, 542 and 440 were significantly associated for CAD, AAA and TAA, respectively, following FDR correction (Supplementary Table 13.–15.). Our strongest gene-sets for CAD underlined the importance of lipid metabolism as well as heart and circulatory system development processes consistently identified in previous studies (Figure 3.). AAA was likewise linked most prominently to lipid-related pathways and displayed general increase in their enrichment in multi-trait configurations involving CAD, and conversely when analysed pairwise with TAA (Figure 3.). For TAA, associations were notably distinct, involving gene-sets related to formation of elastic fibres, cardiac system and muscle development, and blood vessel morphology (Figure 3.).

**Figure 3.**
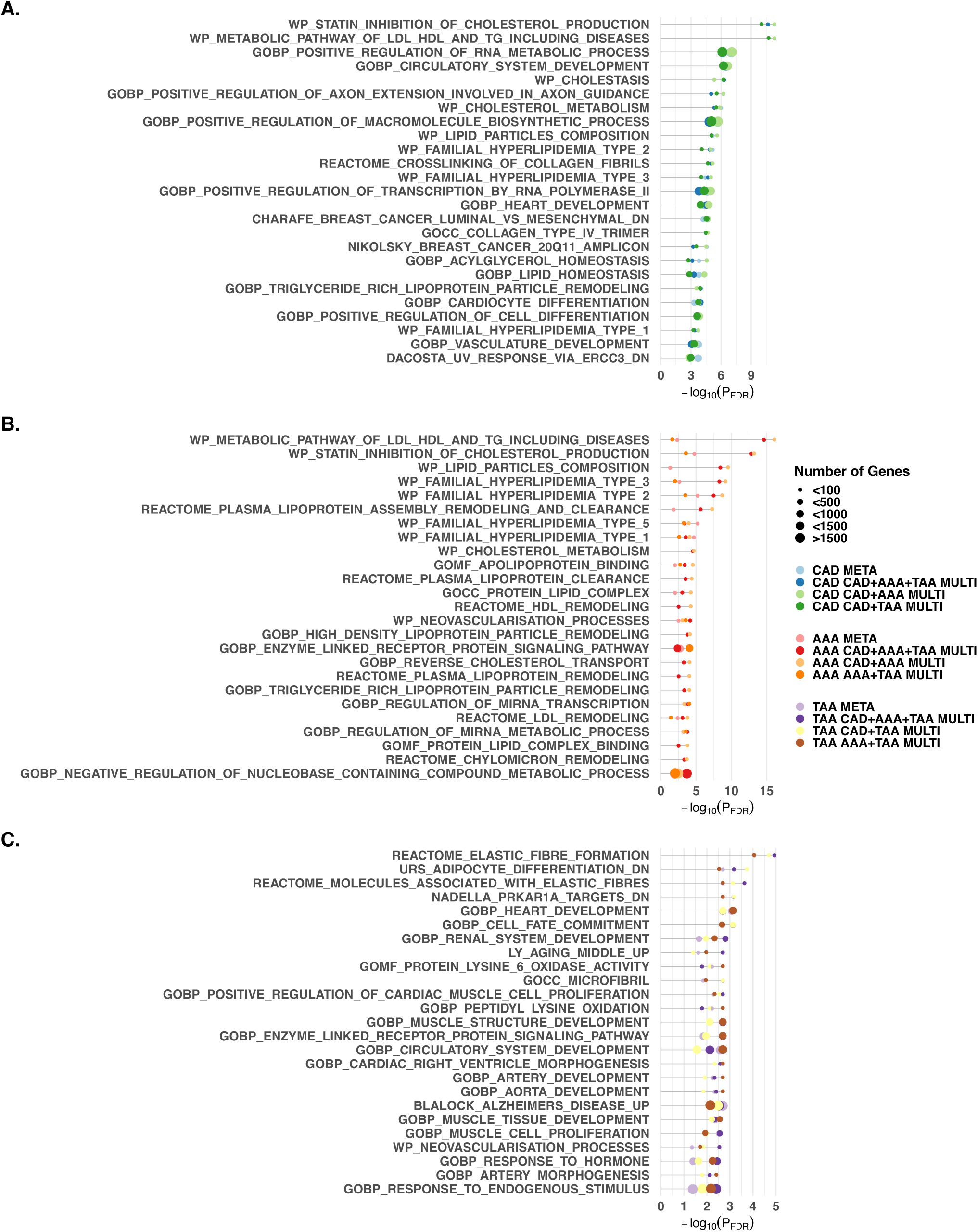
Summary of the strongest functional gene-set associations identified by MAGMA. (**A.**), (**B**.), (**C.**) top 25 significant (PFDR ≤ 0.05) gene-sets for CAD, AAA and TAA, respectively. Associations ranked by strength defined as -log10(PFDR), and number of genes in the set. Sticks on the X-axes represent the strength of the associations as -log10(PFDR). Gene-set identifiers listed on the Y-axis. Dot size corresponds to the number of genes within a set, in intervals of <100, <500, <1000, <1500 and >1500 genes. Dot coloured per different analysis trait configurations. AAA, abdominal aortic aneurysm; CAD, coronary artery disease; FDR, false discovery rate; META, meta-analysis; MULTI, multi-trait analysis. TAA, thoracic aortic aneurysm.

We further interrogated the pleiotropic associations of our novel lead variants by screening the approximately 5,000 phenotypes available at the Open Targets Genetics platform. Unsurprisingly, several of independent CAD variants were prominently associated with different haematological traits, cardiometabolic traits and risk factors, followed by anthropometric and cancer related traits (Supplementary Table 8.). The phenome-wide association patterns of our AAA lead variants were similarly linked to other cardiometabolic as well as haematological traits. Moreover, AAA variants had broader pleiotropic associations of infectious and inflammatory nature (Supplementary Table 8.).

TAA likewise displayed most associations towards various blood cell measurements and other diseases of the circulatory system, followed by traits related to the metabolic system and cancer (Supplementary Table 8.).

Via our annotation process, we identified 20 drug molecules targeting the protein product of our assigned candidate genes (Supplementary Table 8.). In addition to the now discontinued Rebimastat (Li *et al*, 2023), we found Verdiperstat as a potential drug candidate targeting *MPO*. Verdiperstat is an irreversible myeloperoxidase inhibitor under investigation as a potential disease-modifying therapy for neurodegenerative diseases (Writing Committee for the HEALEY ALS Platform Trial *et al*, 2025). Several endothelin receptor antagonists (Ambrisentan, Bosentan, Macitentan) were indicated for AAA candidate genes. These agents block vasoconstrictive endothelin-1 (ET-1) and are widely used to treat pulmonary arterial hypertension (PAH) (Enevoldsen *et al*, 2020). In turn, soluble guanylate cyclase (sGC) stimulators are vasodilators enhancing nitric oxide–cGMP signalling and target pulmonary or systemic vascular disease (Follmann *et al*, 2017). The drugs Aminophylline, Cilostazol and Anagrelide, highlighted for TAA, belong to the phosphodiesterase (PDE) inhibitors and adenosine antagonist category used to treat COPD and intermittent claudication in peripheral arterial disease (Bondarev *et al*, 2022). Furthermore, Levosimendan, indicated for acute decompensated heart failure, is an inodilator that sensitises cardiac myofilaments to calcium and opens ion channels in VSMCs (Burkhoff *et al*, 2021).

### Genetic correlations

By applying LD score regression on our meta-analysed summary statistics, we estimated the between-trait genetic correlations (r_g_) of CAD, AAA and TAA to be 0.29 (*P* < 5.81×10^-10^) and 0.06 (*P* < 0.07), respectively, and 0.41 (*P* < 4.25×10^-18^) between AAA and TAA (Supplementary Table 9.).

When observing correlations to other complex disorders and traits genome-wide level, we consistently saw well-established, positive correlations between CAD and other cardiovascular conditions, known risk factors such as LDL-C levels, smoking, blood pressure, and statin medication use (Supplementary Figure 1., Supplementary Table 10.). Similarly, we found AAA to correlate with other cardiovascular events, smoking, blood pressure, lipid levels and related medication (Supplementary Figure 2., Supplementary Table 11.). TAA presented the most prominent correlations to blood pressure traits and family member health indicators. Interestingly, unspecified haematuria (R31) was observed to among the highest positively correlated traits. Significant common correlation patterns for CAD and AAs were considerably more sparse and predominantly related to blood pressure traits and parental longevity (Figure 4., Supplementary Table 11.–12.).

**Figure 4.**
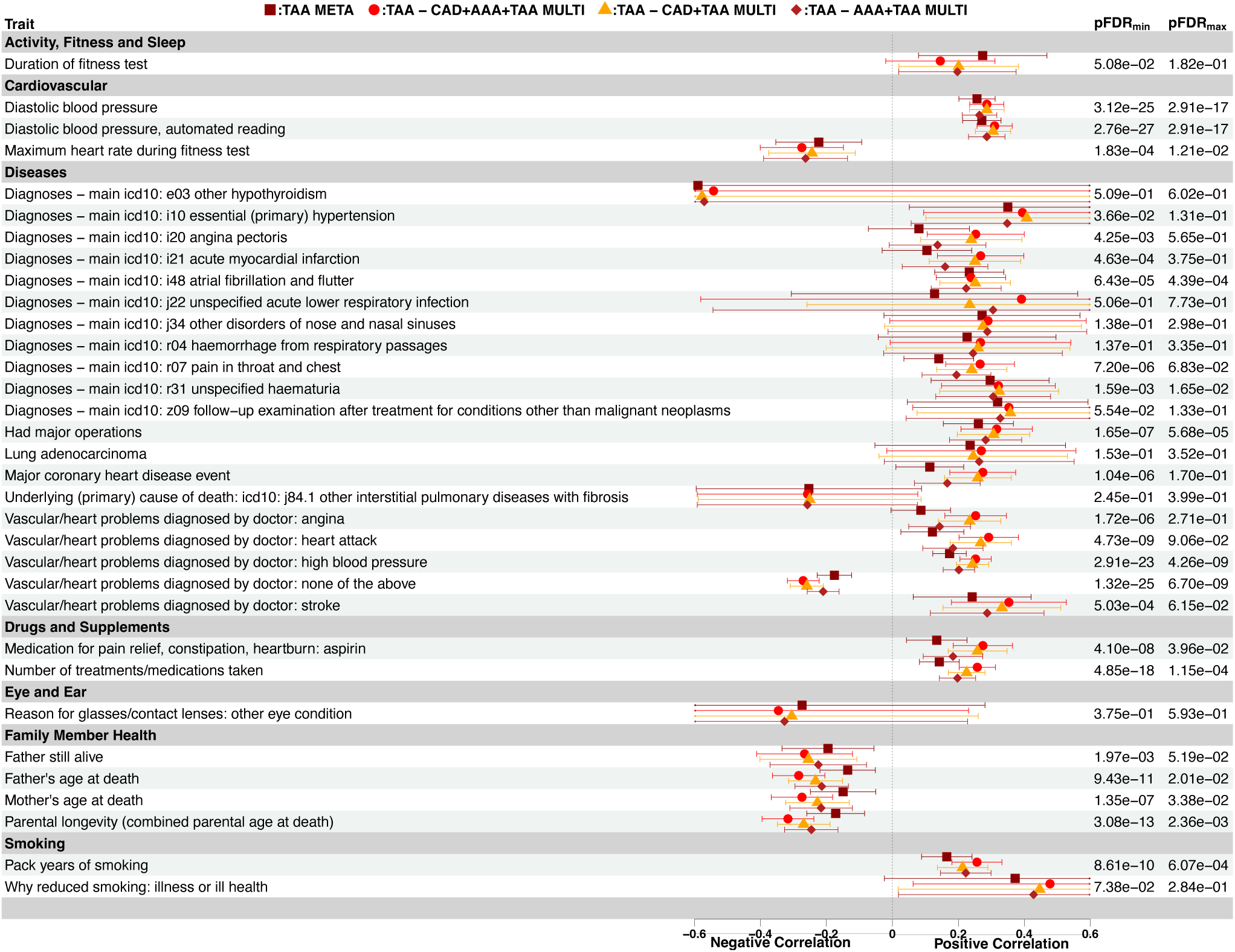
Summary of genome-wide genetic correlation patterns for TAA. Forest plot of genetic correlations computed between TAA and other complex traits with LDSC. Correlated traits are shown on the left column, divided to related categories. Genetic correlation (rg) and the associated 95% confidence intervals are presented in the center panel. Trait configurations indicated by coloured symbols. The maximum and minimum FDR-adjusted P values for each correlation across different trait configurations are listed on right-side columns. Only the strongest (-0.25 > rg > 0.25) are shown on the figure. AAA, abdominal aortic aneurysm; CAD, coronary artery disease; FDR, false discovery rate; LDSC, linkage disequilibrium score regression, META, meta-analysis; MULTI, multi-trait analysis; rg, genetic correlation; TAA, thoracic aortic aneurysm.

### Tissue and cell type specificity

Next, we applied stratified LD score regression (sLDSC) on activity profiles of candidate *cis*-regulatory elements (cCREs) from the CATLAS version 1 database to evaluate whether CAD, AAA and TAA heritability explained by genome-wide common genetic variants were significantly enriched for certain cell types. With this approach, we found CAD signals to be consistently most enriched in fibroblasts, pericytes, vascular smooth muscle cells (VSMCs) and endothelial cell types. AAA displayed enrichment patterns reminiscent of CAD, apart from macrophages and ventricular cardiomyocytes, the former appearing significant only for AAA in all but the three-way multi-trait configuration, whereas the latter were found to significant specifically for CAD. Furthermore, in our analysis adipocytes were significantly enriched for AAA only in multi-trait settings. Lastly, TAA presented considerably different from CAD and AAA, its signals distinctively enriched for fibroblasts and VSMCs, with enrichment in pericytes observed at joint analysis with AAA (Figure 5.). Due to limitations in the LDSC model, a low-level of non-significant negative enrichment was observed for cell types with smaller annotations (Supplementary Table 16.). Secondly, we also observed the enrichment of CAD, AAA and TAA signals in proximity of tissue-specific gene expression profiles in

**Figure 5.**
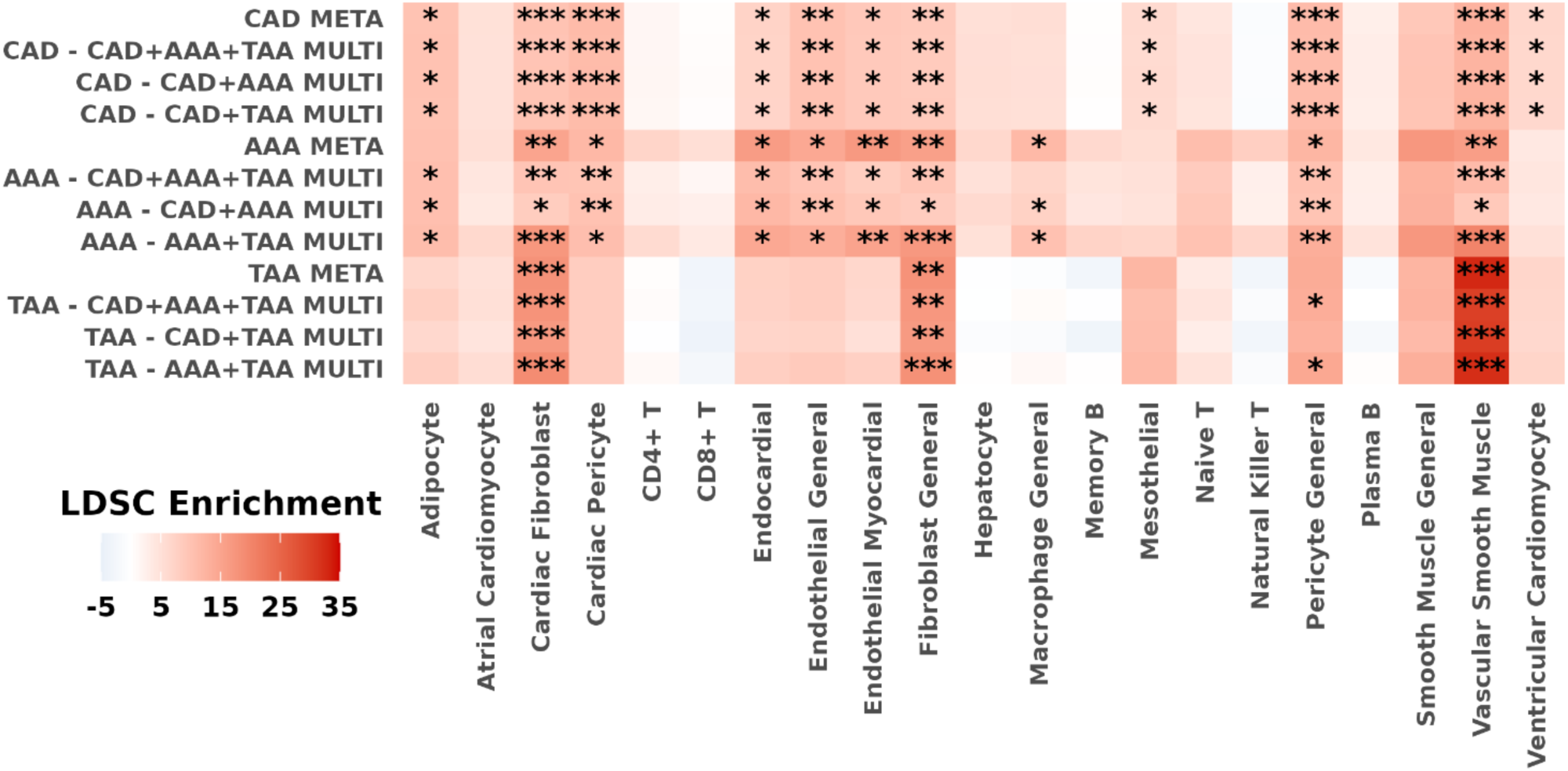
Overview of GWAS heritability enrichment in relevant human cell types. Heatmap shows the enrichment of GWAS risk variants across different trait configurations (Y-axis) in human cell-type-resolved cCREs (X-axis) computed using stratified LDSC. FDR-adjusted P of the LDSC coefficient denoted as * PFDR < 0.05, ** PFDR < 0.01, *** PFDR < 0.001. AAA, abdominal aortic aneurysm; CAD, coronary artery disease; cCRE, candidate cis-regulatory DNA element; FDR, false discovery rate; GWAS, genome-wide association study; LDSC, linkage disequilibrium score regression; META, meta-analysis; MULTI, multi-trait analysis; TAA, thoracic aortic aneurysm.

Genotype-Tissue Expression (GTEx V8) project (GTEx Consortium *et al*, 2017). Using gene expression data from multiple tissues, we found the three types of arterial tissues, coronary, aortic and tibial to be most significantly enriched for CAD. Though our AAA meta-analysis lacked the power to reach significant enrichment in any tissue type, the multi-trait configurations showed most prominent enrichment for different arterial tissues as well as for uterus and lung. Lastly, the gene expression profile for TAA was similarly highly enriched for arterial tissues and considerably less enriched in different reproductive tissues and lung (Supplementary Figure 3. – 14.).

### Colocalisation with eQTL signals

Finally, we inspected the colocalisation of our GWAS signals with tissue-specific molecular eQTLs to compute posterior probabilities for a given SNV to be associated both with disease risk and altered gene expression. To this end we applied HyPrColoc on a 1 MB (± 500 000 bases) window around GWAS lead variants in seven CVD-relevant tissues derived from the Genotype-Tissue Expression (GTEx) V8 project and eQTL Catalog. Approximately 17% of the novel loci across all meta-analyses and multi-trait analyses demonstrated significant colocalisation (PP ≥ 75%) with an eGene in relevant tissues, implying the correlated variant and/or variants in its proximity simultaneously influence gene expression and disease risk at the region (Supplementary Table 17.). For CAD, we consistently detected a strong colocalisation at *CLEC18A* in GTEx coronary artery eQTLs (PP ≥ 0.99). No colocalisations were detected for our AAA meta-analysis, but different multi-trait configurations indicated signals at *MIA3* to colocalise in ventricular heart (PP ≥ 0.87) and visceral adipose (PP ≥ 0.91). Considerable evidence was found for our TAA signals to colocalise at several key tissues, for instance, *PLA2G4B* at atrial (PP ≥ 0.92) and ventricular heart (PP ≥ 0.91) as well as visceral adipose (PP ≥ 0.85). Moreover, *ZNF589* and *AMH* both were indicated for multiple tissues at a significant level. Lastly, only AA multi-trait locus to display likely colocalisation was *CEP57* in ventricular heart (PP > 0.92) (Figure 6.)

**Figure 6.**
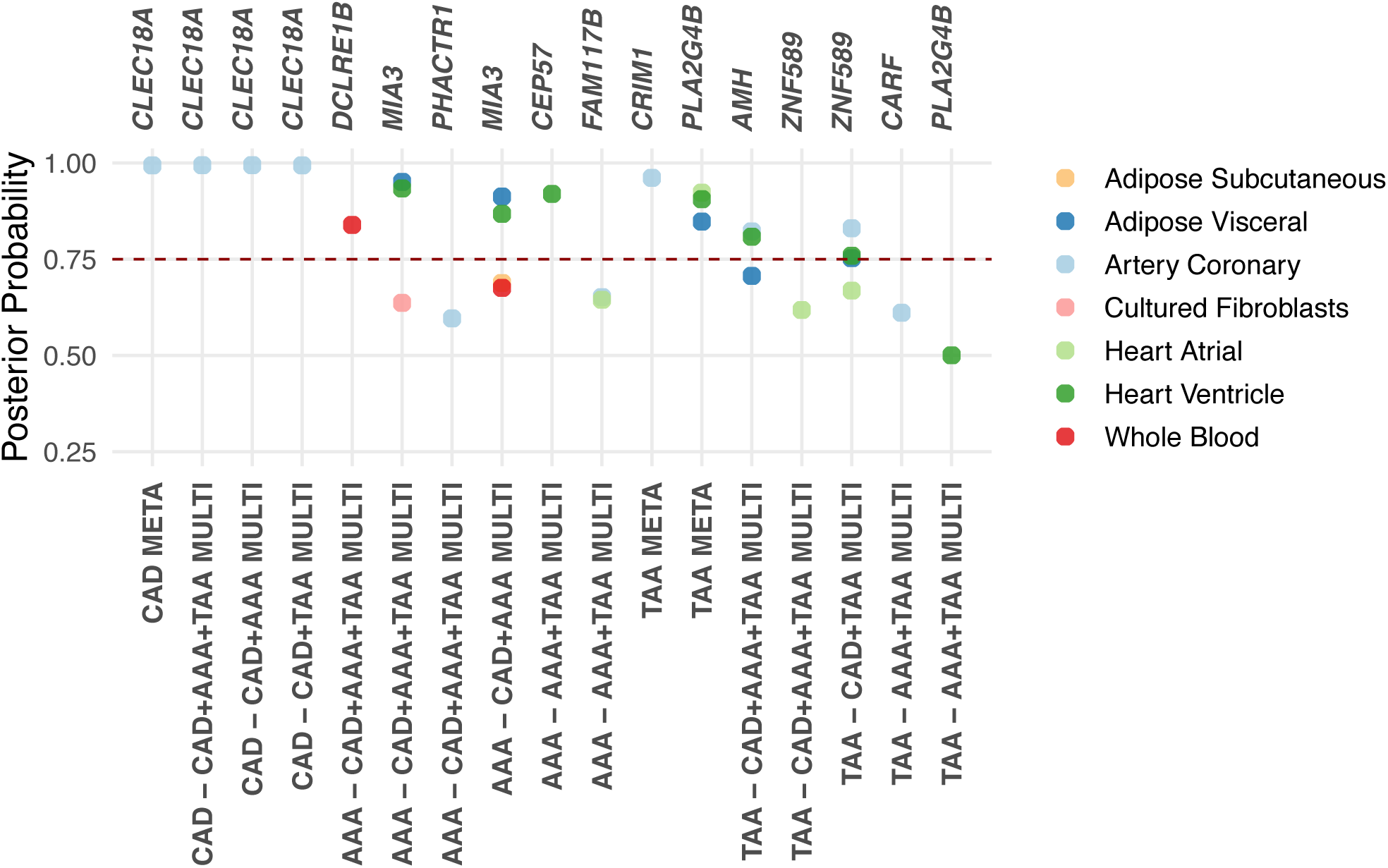
Colocalisation of novel GWAS signals with eQTLs in relevant human tissues and cell types. A dot plot displaying the colocalisation PP between novel loci and human tissue or cell-type derived eQTLs. The X-axes depict the strongest candidate loci for colocalisation as estimated by HyPrColoc (top) in different GWAS trait-configurations (bottom). Colour denotes the colocalised GTEx V8 tissue or cell type. Threshold for significant colocalisation (PP > 0.75) marked as a dashed red horizontal line. AAA, abdominal aortic aneurysm; CAD, coronary artery disease; eQTL, expression quantitative loci; GTEx, Genotype-Tissue Expression Project, GWAS, genome-wide association study; META, meta-analysis; MULTI, multi-trait analysis; PP, posterior probability; TAA, thoracic aortic aneurysm.

## DISCUSSION

Here, we report genome-wide multi-trait meta-analyses of CAD, AAA and TAA. Our analyses into genetic their relationship yielded improved power to detect 59 previously unreported loci for this collection of traits and implicated both known cardiovascular and previously unreported variants involved in the pathophysiology of thereof. In further downstream analyses, we highlighted trait-specific and pleiotropic variants and their likely causal genes, biological pathways, and patterns for key cell and tissue types for future study of atherosclerotic vascular disease.

First, our study underscores the key value of leveraging multivariate GWAS in analyses of complex phenotypes to improve genetic discovery. While large-scale single-trait GWAS continue to identify novel genetic determinants for disease, incorporating traits with overlapping genomic risk factors add greatly to our understanding of phenotypes with smaller sample sizes and aid in contextualising new discoveries. Across our analyses, we identified multiple variants previously linked to various cardiometabolic traits, many of which are risk factors for CAD and AA. The continued growth of resources linking genomic data to electronic health records, laboratory measurement and clinical imaging, will allow for the characterisation of even more refined phenotypes, and further empowering future multi-trait efforts.

Indeed, our findings acutely demonstrate the potential gains of integrating GWASs of correlated traits from studies of varying sample sizes. Many of our discoveries recapitulate the well-established paradigm of studying genetic variation of cardiovascular diseases in large population cohorts, where the same effector genes and pathways are replicated withing multiple related traits. For example, recent multi-trait GWAS of heart failure established risk variants previously associated with several other cardiometabolic risk factors (Zhu *et al*, 2024). In our work, these common patterns were particular evident from the overlaps in genetic correlation and in the enrichment of functional phenotype categories.

Supported by multiple lines of evidence, we implicate both known and previously unreported candidate genes and pathways to be associated with CAD, AAA and TAA. Our annotations converge on gene sets involved in VSMC contractile function, NO signalling, endothelial cell signalling and lipid uptake. For example, *ANO1* is essential for function of calcium-activated chloride channel in muscle, and associated genetic variation has been previously linked to aortic distensibility and size and bloop pressure. Moreover, we linked a novel CAD association to *CLEC18A*, a member of the C-type lectin 18 family, ubiquitously expressed in several tissues, with the variation in the gene previously found to promote metabolic and proliferative shifts, as well as immune deactivation in human cells (Hsu *et al*, 2022).

Overall, for CAD and AAA we observed, unsurprisingly, significant enrichment of genes involved in processing of lipoproteins. In TAA, the strongest shared pathways encompassed formation of elastic fibres as well as heart and circulatory system development. Taken together, our results demonstrate well the utility of integrating genomic data, drawing from genetic associations, gene expression changes and tissue- and cell-type-specific sources, to assign effector genes for disease. The findings here also support the present notion of pleiotropic contributors to TAA (Klarin *et al*, 2023). Lastly, the discovery of shared disease genes and pathways may be transformed into new treatment proposals. Most drug candidates we discovered have been used to treat other CVDs, and some are in clinical trials for repurposing.

Of note, our heritability enrichment analyses reinforced the shared significance of VSMCs and fibroblasts in CAD and AA pathophysiology, and further pinpointed pericytes as one the most enriched common cell type. Like VSMCs, pericytes are essential for maintaining endothelial stability during angiogenesis (Chiaverina *et al*, 2019), acting together with endothelial cells to form the vascular basement membrane (Su *et al*, 2021). The role of pericytes in modulating intercellular signals of proliferation and migration poises them to play an integral role in both CAD and AAs. Together these observations highlight the need for further examinations into pericyte biology within the cardiovascular context.

The strengths of the current analysis are multiple. First, the inclusion of large-scale cohorts included in our meta-analysis led us to identify novel variants and putative effector genes for CAD, AAA and TAA, and expand them further through our multivariate GWAS analyses. Second, we control against the potential inflation of our observed multi-trait associations by joint cross-trait implementation.

Thirdly, we applied multiple complementary annotations strategies, state-of-the-art pipelines, eQTL, colocalization, gene-based together with extensive manual curation, to assign the most likely gene responsible for our GWAS signals.

Nevertheless, we interpret our findings in context of their limitations, of which this study has several. First, the GWAS allows only the analysis of common SNVs but excludes other variant types, such as rare variants, short insertions and deletions, and structural variants. Second, CVDs encompass a wide range of diseases and physiological traits, and the ones analysed here represent only a subset. Third, our ability to infer molecular and cellular mechanisms is constrained by the available reference knowledgebases and databases, namely most downstream investigations were limited to use of a European reference panel. Additionally, further larger meta-analysis or independent studies are needed to validate these findings. Overall, the multivariate study design allows the detection of shared genetic factors, yet the approach has limited value to identify genetic determinants specific to a given trait. Further functional research is required to explore causal variants and underlying mechanisms at the discovered loci and enhance our ability to understand genetic mechanisms underlying shared and distinct CVD risk.

In conclusion, we are to our knowledge the first to perform, a large-scale multivariate discovery GWAS meta-analysis into the shared genetic architecture of CAD, AAA and TAA. Our findings not only help to elucidate the common genetic aetiology between CAD and AAs but also provide potential future therapeutic and diagnostic avenues.

## ACKNOWLEDGEMENTS

This study was supported by funding from the Finnish Foundation for Cardiovascular Research, Sigrid Juselius Foundation and the Medical Research Center of Oulu University Hospital and University of Oulu. M.U.K was co-supported by the European Union (ERC, SECRET, 101126115). The authors acknowledge the CSC – IT Center for Science, Finland, and UEF Bioinformatics Center and Biocenter Kuopio, University of Eastern Finland, Finland for computational resources and infrastructure. We thank all the participants and investigators of the FinnGen Study. The FinnGen project is funded by two grants from Business Finland (HUS 4685/31/2016 and UH 4386/31/2016) and the following industry partners: AbbVie Inc., AstraZeneca UK Ltd, Biogen MA Inc., Bristol Myers Squibb Inc. (and Celgene Corporation & Celgene International II Sàrl), Genentech Inc., Merck Sharp & Dohme LCC, Pfizer Inc., GlaxoSmithKline Intellectual Property Development Ltd., Sanofi US Services Inc., Maze Therapeutics Inc., Johnson&Johnson Innovative Medicine Inc., Novartis AG, Boehringer Ingelheim International GmbH and Bayer AG. Following biobanks are acknowledged for delivering biobank samples to FinnGen: Auria Biobank (www.auria.fi/biopankki), THL Biobank (www.thl.fi/biobank), Helsinki Biobank (www.helsinginbiopankki.fi), Biobank Borealis of Northern Finland (https://www.ppshp.fi/Tutkimus-ja-opetus/Biopankki/Pages/Biobank-Borealis-briefly-in-English.aspx), Finnish Clinical Biobank Tampere (www.tays.fi/en-US/Research_and_development/Finnish_Clinical_Biobank_Tampere), Biobank of Eastern Finland (www.ita-suomenbi-opankki.fi/en), Central Finland Biobank (www.ksshp.fi/fi-FI/Potilaalle/Biopankki), Finnish Red Cross Blood Service Biobank (www.veripalvelu.fi/verenluovutus/biopankkitoiminta), Terveystalo Biobank (www.terveystalo.com/fi/Yritystietoa/Terveystalo-Biopankki/Biopankki/) and Arctic Biobank (https://www.oulu.fi/en/university/faculties-and-units/faculty-medicine/northern-finland-birth-cohorts-and-arctic-biobank). All Finnish Biobanks are members of BBMRI.fi infrastructure (https://www.bbmri-eric.eu/national-nodes/finland/). Finnish Biobank Cooperative -FINBB (https://finbb.fi/) is the coordinator of BBMRI-ERIC operations in Finland. The Finnish biobank data can be accessed through the Fingenious® services (https://site.fingenious.fi/en/) managed by FINBB.

## SUPPLEMENTARY MATERIAL

**Supplementary Note 1.** FinnGen R12 ethics statement

**Supplementary Note 2.** Competing interests statement

**Supplementary Figure 1.** Summary of genome-wide genetic correlation patterns for CAD

**Supplementary Figure 2.** Summary of genome-wide genetic correlation patterns for AAA

**Supplementary Figure 3.** Tissue-specific gene expression profile enrichment of CAD in MTAG meta-analysis

**Supplementary Figure 4.** Tissue-specific gene expression profile enrichment of CAD in MTAG CAD+AAA+TAA multi-trait meta-analysis

**Supplementary Figure 5.** Tissue-specific gene expression profile enrichment of CAD in MTAG CAD+AAA multi-trait meta-analysis

**Supplementary Figure 6.** Tissue-specific gene expression profile enrichment of CAD in MTAG CAD+TAA multi-trait meta-analysis

**Supplementary Figure 7.** Tissue-specific gene expression profile enrichment of AAA in MTAG meta-analysis

**Supplementary Figure 8.** Tissue-specific gene expression profile enrichment of AAA in MTAG CAD+AAA+TAA multi-trait meta-analysis

**Supplementary Figure 9.** Tissue-specific gene expression profile enrichment of AAA in MTAG CAD+AAA multi-trait meta-analysis

**Supplementary Figure 10.** Tissue-specific gene expression profile enrichment of AAA in MTAG AAA+TAA multi-trait meta-analysis

**Supplementary Figure 11.** Tissue-specific gene expression profile enrichment of TAA in MTAG meta-analysis

**Supplementary Figure 12.** Tissue-specific gene expression profile enrichment of TAA in MTAG CAD+AAA+TAA multi-trait meta-analysis

**Supplementary Figure 13.** Tissue-specific gene expression profile enrichment of TAA in MTAG CAD+TAA multi-trait meta-analysis

**Supplementary Figure 14.** Tissue-specific gene expression profile enrichment of TAA in MTAG AAA+TAA multi-trait meta-analysis

**Supplementary Figure 15.** Regional association plot of previously unreported TAA signal at 6q27 around the GWAS lead variant rs1322640

**Supplementary Figure 16.** Regional association plot of previously unreported CAD signal at 11q14.1 around the GWAS lead variant rs567007

**Supplementary Figure 17.** Regional association plot of previously unreported TAA signal at 2p22.2 around the GWAS lead variant rs848549

**Supplementary Figure 18.** Regional association plot of previously unreported CAD signal at 5q31.2 around the GWAS lead variant rs11740931

**Supplementary Figure 19.** Regional association plot of previously unreported TAA signal at 15q26.1 around the GWAS lead variant rs8025293

**Supplementary Figure 20.** Regional association plot of previously unreported CAD signal at 10q26.2 around the GWAS lead variant rs11244896

**Supplementary Figure 21.** Regional association plot of previously unreported TAA signal at 20q13.12 around the GWAS lead variant rs6131010

**Supplementary Figure 22.** Regional association plot of previously unreported CAD signal at 5q35.1 around the GWAS lead variant rs10071514

**Supplementary Figure 23.** Regional association plot of previously unreported TAA signal at 17q23.2 around the GWAS lead variant rs740698

**Supplementary Figure 24.** Regional association plot of previously unreported AAA signal at 7p12.3 around the GWAS lead variant rs6969255

**Supplementary Figure 25.** Regional association plot of previously unreported TAA signal at 19q13.2 around the GWAS lead variant rs113473882

**Supplementary Figure 26.** Regional association plot of previously unreported AAA signal at 4q21.21 around the GWAS lead variant rs12509595

**Supplementary Figure 27.** Regional association plot of previously unreported TAA signal at 15q24.1 around the GWAS lead variant rs893816

**Supplementary Figure 28.** Regional association plot of previously unreported CAD signal at 14q24.2 around the GWAS lead variant rs6573939

**Supplementary Figure 29.** Regional association plot of previously unreported CAD signal at 14q11.2 around the GWAS lead variant rs17880989

**Supplementary Figure 30.** Regional association plot of previously unreported CAD signal at 19q13.31 around the GWAS lead variant rs4760

**Supplementary Figure 31.** Regional association plot of previously unreported TAA signal at 20p12.2 around the GWAS lead variant rs857011

**Supplementary Figure 32.** Regional association plot of previously unreported TAA signal at 1p21.2 around the GWAS lead variant rs7543039

**Supplementary Figure 33.** Regional association plot of previously unreported TAA signal at 12q14.3 around the GWAS lead variant rs1979440

**Supplementary Figure 34.** Regional association plot of previously unreported TAA signal at 6p12.1 around the GWAS lead variant rs1925145

**Supplementary Figure 35.** Regional association plot of previously unreported CAD signal at 9q21.2 around the GWAS lead variant rs17725735

**Supplementary Figure 36.** Regional association plot of previously unreported AAA signal at 12q24.12 around the GWAS lead variant rs10774625

**Supplementary Figure 37.** Regional association plot of previously unreported CAD signal at 1q22 around the GWAS lead variant rs1109751

**Supplementary Figure 38.** Regional association plot of previously unreported AAA signal at 1p13.2 around the GWAS lead variant rs10858023

**Supplementary Figure 39.** Regional association plot of previously unreported TAA signal at 17p12 around the GWAS lead variant rs11869087

**Supplementary Figure 40.** Regional association plot of previously unreported TAA signal at 11p14.1 around the GWAS lead variant rs10835708

**Supplementary Figure 41.** Regional association plot of previously unreported AAA signal at 2q33.2 around the GWAS lead variant rs146902012

**Supplementary Figure 42.** Regional association plot of previously unreported TAA signal at 17q21.32 around the GWAS lead variant rs72829868

**Supplementary Figure 43.** Regional association plot of previously unreported AAA signal at 1q42.13 around the GWAS lead variant rs11122456.

**Supplementary Figure 44.** Regional association plot of previously unreported CAD signal at 3p14.3 around the GWAS lead variant rs6792170

**Supplementary Figure 45.** Regional association plot of previously unreported TAA signal at 7p15.1 around the GWAS lead variant rs917275

**Supplementary Figure 46.** Regional association plot of previously unreported TAA signal at 2q13 around the GWAS lead variant rs17269661.

**Supplementary Figure 47.** Regional association plot of previously unreported TAA signal at 11q13.3 around the GWAS lead variant rs875106

**Supplementary Figure 48.** Regional association plot of previously unreported AAA signal at 3p14.1 around the GWAS lead variant rs9867045

**Supplementary Figure 49.** Regional association plot of previously unreported CAD signal at 17q22 around the GWAS lead variant rs2680688

**Supplementary Figure 50.** Regional association plot of previously unreported CAD signal at 20q11.23 around the GWAS lead variant rs2247054

**Supplementary Figure 51.** Regional association plot of previously unreported AAA signal at 1q41 around the GWAS lead variant rs4846769

**Supplementary Figure 52.** Regional association plot of previously unreported TAA signal at 3p21.31 around the GWAS lead variant rs62263038

**Supplementary Figure 53.** Regional association plot of previously unreported AAA signal at 4q31.22 around the GWAS lead variant rs77028772

**Supplementary Figure 54.** Regional association plot of previously unreported AAA signal at 7q36.1 around the GWAS lead variant rs3918226

**Supplementary Figure 55.** Regional association plot of previously unreported AAA signal at 4q32.1 around the GWAS lead variant rs12643599

**Supplementary Figure 56.** Regional association plot of previously unreported TAA signal at 15q15.1 around the GWAS lead variant rs17677757

**Supplementary Figure 57.** Regional association plot of previously unreported TAA signal at 2q33.2 around the GWAS lead variant rs6719001

**Supplementary Figure 58.** Regional association plot of previously unreported TAA signal at 8q23.1 around the GWAS lead variant rs4397378

**Supplementary Figure 59.** Regional association plot of previously unreported CAD signal at 3q21.3 around the GWAS lead variant rs62266959

**Supplementary Figure 60.** Regional association plot of previously unreported TAA signal at 19p13.3 around the GWAS lead variant rs4807216

**Supplementary Figure 61.** Regional association plot of previously unreported AAA signal at 1p32.2 around the GWAS lead variant rs72664332

**Supplementary Figure 62.** Regional association plot of previously unreported TAA signal at 13q12.2 around the GWAS lead variant rs9507870

**Supplementary Figure 63.** Regional association plot of previously unreported CAD signal at 17p13.1 around the GWAS lead variant rs7226020

**Supplementary Figure 64.** Regional association plot of previously unreported TAA signal at 6p21.2 around the GWAS lead variant rs3176334

**Supplementary Figure 65.** Regional association plot of previously unreported AAA signal at 6p21.31 around the GWAS lead variant rs75104038

**Supplementary Figure 66.** Regional association plot of previously unreported CAD signal at 16q22.2 around the GWAS lead variant rs145973320

**Supplementary Figure 67.** Regional association plot of previously unreported AAA signal at 6p24.1 around the GWAS lead variant rs9349379

**Supplementary Figure 68.** Regional association plot of previously unreported CAD signal at 7p15.3 around the GWAS lead variant rs13227860

**Supplementary Figure 69.** Regional association plot of previously unreported TAA signal at 21q22.2 around the GWAS lead variant rs77808533

**Supplementary Figure 70.** Regional association plot of previously unreported TAA signal at 5q23.1 around the GWAS lead variant rs77298376

**Supplementary Figure 71.** Regional association plot of previously unreported TAA signal at 12p12.2 around the GWAS lead variant rs4762746

**Supplementary Figure 72.** Regional association plot of previously unreported AAA signal at 11q21 around the GWAS lead variant rs1255179

**Supplementary Figure 73.** Regional association plot of previously unreported CAD signal at 14q24.2 around the GWAS lead variant rs2239222

**Supplementary Table 1**. Study population characteristics

**Supplementary Table 2.** Number of genetic variants in meta-analyses and multi-trait analyses

**Supplementary Table 3.** MTAG effective N calculations

**Supplementary Table 4a**. MTAG inflation estimation metrics

**Supplementary Table 4b**. MTAG maxFDR estimates

**Supplementary Table 5a.** CAD lead variants detected in MTAG meta-analysis

**Supplementary Table 5b**. CAD lead variants detected in MTAG CAD+AAA+TAA multi-trait meta-analysis

**Supplementary Table 5c.** CAD lead variants detected in MTAG CAD+AAA multi-trait meta-analysis

**Supplementary Table 5d.** CAD lead variants detected in MTAG CAD+TAA multi-trait meta-analysis

**Supplementary Table 6a**. AAA lead variants detected in MTAG meta-analysis

**Supplementary Table 6b.** AAA lead variants detected in MTAG CAD+AAA+TAA multi-trait meta-analysis

**Supplementary Table 6c.** AAA lead variants detected in MTAG CAD+AAA multi-trait meta-analysis

**Supplementary Table 6d**. AAA lead variants detected in MTAG AAA+TAA multi-trait meta-analysis

**Supplementary Table 7a**. TAA lead variants detected in MTAG meta-analysis

**Supplementary Table 7b**. TAA lead variants detected in MTAG CAD+AAA+TAA multi-trait meta-analysis

**Supplementary Table 7c.** TAA lead variants detected in MTAG CAD+TAA multi-trait meta-analysis

**Supplementary Table 7d.** TAA lead variants detected in MTAG AAA+TAA multi-trait meta-analysis

**Supplementary Table 8a.** Annotated novel CAD loci identified in MTAG meta-analysis

**Supplementary Table 8b.** Annotated novel AAA loci identified in MTAG meta-analysis

**Supplementary Table 8c.** Annotated novel TAA loci identified in MTAG meta-analysis

**Supplementary Table 8d.** Annotated novel CAD loci identified in MTAG CAD+AAA+TAA multi-trait meta-analysis

**Supplementary Table 8e.** Annotated novel CAD loci identified in MTAG CAD+AAA multi-trait meta-analysis

**Supplementary Table 8f.** Annotated novel CAD loci identified in MTAG CAD+TAA multi-trait meta-analysis

**Supplementary Table 8g.** Annotated novel AAA loci identified in MTAG CAD+AAA+TAA multi-trait meta-analysis

**Supplementary Table 8h.** Annotated novel AAA loci identified in MTAG CAD+AAA multi-trait meta-analysis

**Supplementary Table 8i.** Annotated novel AAA loci identified in MTAG AAA+TAA multi-trait meta-analysis

**Supplementary Table 8j.** Annotated novel TAA loci identified in MTAG CAD+AAA+TAA multi-trait meta-analysis

**Supplementary Table 8k.** Annotated novel TAA loci identified in MTAG CAD+TAA multi-trait meta-analysis

**Supplementary Table 8l.** Annotated novel TAA loci identified in MTAG AAA+TAA multi-trait meta-analysis

**Supplementary Table 9.** Pair-wise genetic correlations computed with LD score regression

**Supplementary Table 10.** Significant genome-wide genetic correlations observed for CAD

**Supplementary Table 11**. Significant genome-wide genetic correlations observed for AAA

**Supplementary Table 12**. Significant genome-wide genetic correlations observed for TAA

**Supplementary Table 13**. Significantly enriched MAGMA gene-sets and GO terms observed in CAD MTAG analyses

**Supplementary Table 14.** Significantly enriched MAGMA gene-sets and GO terms observed in AAA MTAG analyses

**Supplementary Table 15.** Significantly enriched MAGMA gene-sets and GO terms observed in TAA MTAG analyses

**Supplementary Table 16.** Stratified LD score regression cell type heritability enrichment

**Supplementary Table 17.** GWAS-eQTL colocalisation posterior probabilities

