## Supplementary Figures for "Genome-wide Cross-multi-trait Analysis into Shared Genetic Architecture of Coronary Artery Disease and Aortic Aneurysms"

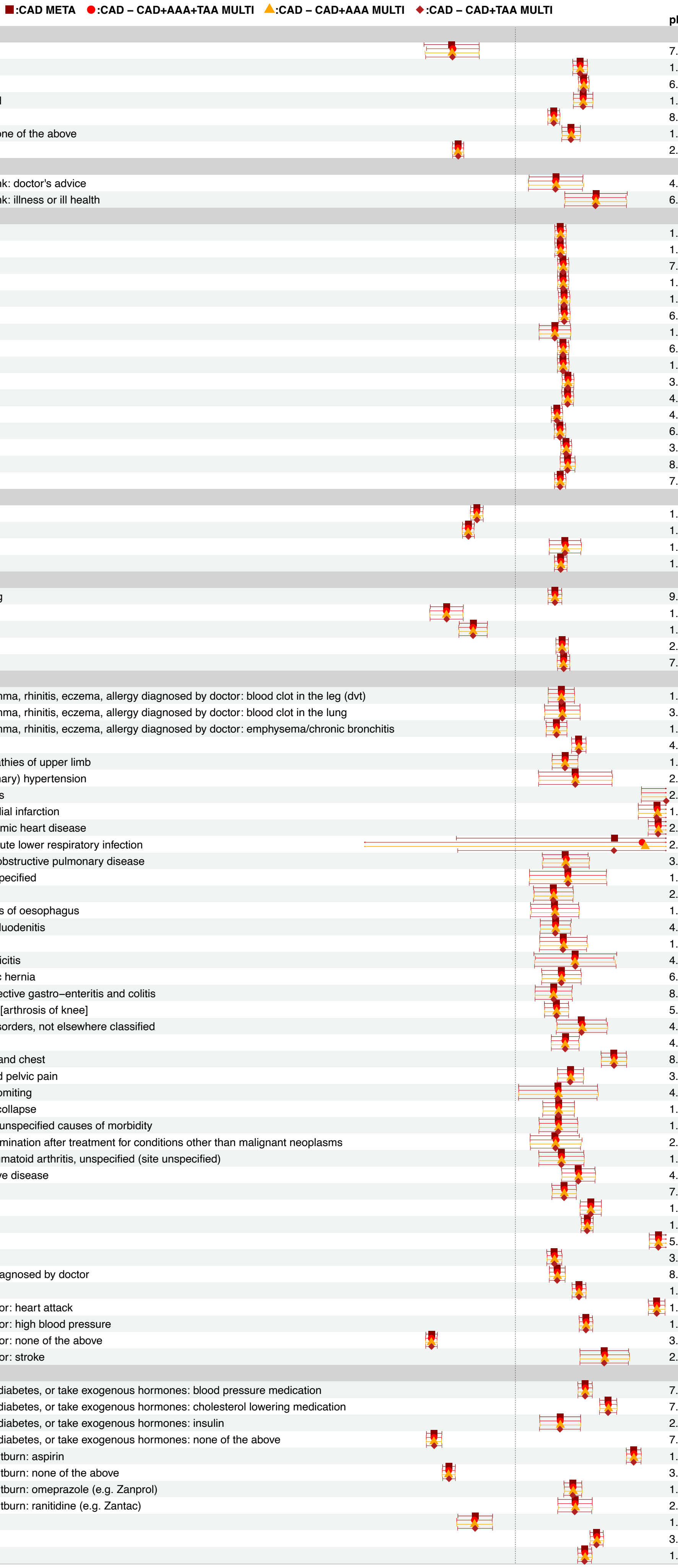


| Trait | $pFDR_{min}$ | $pFDR_{max}$ |
|---|---|---|
| **Activity, Fitness and Sleep** | | |
| Duration of fitness test | 7.04e−06 | 1.13e−05 |
| Health satisfaction | 1.57e−67 | 3.16e−66 |
| Overall health rating | 6.95e−143 | 2.91e−139 |
| Shortness of breath walking on level ground | 1.05e−41 | 2.26e−41 |
| Time spent watching television (tv) | 8.48e−36 | 3.23e−35 |
| Types of physical activity in last 4 weeks: none of the above | 1.26e−31 | 4.11e−31 |
| Usual walking pace | 2.02e−89 | 1.61e−87 |
| **Alcohol** | | |
| Reason for reducing amount of alcohol drunk: doctor's advice | 4.71e−03 | 5.68e−03 |
| Reason for reducing amount of alcohol drunk: illness or ill health | 6.12e−07 | 8.98e−07 |
| **Anthropometry** | | |
| Arm fat mass (left) | 1.03e−54 | 2.56e−54 |
| Arm fat mass (right) | 1.81e−55 | 1.31e−54 |
| Arm fat percentage (left) | 7.95e−64 | 3.86e−63 |
| Arm fat percentage (right) | 1.80e−63 | 4.64e−63 |
| Body fat percentage | 1.62e−63 | 3.21e−63 |
| Body mass index (bmi) | 6.07e−66 | 6.27e−65 |
| Extreme waist−to−hip ratio | 1.54e−06 | 2.19e−06 |
| Leg fat mass (left) | 6.06e−60 | 2.45e−59 |
| Leg fat mass (right) | 1.41e−59 | 6.94e−59 |
| Leg fat percentage (left) | 3.39e−68 | 1.02e−67 |
| Leg fat percentage (right) | 4.28e−69 | 1.52e−68 |
| Trunk fat mass | 4.87e−47 | 1.65e−46 |
| Trunk fat percentage | 6.11e−52 | 1.27e−51 |
| Waist circumference | 3.87e−75 | 3.00e−74 |
| Waist−to−hip ratio | 8.34e−42 | 2.35e−41 |
| Whole body fat mass | 7.78e−55 | 3.23e−54 |
| **Biomarkers** | | |
| Apolipoprotein a−i | 1.15e−32 | 1.88e−32 |
| Hdl cholesterol | 1.53e−54 | 5.41e−53 |
| Homa−ir | 1.92e−09 | 2.21e−09 |
| Triglycerides | 1.14e−44 | 9.04e−44 |
| **Cardiovascular** | | |
| Diastolic blood pressure, automated reading | 9.77e−29 | 2.11e−28 |
| Maximum workload during fitness test | 1.39e−15 | 1.96e−15 |
| Pulse wave peak to peak time | 1.11e−08 | 1.30e−08 |
| Systolic blood pressure | 2.19e−48 | 5.95e−48 |
| Systolic blood pressure, automated reading | 7.06e−52 | 2.10e−49 |
| **Diseases** | | |
| Blood clot, dvt, bronchitis, emphysema, asthma, rhinitis, eczema, allergy diagnosed by doctor: blood clot in the leg (dvt) | 1.48e−11 | 2.45e−11 |
| Blood clot, dvt, bronchitis, emphysema, asthma, rhinitis, eczema, allergy diagnosed by doctor: blood clot in the lung | 3.50e−07 | 5.75e−07 |
| Blood clot, dvt, bronchitis, emphysema, asthma, rhinitis, eczema, allergy diagnosed by doctor: emphysema/chronic bronchitis | 1.88e−14 | 4.62e−14 |
| Diabetes diagnosed by doctor | 4.64e−66 | 2.45e−65 |
| Diagnoses − main icd10: g56 mononeuropathies of upper limb | 1.42e−13 | 1.98e−13 |
| Diagnoses − main icd10: i10 essential (primary) hypertension | 2.01e−03 | 2.25e−03 |
| Diagnoses − main icd10: i20 angina pectoris | 2.01e−31 | 1.53e−30 |
| Diagnoses − main icd10: i21 acute myocardial infarction | 1.59e−50 | 4.55e−47 |
| Diagnoses − main icd10: i25 chronic ischaemic heart disease | 2.39e−171 | 1.16e−169 |
| Diagnoses − main icd10: j22 unspecified acute lower respiratory infection | 2.58e−01 | 4.51e−01 |
| Diagnoses − main icd10: j44 other chronic obstructive pulmonary disease | 3.21e−05 | 4.86e−05 |
| Diagnoses − main icd10: j45.9 asthma, unspecified | 1.04e−02 | 1.14e−02 |
| Diagnoses − main icd10: k20 oesophagitis | 2.32e−04 | 2.94e−04 |
| Diagnoses − main icd10: k22 other diseases of oesophagus | 1.76e−03 | 2.37e−03 |
| Diagnoses − main icd10: k29 gastritis and duodenitis | 4.43e−07 | 6.75e−07 |
| Diagnoses − main icd10: k30 dyspepsia | 1.20e−04 | 1.35e−04 |
| Diagnoses − main icd10: k35 acute appendicitis | 4.73e−03 | 6.78e−03 |
| Diagnoses − main icd10: k44 diaphragmatic hernia | 6.92e−06 | 1.03e−05 |
| Diagnoses − main icd10: k52 other non−infective gastro−enteritis and colitis | 8.14e−05 | 1.00e−04 |
| Diagnoses − main icd10: m17 gonarthrosis [arthrosis of knee] | 5.65e−11 | 6.63e−11 |
| Diagnoses − main icd10: m25 other joint disorders, not elsewhere classified | 4.39e−07 | 6.46e−07 |
| Diagnoses − main icd10: m54 dorsalgia | 4.66e−12 | 7.96e−12 |
| Diagnoses − main icd10: r07 pain in throat and chest | 8.77e−52 | 5.01e−51 |
| Diagnoses − main icd10: r10 abdominal and pelvic pain | 3.39e−16 | 9.56e−16 |
| Diagnoses − main icd10: r11 nausea and vomiting | 4.50e−02 | 4.59e−02 |
| Diagnoses − main icd10: r55 syncope and collapse | 1.57e−07 | 1.98e−07 |
| Diagnoses − main icd10: r69 unknown and unspecified causes of morbidity | 1.99e−05 | 2.63e−05 |
| Diagnoses − main icd10: z09 follow−up examination after treatment for conditions other than malignant neoplasms | 2.62e−03 | 3.33e−03 |
| Diagnoses − secondary icd10: m06.99 rheumatoid arthritis, unspecified (site unspecified) | 1.09e−04 | 1.16e−04 |
| Eye problems/disorders: diabetes related eye disease | 4.90e−13 | 6.10e−13 |
| Had major operations | 7.92e−15 | 2.12e−14 |
| Ischemic stroke | 1.03e−43 | 1.96e−42 |
| Long−standing illness, disability or infirmity | 1.14e−121 | 1.07e−118 |
| Major coronary heart disease event | 5.45e−206 | 3.74e−204 |
| Number of operations, self−reported | 3.25e−24 | 1.19e−23 |
| Other serious medical condition/disability diagnosed by doctor | 8.89e−25 | 2.57e−24 |
| Type 2 diabetes | 1.98e−71 | 4.38e−71 |
| Vascular/heart problems diagnosed by doctor: heart attack | 1.17e−247 | 8.35e−238 |
| Vascular/heart problems diagnosed by doctor: high blood pressure | 1.09e−94 | 9.20e−93 |
| Vascular/heart problems diagnosed by doctor: none of the above | 3.60e−179 | 1.79e−174 |
| Vascular/heart problems diagnosed by doctor: stroke | 2.29e−12 | 4.31e−12 |
| **Drugs and Supplements** | | |
| Medication for cholesterol, blood pressure, diabetes, or take exogenous hormones: blood pressure medication | 7.81e−79 | 3.90e−78 |
| Medication for cholesterol, blood pressure, diabetes, or take exogenous hormones: cholesterol lowering medication | 7.32e−98 | 2.12e−97 |
| Medication for cholesterol, blood pressure, diabetes, or take exogenous hormones: insulin | 2.99e−05 | 3.68e−05 |
| Medication for cholesterol, blood pressure, diabetes, or take exogenous hormones: none of the above | 7.55e−96 | 5.76e−92 |
| Medication for pain relief, constipation, heartburn: aspirin | 1.07e−216 | 5.74e−209 |
| Medication for pain relief, constipation, heartburn: none of the above | 3.11e−91 | 5.30e−90 |
| Medication for pain relief, constipation, heartburn: omeprazole (e.g. Zanprol) | 1.15e−36 | 4.72e−35 |
| Medication for pain relief, constipation, heartburn: ranitidine (e.g. Zantac) | 2.11e−11 | 2.85e−11 |
| Mineral and other dietary supplements: iron | 1.14e−05 | 1.46e−05 |
| Number of treatments/medications taken | 3.83e−126 | 3.26e−124 |
| Taking other prescription medications | 1.15e−83 | 3.12e−82 |

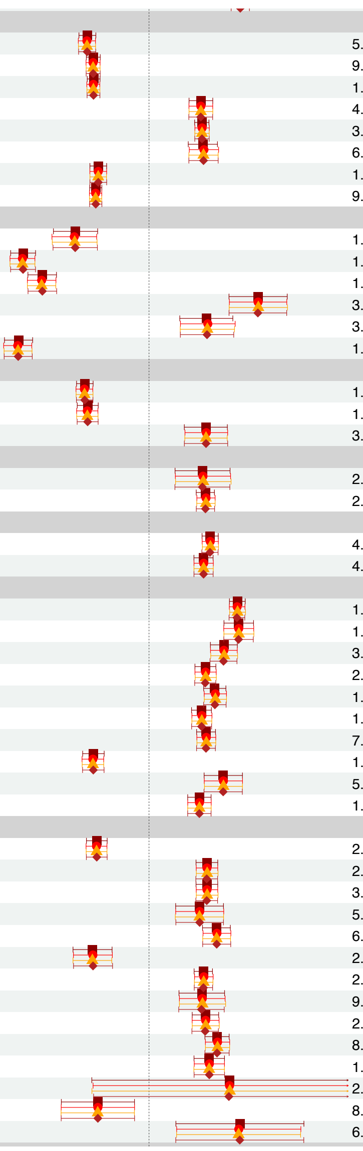

| | | |
|---|---|---|
| **Education and Intelligence** | | |
| Age completed full time education | 5.09e−45 | 2.17e−44 |
| Qualifications: a levels/as levels or equivalent | 9.88e−55 | 1.02e−53 |
| Qualifications: college or university degree | 1.70e−63 | 5.45e−62 |
| Qualifications: cses or equivalent | 4.83e−17 | 8.05e−17 |
| Qualifications: none of the above | 3.09e−46 | 4.83e−46 |
| Qualifications: nvq or hnd or hnc or equivalent | 6.85e−13 | 1.64e−12 |
| Qualifications: o levels/gcses or equivalent | 1.94e−32 | 1.67e−31 |
| Years of schooling | 9.06e−65 | 5.00e−63 |
| **Family Member Health** | | |
| Father still alive | 1.54e−10 | 2.14e−10 |
| Father's age at death | 1.10e−86 | 7.79e−86 |
| Mother's age at death | 1.80e−48 | 5.63e−47 |
| Non−accidental death in close genetic family | 3.26e−13 | 7.07e−13 |
| Number of older siblings | 3.83e−05 | 4.66e−05 |
| Parental longevity (combined parental age at death) | 1.13e−73 | 4.67e−72 |
| **Gynecology and Fertility** | | |
| Age at first live birth | 1.91e−51 | 2.10e−49 |
| Age at last live birth | 1.74e−29 | 7.83e−29 |
| Diagnoses – main icd10: n92 excessive, frequent and irregular menstruation | 3.02e−07 | 5.44e−07 |
| **Lung Function** | | |
| Childhood asthma (age<16) | 2.17e−04 | 2.40e−04 |
| Wheeze or whistling in the chest in last year | 2.42e−33 | 1.36e−32 |
| **Mood and Behaviour** | | |
| Frequency of tiredness / lethargy in last 2 weeks | 4.23e−49 | 9.37e−48 |
| Frequency of unenthusiasm / disinterest in last 2 weeks | 4.59e−28 | 1.17e−27 |
| **Pain and Skeletal** | | |
| Chest pain or discomfort | 1.38e−106 | 8.33e−10[illegible] |
| Chest pain or discomfort walking normally | 1.26e−31 | 8.49e−31 |
| Leg pain on walking | 3.93e−27 | 1.71e−26 |
| Pain type(s) experienced in last month: back pain | 2.88e−25 | 1.15e−24 |
| Pain type(s) experienced in last month: hip pain | 1.04e−30 | 1.11e−29 |
| Pain type(s) experienced in last month: knee pain | 1.92e−24 | 3.35e−24 |
| Pain type(s) experienced in last month: neck or shoulder pain | 7.58e−32 | 2.91e−31 |
| Pain type(s) experienced in last month: none of the above | 1.25e−23 | 2.51e−23 |
| Pain type(s) experienced in last month: pain all over the body | 5.72e−14 | 1.19e−13 |
| Pain type(s) experienced in last month: stomach or abdominal pain | 1.33e−16 | 1.71e−16 |
| **Smoking** | | |
| Age of smoking initiation | 2.63e−22 | 3.99e−22 |
| Cigarettes per day | 2.99e−25 | 1.53e−24 |
| Cigarettes smoked per day | 3.18e−25 | 1.73e−24 |
| Ever stopped smoking for 6+ months | 5.61e−05 | 7.69e−05 |
| Exposure to tobacco smoke at home | 6.05e−21 | 1.72e−20 |
| Former vs current smoker | 2.45e−08 | 4.38e−08 |
| Maternal smoking around birth | 2.99e−30 | 5.97e−30 |
| Number of cigarettes currently smoked daily (current cigarette smokers) | 9.70e−06 | 1.22e−05 |
| Number of cigarettes previously smoked daily | 2.78e−16 | 8.33e−16 |
| Pack years of smoking | 8.16e−28 | 2.64e−27 |
| Smoking/smokers in household | 1.92e−14 | 4.34e−14 |
| Why reduced smoking: doctor's advice | 2.90e−01 | 2.95e−01 |
| Why reduced smoking: health precaution | 8.98e−03 | 9.26e−03 |
| Why reduced smoking: illness or ill health | 6.87e−03 | 8.09e−03 |

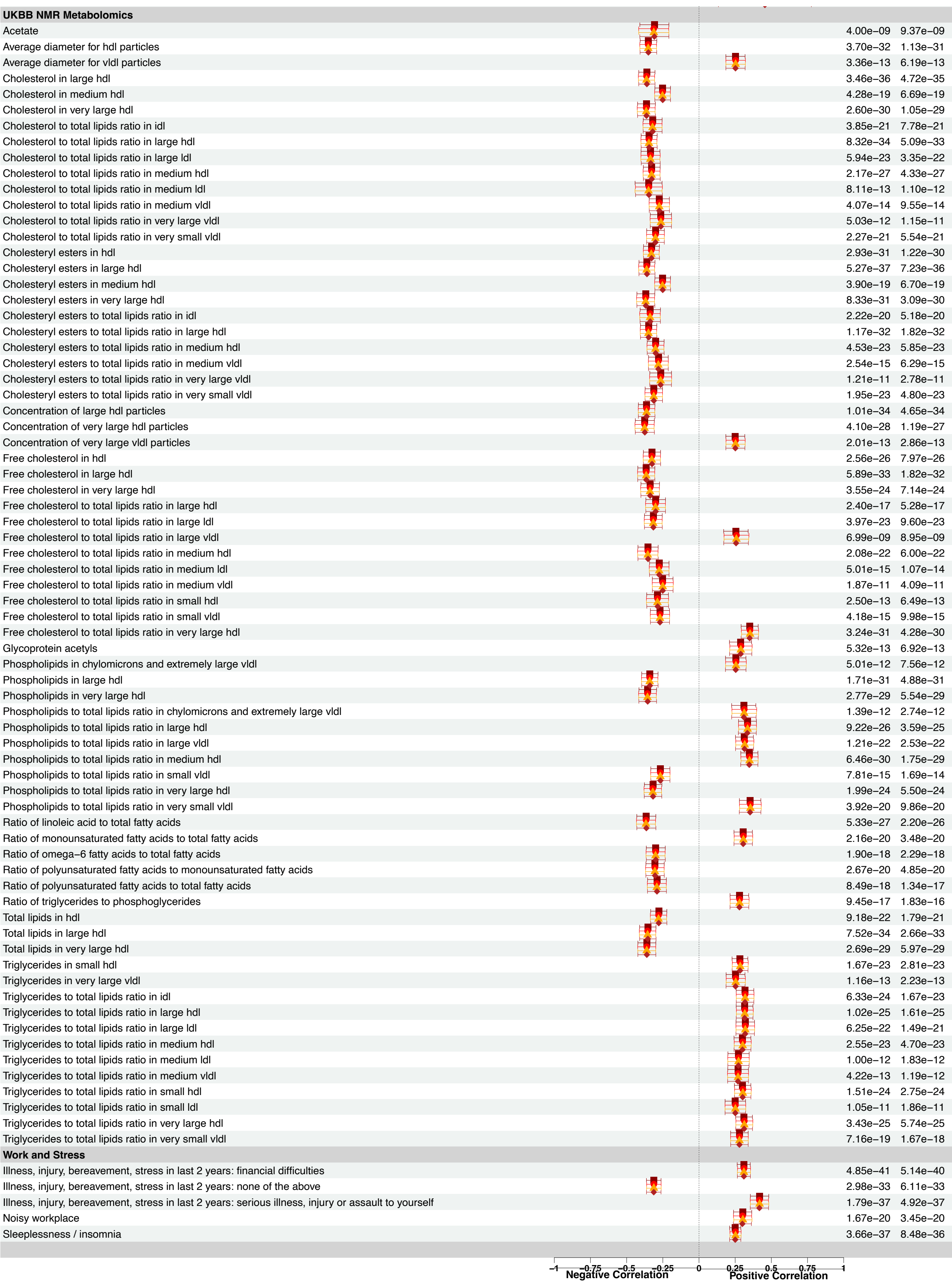


Figure 1. **Summary of genome-wide genetic correlation patterns for CAD.** Forest plot of genetic correlations computed between TAA and other complex traits with LDSC. Correlated traits are shown on the left column, divided to related categories. Genetic correlation ($r_g$) and the associated 95% confidence intervals are presented in the center panel. Trait

configurations indicated by coloured symbols. The maximum and minimum FDR-adjusted P values for each correlation across different trait configurations are listed on right-side columns. Only the strongest ($-0.25 > r_g > 0.25$) are shown on the figure. AAA, abdominal aortic aneurysm; CAD, coronary artery disease; FDR, false discovery rate; LDSC, linkage disequilibrium score regression, META, meta-analysis; MULTI, multi-trait analysis; $r_g$, genetic correlation; TAA, thoracic aortic aneurysm.

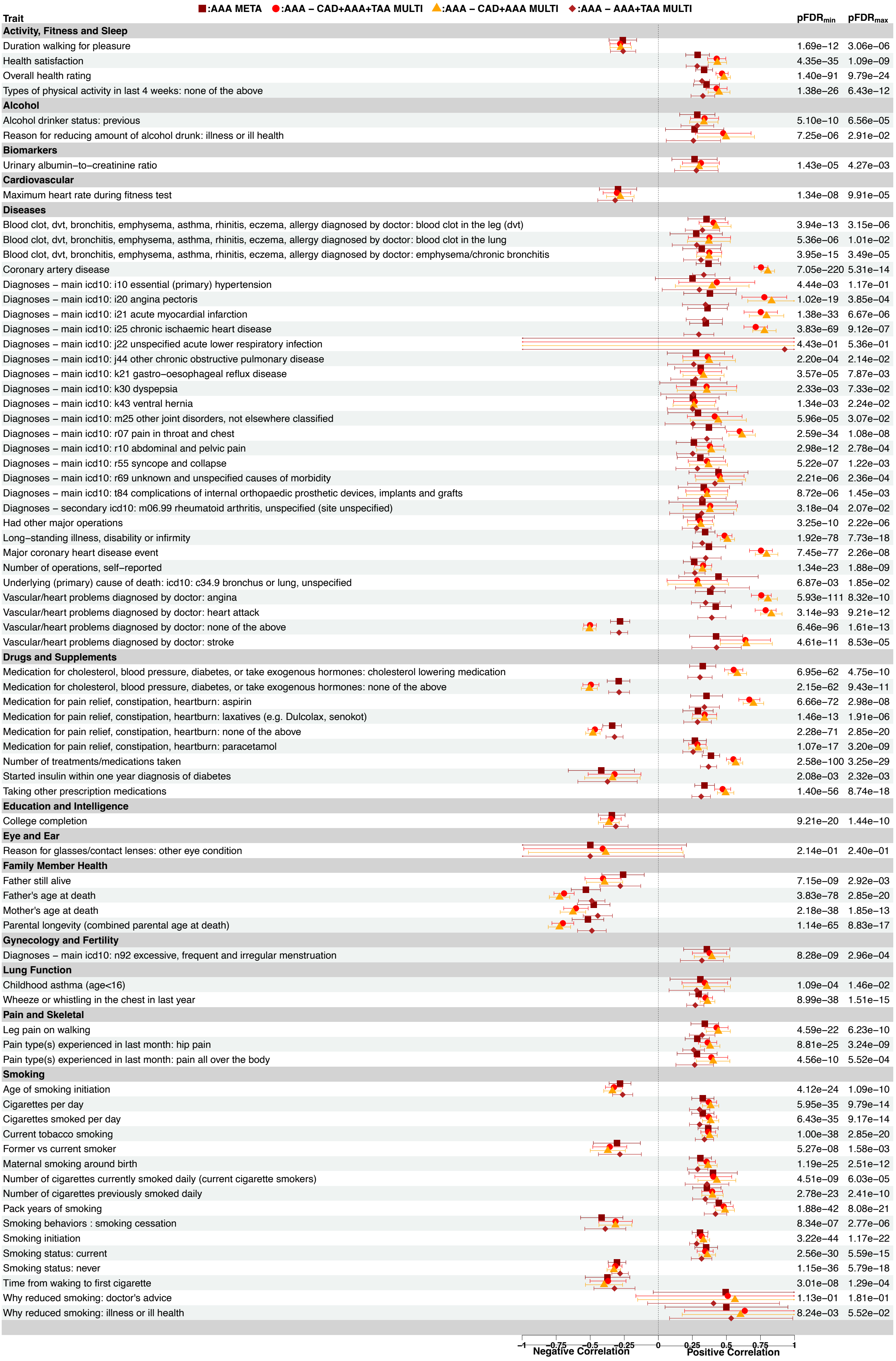
■:AAA META ●:AAA – CAD+AAA+TAA MULTI ▲:AAA – CAD+AAA MULTI ◆:AAA – AAA+TAA MULTI
Trait
pFDRmin pFDRmax
Activity, Fitness and Sleep
Duration walking for pleasure 1.69e−12 3.06e−06
Health satisfaction 4.35e−35 1.09e−09
Overall health rating 1.40e−91 9.79e−24
Types of physical activity in last 4 weeks: none of the above 1.38e−26 6.43e−12
Alcohol
Alcohol drinker status: previous 5.10e−10 6.56e−05
Reason for reducing amount of alcohol drunk: illness or ill health 7.25e−06 2.91e−02
Biomarkers
Urinary albumin−to−creatinine ratio 1.43e−05 4.27e−03
Cardiovascular
Maximum heart rate during fitness test 1.34e−08 9.91e−05
Diseases
Blood clot, dvt, bronchitis, emphysema, asthma, rhinitis, eczema, allergy diagnosed by doctor: blood clot in the leg (dvt) 3.94e−13 3.15e−06
Blood clot, dvt, bronchitis, emphysema, asthma, rhinitis, eczema, allergy diagnosed by doctor: blood clot in the lung 5.36e−06 1.01e−02
Blood clot, dvt, bronchitis, emphysema, asthma, rhinitis, eczema, allergy diagnosed by doctor: emphysema/chronic bronchitis 3.95e−15 3.49e−05
Coronary artery disease 7.05e−220 5.31e−14
Diagnoses − main icd10: i10 essential (primary) hypertension 4.44e−03 1.17e−01
Diagnoses − main icd10: i20 angina pectoris 1.02e−19 3.85e−04
Diagnoses − main icd10: i21 acute myocardial infarction 1.38e−33 6.67e−06
Diagnoses − main icd10: i25 chronic ischaemic heart disease 3.83e−69 9.12e−07
Diagnoses − main icd10: j22 unspecified acute lower respiratory infection 4.43e−01 5.36e−01
Diagnoses − main icd10: j44 other chronic obstructive pulmonary disease 2.20e−04 2.14e−02
Diagnoses − main icd10: k21 gastro−oesophageal reflux disease 3.57e−05 7.87e−03
Diagnoses − main icd10: k30 dyspepsia 2.33e−03 7.33e−02
Diagnoses − main icd10: k43 ventral hernia 1.34e−03 2.24e−02
Diagnoses − main icd10: m25 other joint disorders, not elsewhere classified 5.96e−05 3.07e−02
Diagnoses − main icd10: r07 pain in throat and chest 2.59e−34 1.08e−08
Diagnoses − main icd10: r10 abdominal and pelvic pain 2.98e−12 2.78e−04
Diagnoses − main icd10: r55 syncope and collapse 5.22e−07 1.22e−03
Diagnoses − main icd10: r69 unknown and unspecified causes of morbidity 2.21e−06 2.36e−04
Diagnoses − main icd10: t84 complications of internal orthopaedic prosthetic devices, implants and grafts 8.72e−06 1.45e−03
Diagnoses − secondary icd10: m06.99 rheumatoid arthritis, unspecified (site unspecified) 3.18e−04 2.07e−02
Had other major operations 3.25e−10 2.22e−06
Long−standing illness, disability or infirmity 1.92e−78 7.73e−18
Major coronary heart disease event 7.45e−77 2.26e−08
Number of operations, self−reported 1.34e−23 1.88e−09
Underlying (primary) cause of death: icd10: c34.9 bronchus or lung, unspecified 6.87e−03 1.85e−02
Vascular/heart problems diagnosed by doctor: angina 5.93e−111 8.32e−10
Vascular/heart problems diagnosed by doctor: heart attack 3.14e−93 9.21e−12
Vascular/heart problems diagnosed by doctor: none of the above 6.46e−96 1.61e−13
Vascular/heart problems diagnosed by doctor: stroke 4.61e−11 8.53e−05
Drugs and Supplements
Medication for cholesterol, blood pressure, diabetes, or take exogenous hormones: cholesterol lowering medication 6.95e−62 4.75e−10
Medication for cholesterol, blood pressure, diabetes, or take exogenous hormones: none of the above 2.15e−62 9.43e−11
Medication for pain relief, constipation, heartburn: aspirin 6.66e−72 2.98e−08
Medication for pain relief, constipation, heartburn: laxatives (e.g. Dulcolax, senokot) 1.46e−13 1.91e−06
Medication for pain relief, constipation, heartburn: none of the above 2.28e−71 2.85e−20
Medication for pain relief, constipation, heartburn: paracetamol 1.07e−17 3.20e−09
Number of treatments/medications taken 2.58e−100 3.25e−29
Started insulin within one year diagnosis of diabetes 2.08e−03 2.32e−03
Taking other prescription medications 1.40e−56 8.74e−18
Education and Intelligence
College completion 9.21e−20 1.44e−10
Eye and Ear
Reason for glasses/contact lenses: other eye condition 2.14e−01 2.40e−01
Family Member Health
Father still alive 7.15e−09 2.92e−03
Father's age at death 3.83e−78 2.85e−20
Mother's age at death 2.18e−38 1.85e−13
Parental longevity (combined parental age at death) 1.14e−65 8.83e−17
Gynecology and Fertility
Diagnoses − main icd10: n92 excessive, frequent and irregular menstruation 8.28e−09 2.96e−04
Lung Function
Childhood asthma (age<16) 1.09e−04 1.46e−02
Wheeze or whistling in the chest in last year 8.99e−38 1.51e−15
Pain and Skeletal
Leg pain on walking 4.59e−22 6.23e−10
Pain type(s) experienced in last month: hip pain 8.81e−25 3.24e−09
Pain type(s) experienced in last month: pain all over the body 4.56e−10 5.52e−04
Smoking
Age of smoking initiation 4.12e−24 1.09e−10
Cigarettes per day 5.95e−35 9.79e−14
Cigarettes smoked per day 6.43e−35 9.17e−14
Current tobacco smoking 1.00e−38 2.85e−20
Former vs current smoker 5.27e−08 1.58e−03
Maternal smoking around birth 1.19e−25 2.51e−12
Number of cigarettes currently smoked daily (current cigarette smokers) 4.51e−09 6.03e−05
Number of cigarettes previously smoked daily 2.78e−23 2.41e−10
Pack years of smoking 1.88e−42 8.08e−21
Smoking behaviors : smoking cessation 8.34e−07 2.77e−06
Smoking initiation 3.22e−44 1.17e−22
Smoking status: current 2.56e−30 5.59e−15
Smoking status: never 1.15e−36 5.79e−18
Time from waking to first cigarette 3.01e−08 1.29e−04
Why reduced smoking: doctor's advice 1.13e−01 1.81e−01
Why reduced smoking: illness or ill health 8.24e−03 5.52e−02
−1 −0.75 −0.5 −0.25 0 0.25 0.5 0.75 1
Negative Correlation
Positive Correlation

Figure 2. **Summary of genome-wide genetic correlation patterns for AAA.** Forest plot of genetic correlations computed between TAA and other complex traits with LDSC. Correlated traits are shown on the left column, divided to related categories. Genetic correlation ($r_g$) and the associated 95% confidence intervals are presented in the center panel. Trait configurations indicated by coloured symbols. The maximum and minimum FDR-adjusted P values for each correlation across different trait configurations are listed on right-side columns. Only the strongest ($-0.25 > r_g > 0.25$) are shown on the figure. AAA, abdominal aortic aneurysm; CAD, coronary artery disease; FDR, false discovery rate; LDSC, linkage disequilibrium score regression, META, meta-analysis; MULTI, multi-trait analysis; $r_g$, genetic correlation; TAA, thoracic aortic aneurysm.

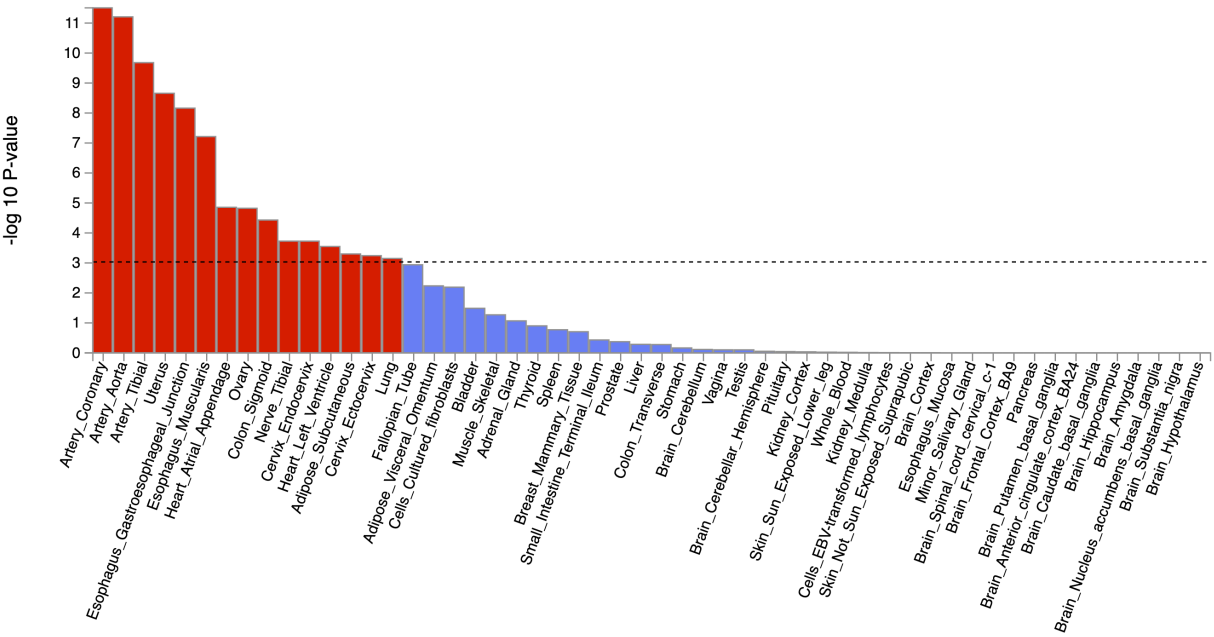


Figure 1. **Tissue-specific gene expression profile enrichment of CAD in MTAG meta-analysis**. Enrichment estimated with MAGMA version 1.08 gene-property analysis as implemented in FUMA version 1.3.4. Tested tissue gene expression profiles are listed on the X-axis. Strength of the association is depicted on the Y-axis as $-\log_{10}(P)$.

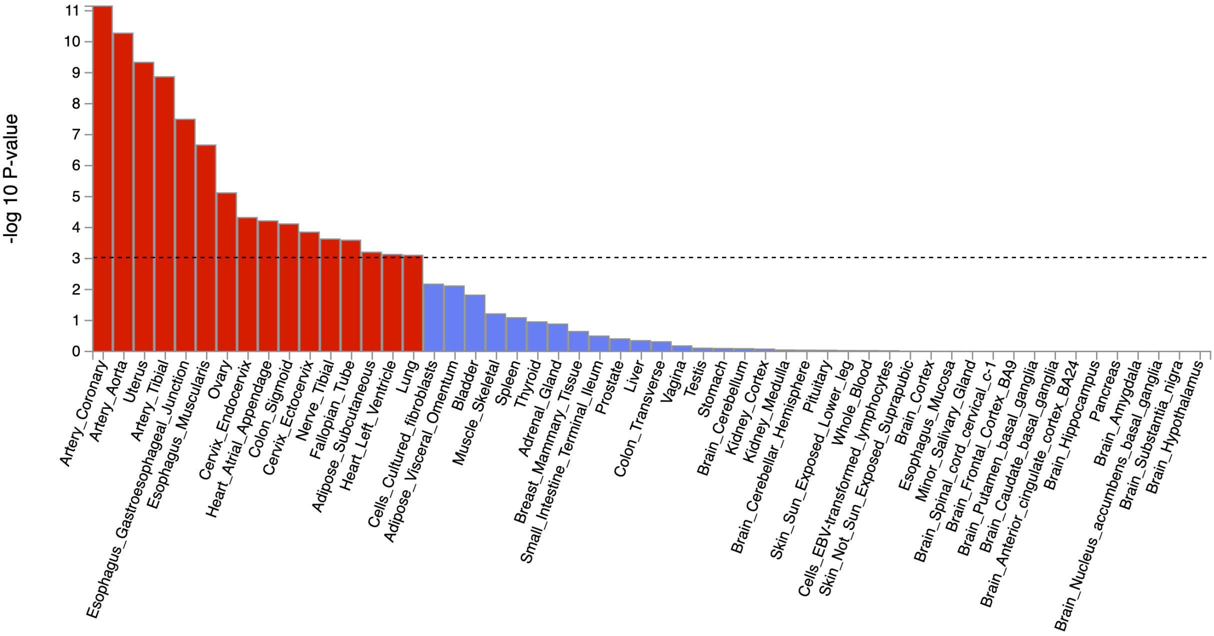


Figure 4. **Tissue-specific gene expression profile enrichment of CAD in MTAG CAD+AAA+TAA multi-trait meta-analysis**. Enrichment estimated with MAGMA version 1.08 gene-property analysis as implemented in FUMA version 1.3.4. Tested tissue gene expression profiles are listed on the X-axis. Strength of the association is depicted on the Y-axis as $-\log_{10}(P)$.

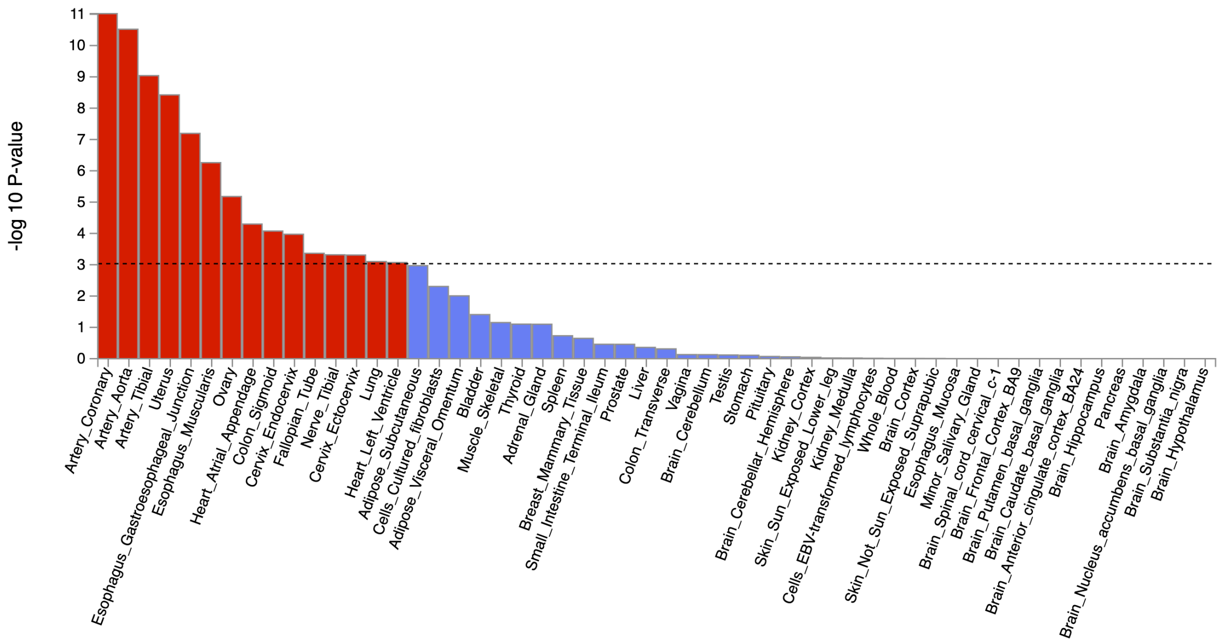


Figure 5. **Tissue-specific gene expression profile enrichment of CAD in MTAG CAD+AAA multi-trait meta-analysis**. Enrichment estimated with MAGMA version 1.08 gene-property analysis as implemented in FUMA version 1.3.4. Tested tissue gene expression profiles are listed on the X-axis. Strength of the association is depicted on the Y-axis as -$\log_{10}(P)$.

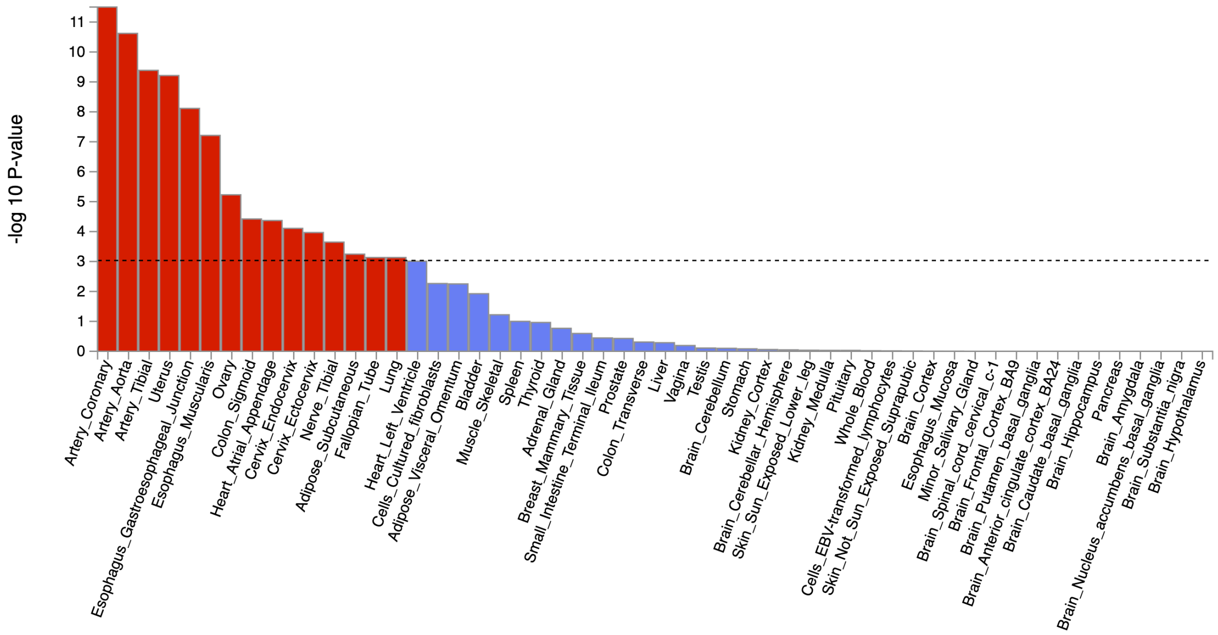


Figure 6. **Tissue-specific gene expression profile enrichment of CAD in MTAG CAD+TAA multi-trait meta-analysis**. Enrichment estimated with MAGMA version 1.08 gene-property analysis as implemented in FUMA version 1.3.4. Tested tissue gene expression profiles are listed on the X-axis. Strength of the association is depicted on the Y-axis as -$\log_{10}(P)$.

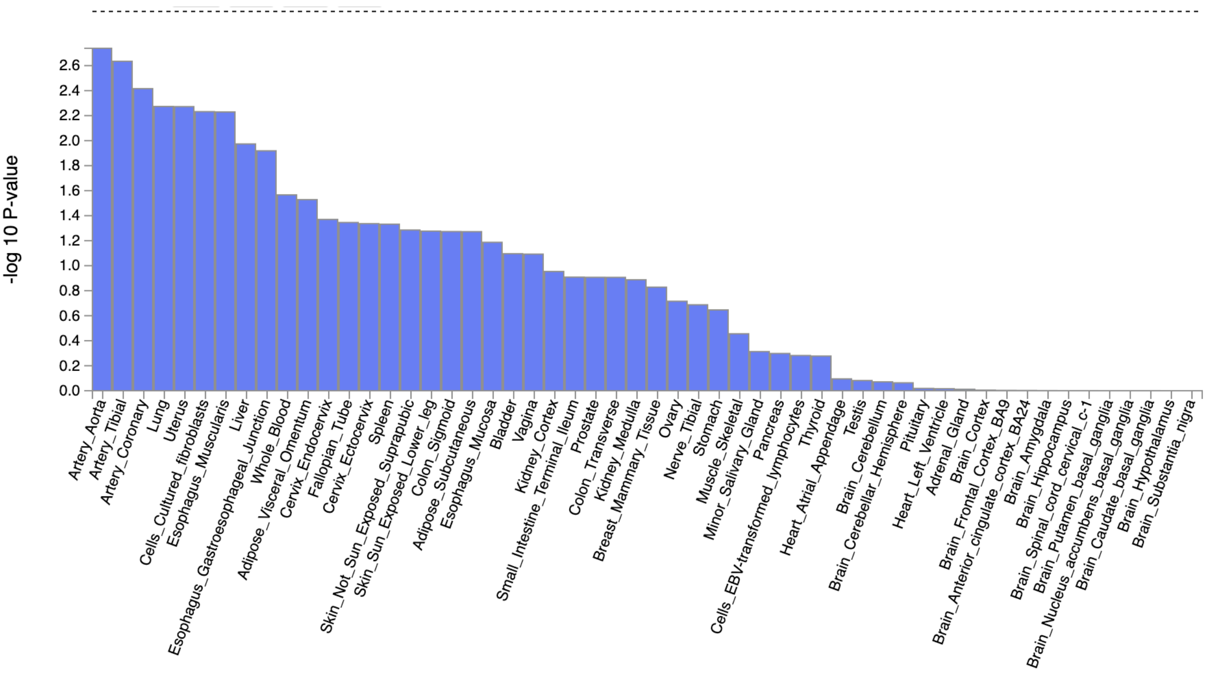


Figure 7. **Tissue-specific gene expression profile enrichment of AAA in MTAG meta-analysis**. Enrichment estimated with MAGMA version 1.08 gene-property analysis as implemented in FUMA version 1.3.4. Tested tissue gene expression profiles are listed on the X-axis. Strength of the association is depicted on the Y-axis as -log$_{10}$($P$).

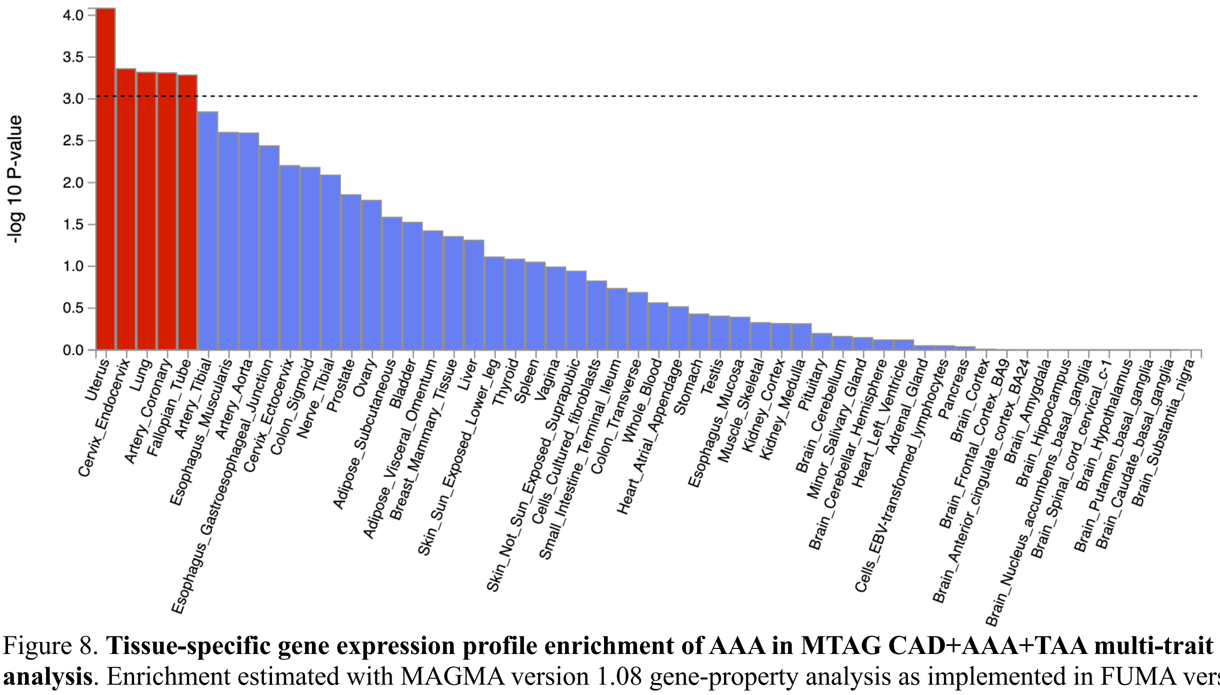


Figure 8. **Tissue-specific gene expression profile enrichment of AAA in MTAG CAD+AAA+TAA multi-trait meta-analysis**. Enrichment estimated with MAGMA version 1.08 gene-property analysis as implemented in FUMA version 1.3.4. Tested tissue gene expression profiles are listed on the X-axis. Strength of the association is depicted on the Y-axis as -log$_{10}$($P$).

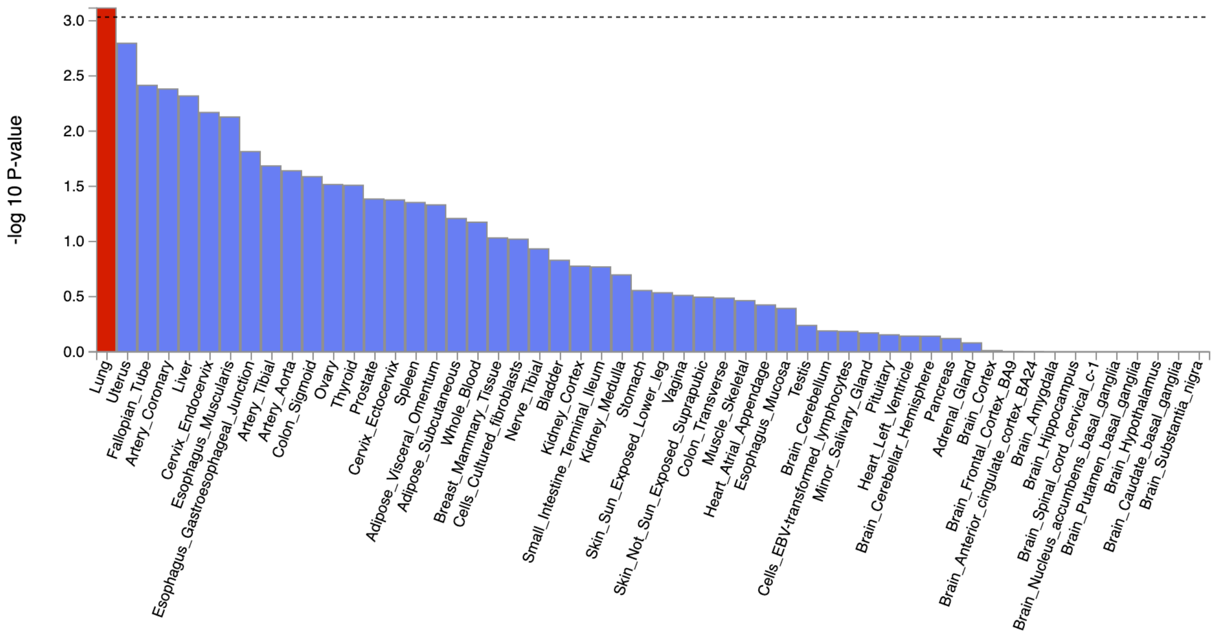


Figure 9. **Tissue-specific gene expression profile enrichment of AAA in MTAG CAD+AAA multi-trait meta-analysis**. Enrichment estimated with MAGMA version 1.08 gene-property analysis as implemented in FUMA version 1.3.4. Tested tissue gene expression profiles are listed on the X-axis. Strength of the association is depicted on the Y-axis as -$\log_{10}(P)$.

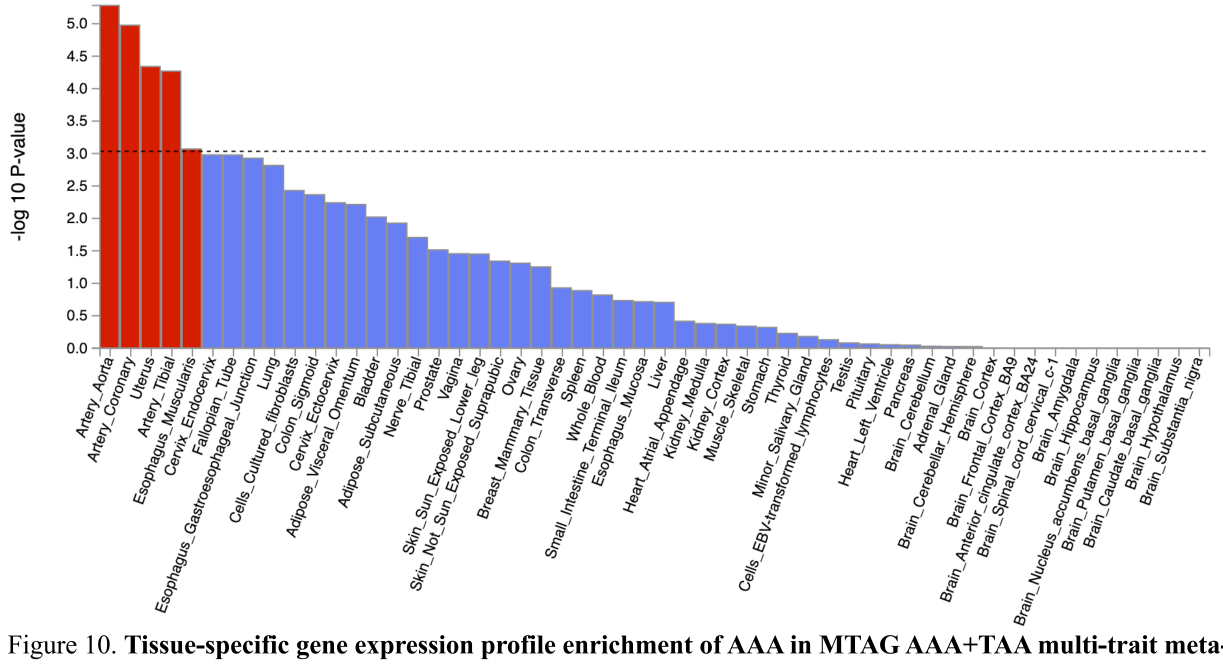


Figure 10. **Tissue-specific gene expression profile enrichment of AAA in MTAG AAA+TAA multi-trait meta-analysis**. Enrichment estimated with MAGMA version 1.08 gene-property analysis as implemented in FUMA version 1.3.4. Tested tissue gene expression profiles are listed on the X-axis. Strength of the association is depicted on the Y-axis as -$\log_{10}(P)$.

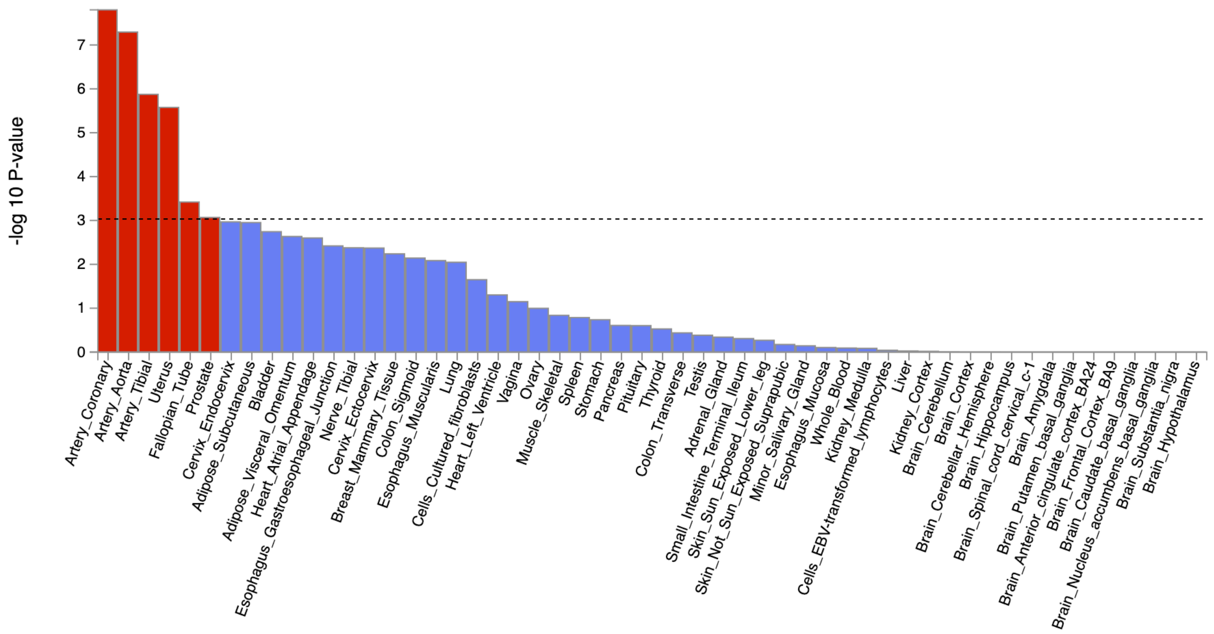


Figure 11. **Tissue-specific gene expression profile enrichment of TAA in MTAG meta-analysis**. Enrichment estimated with MAGMA version 1.08 gene-property analysis as implemented in FUMA version 1.3.4. Tested tissue gene expression profiles are listed on the X-axis. Strength of the association is depicted on the Y-axis as $-\log_{10}(P)$.

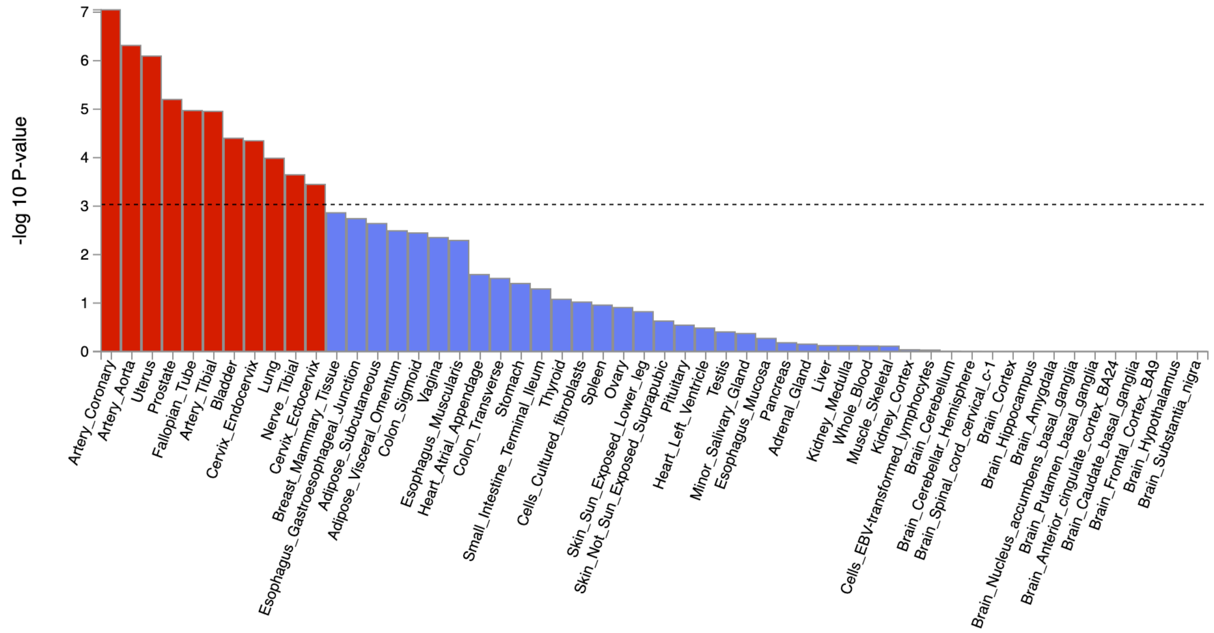


Figure 12. **Tissue-specific gene expression profile enrichment of TAA in MTAG CAD+AAA+TAA multi-trait meta-analysis**. Enrichment estimated with MAGMA version 1.08 gene-property analysis as implemented in FUMA version 1.3.4. Tested tissue gene expression profiles are listed on the X-axis. Strength of the association is depicted on the Y-axis as $-\log_{10}(P)$.

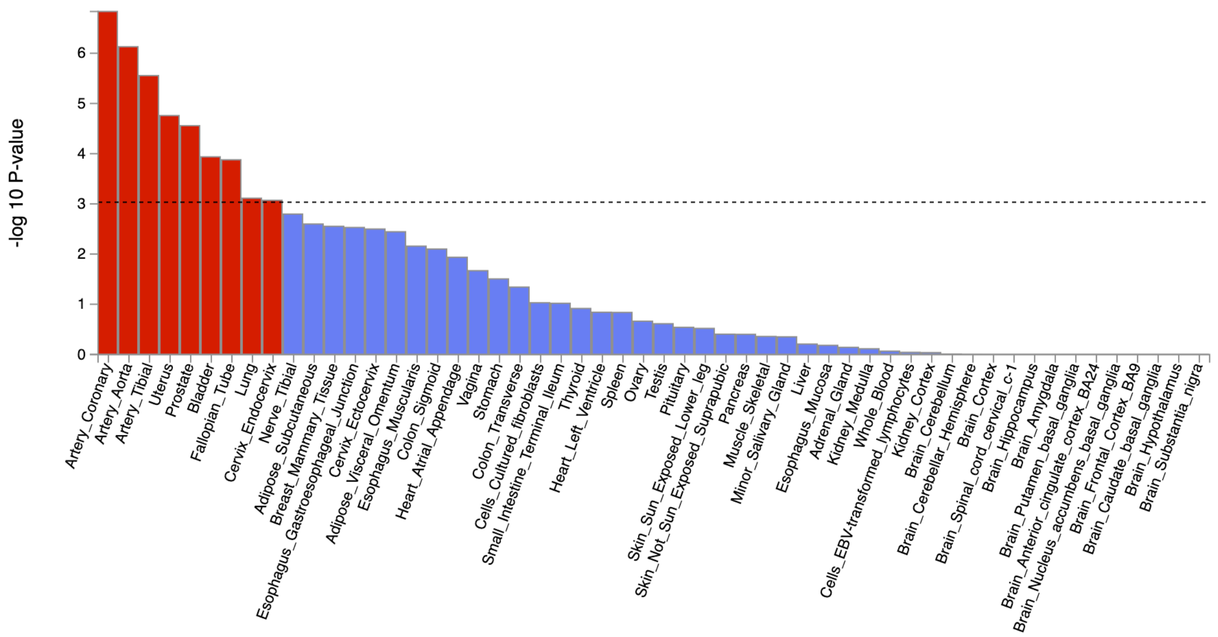


Figure 13. **Tissue-specific gene expression profile enrichment of TAA in MTAG CAD+TAA multi-trait meta-analysis**. Enrichment estimated with MAGMA version 1.08 gene-property analysis as implemented in FUMA version 1.3.4. Tested tissue gene expression profiles are listed on the X-axis. Strength of the association is depicted on the Y-axis as -$\log_{10}(P)$.

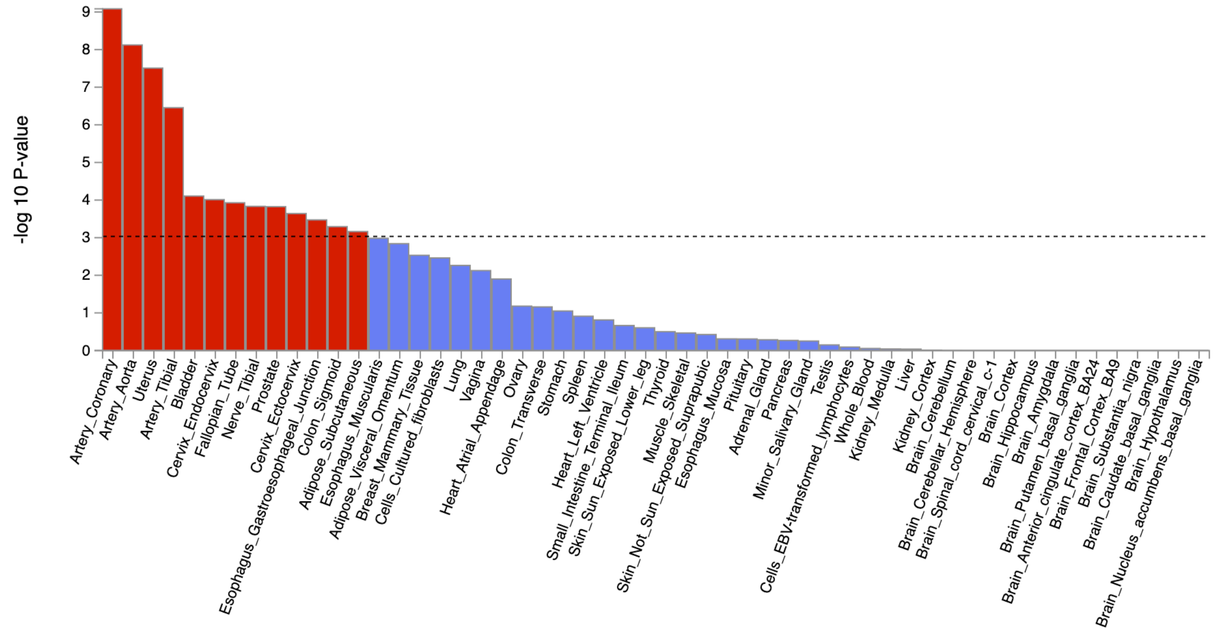


Figure 14. **Tissue-specific gene expression profile enrichment of TAA in MTAG AAA+TAA multi-trait meta-analysis**. Enrichment estimated with MAGMA version 1.08 gene-property analysis as implemented in FUMA version 1.3.4. Tested tissue gene expression profiles are listed on the X-axis. Strength of the association is depicted on the Y-axis as -$\log_{10}(P)$.

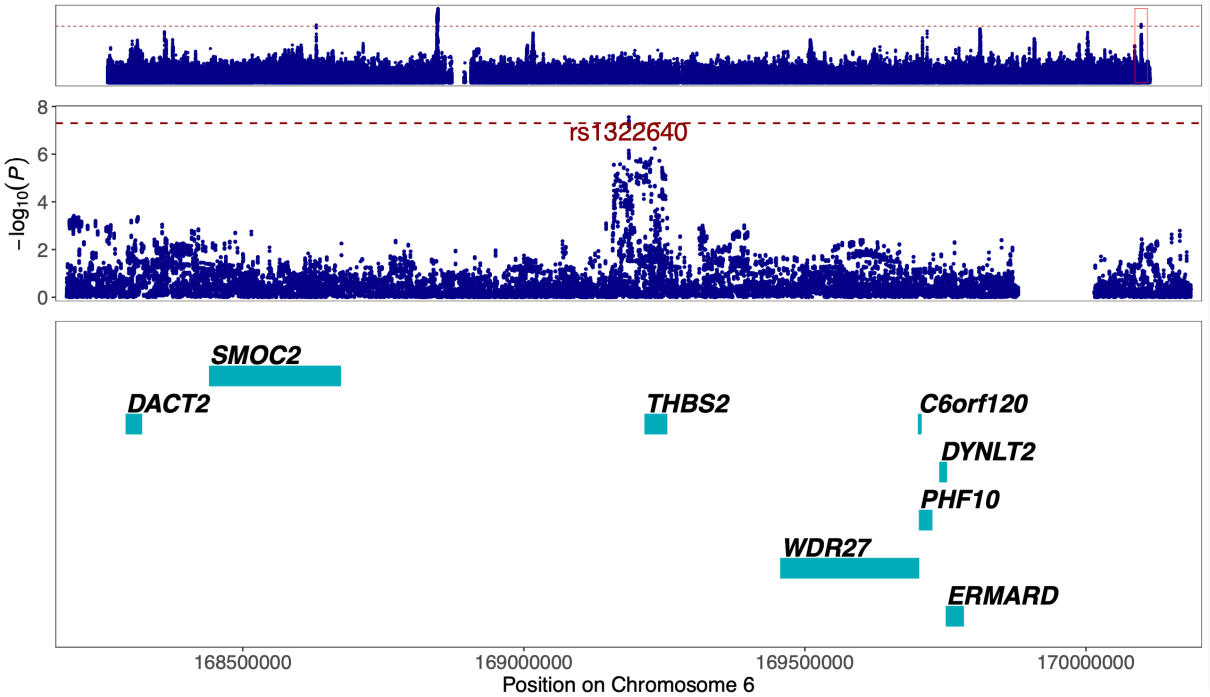


Figure 15. **Regional association plot of previously unreported TAA signal at 6q27 around the GWAS lead variant rs1322640.** The most likely causal gene explaining the association at the locus was predicted to be *THBS2* (*Thrombospondin 2*). The top panel shows an overview of the genomic position, the middle and bottom panels depict individual SNVs and genes within the 2 MB locus window on chromosome 6. The middle Y-axis displays the strength of the association as $-\log_{10}(P)$ with a dashed red line marking the threshold for genome-wide significance ($P < 5 \times 10^{-8}$).

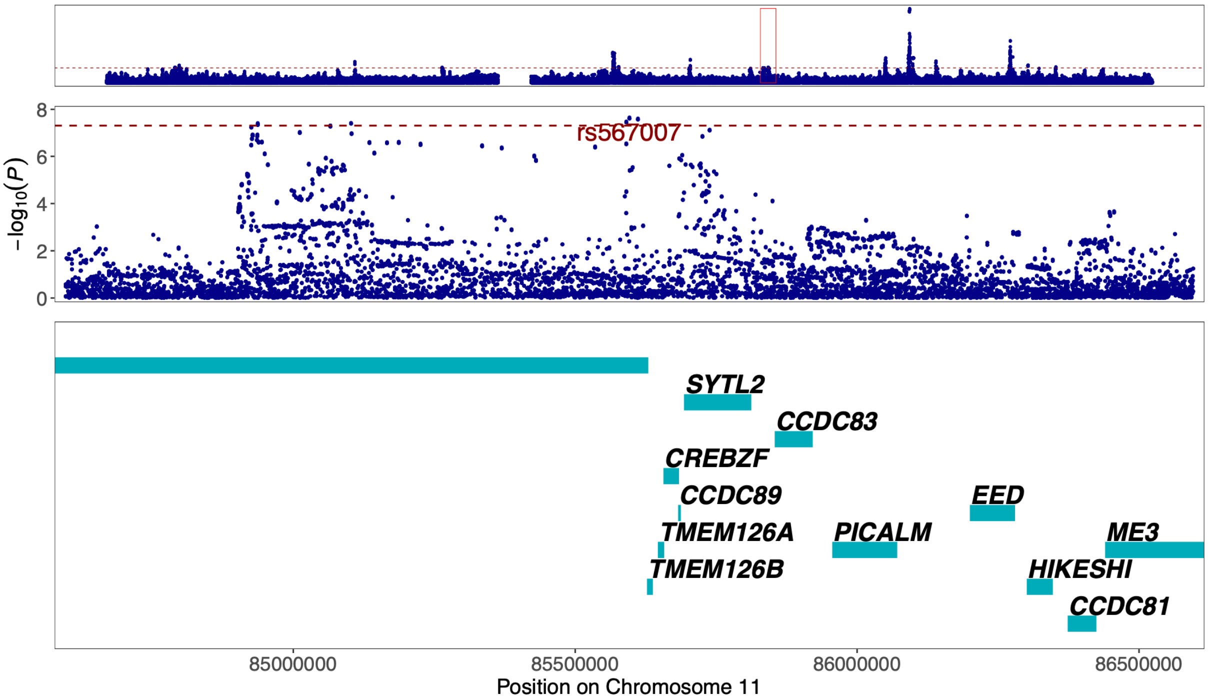


Figure 16. **Regional association plot of previously unreported CAD signal at 11q14.1 around the GWAS lead variant rs567007.** The most likely causal gene explaining the association at the locus was predicted to be *DLG2* (*Discs Large MAGUK Scaffold Protein 2*). The top panel shows an overview of the genomic position, the middle and bottom panels depict individual SNVs and genes within the 2 MB locus window on chromosome 11. The middle Y-axis displays the strength of the association as $-\log_{10}(P)$ with a dashed red line marking the threshold for genome-wide significance ($P < 5 \times 10^{-8}$).

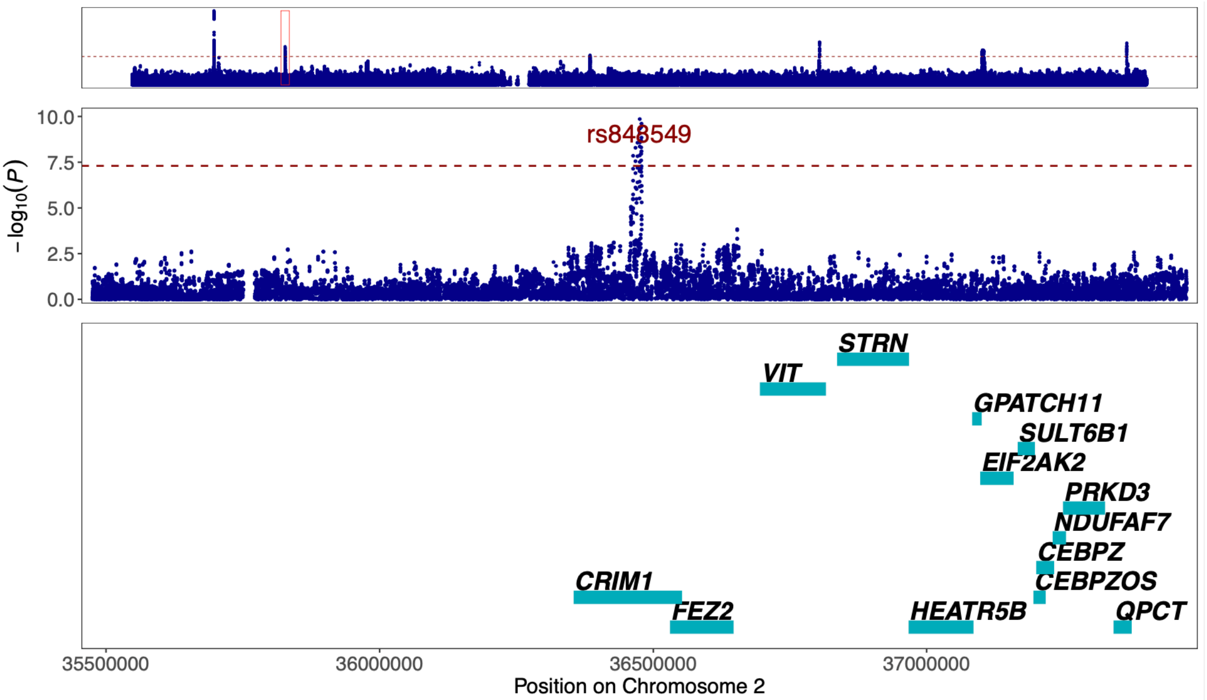


Figure 17. **Regional association plot of previously unreported TAA signal at 2p22.2 around the GWAS lead variant rs848549.** The most likely causal gene explaining the association at the locus was predicted to be *CRIM1* (*Cysteine Rich Transmembrane BMP Regulator 1*). The top panel shows an overview of the genomic position, the middle and bottom panels depict individual SNVs and genes within the 2 MB locus window on chromosome 2. The middle Y-axis displays the strength of the association as -log$_{10}$($P$) with a dashed red line marking the threshold for genome-wide significance ($P < 5 \times 10^{-8}$).

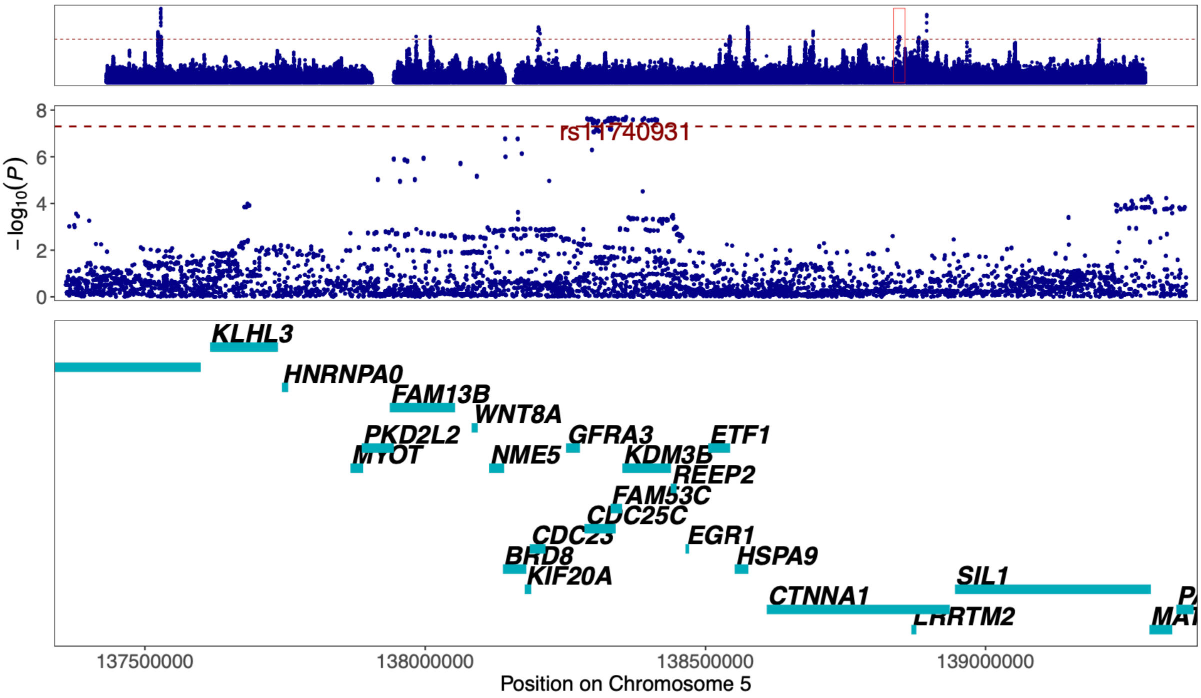


Figure 18. **Regional association plot of previously unreported CAD signal at 5q31.2 around the GWAS lead variant rs11740931.** The most likely causal gene explaining the association at the locus was predicted to be *KDM3B* (*Lysine Demethylase 3B*). The top panel shows an overview of the genomic position, the middle and bottom panels depict individual SNVs and genes within the 2 MB locus window on chromosome 5. The middle Y-axis displays the strength of the association as -log$_{10}$($P$) with a dashed red line marking the threshold for genome-wide significance ($P < 5 \times 10^{-8}$).

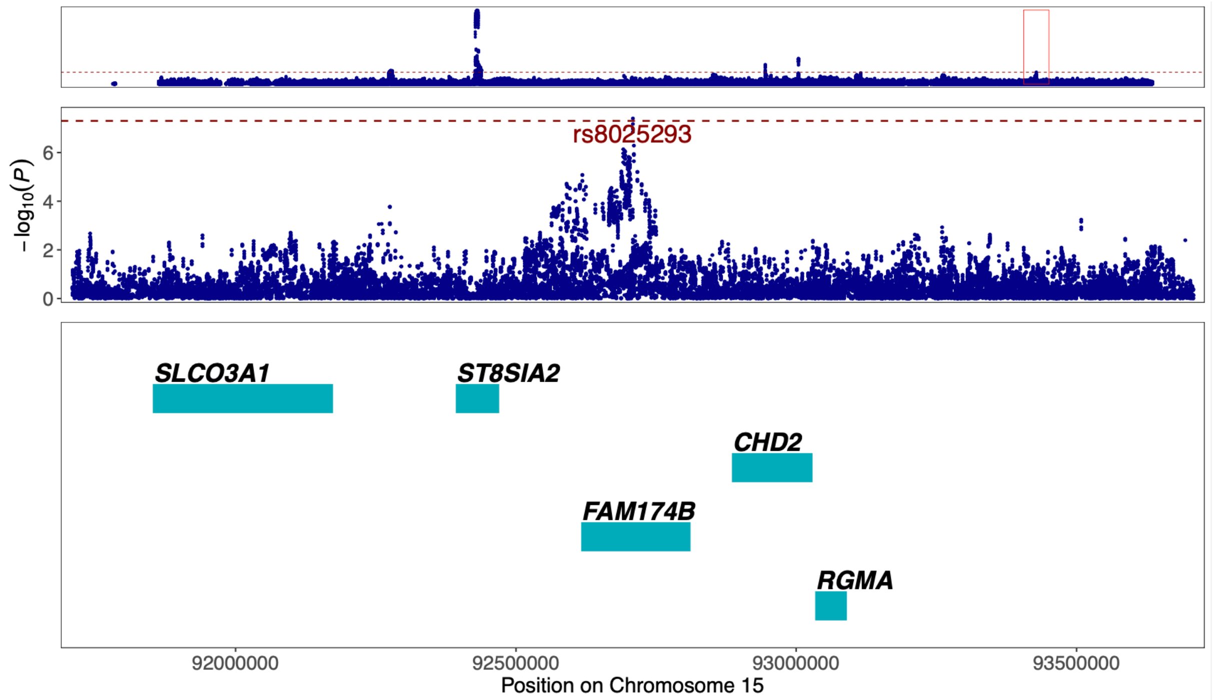


Figure 19. **Regional association plot of previously unreported TAA signal at 15q26.1 around the GWAS lead variant rs8025293.** The most likely causal gene explaining the association at the locus was predicted to be *FAM174B* (*Family With Sequence Similarity 174 Member B*). The top panel shows an overview of the genomic position, the middle and bottom panels depict individual SNVs and genes within the 2 MB locus window on chromosome 15. The middle Y-axis displays the strength of the association as $-\log_{10}(P)$ with a dashed red line marking the threshold for genome-wide significance ($P < 5 \times 10^{-8}$).

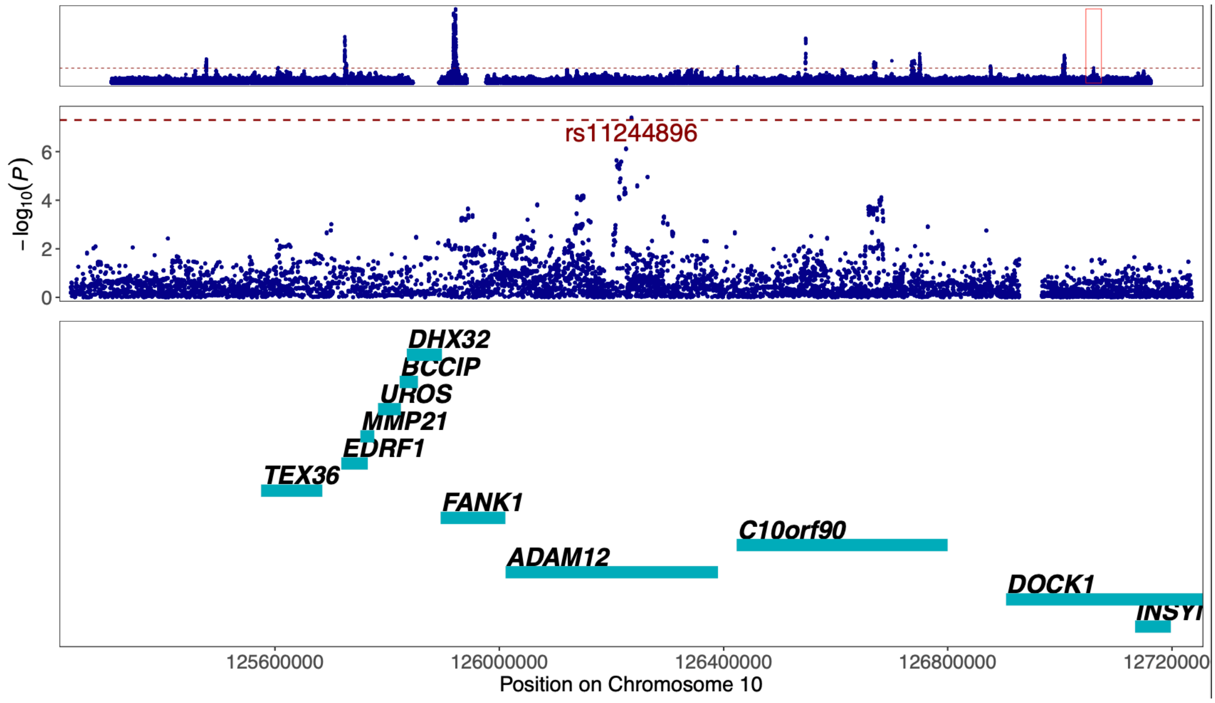


Figure 20. **Regional association plot of previously unreported CAD signal at 10q26.2 around the GWAS lead variant rs11244896.** The most likely causal gene explaining the association at the locus was predicted to be *ADAM12* (*ADAM Metallopeptidase Domain 12*). The top panel shows an overview of the genomic position, the middle and bottom panels depict individual SNVs and genes within the 2 MB locus window on chromosome 10. The middle Y-axis displays the strength of the association as $-\log_{10}(P)$ with a dashed red line marking the threshold for genome-wide significance ($P < 5 \times 10^{-8}$).

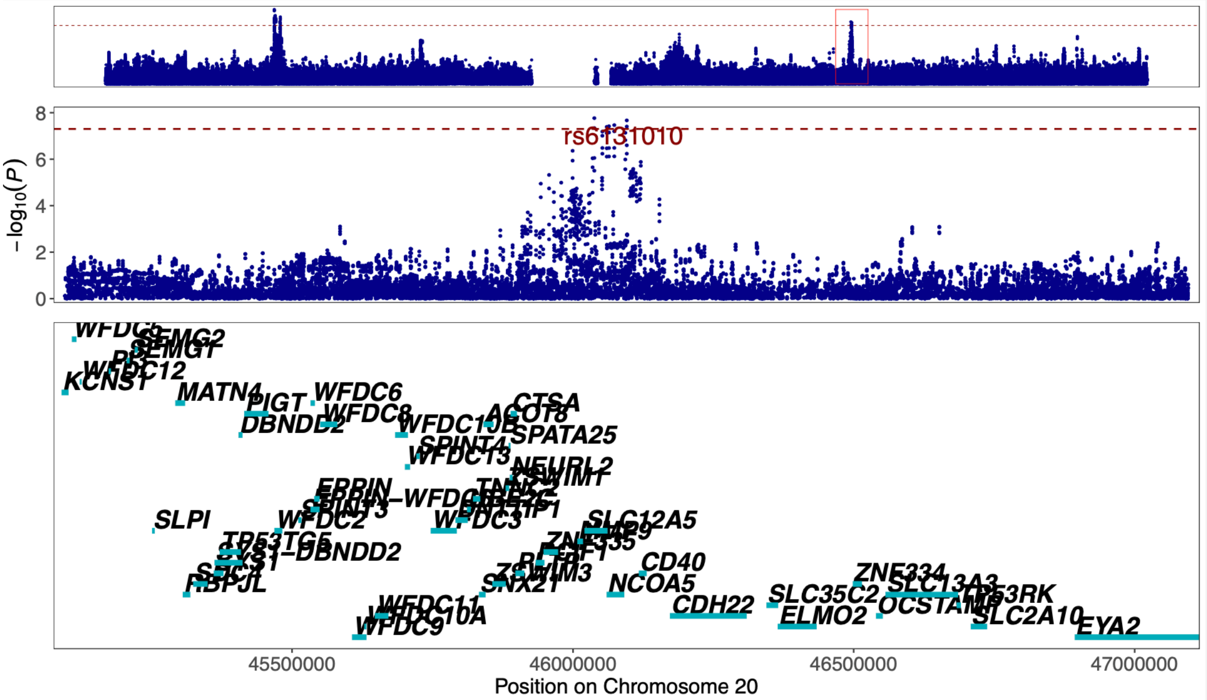


Figure 21. **Regional association plot of previously unreported TAA signal at 20q13.12 around the GWAS lead variant rs6131010.** The most likely causal gene explaining the association at the locus was predicted to be *CD40* (*CD40 Molecule*). The top panel shows an overview of the genomic position, the middle and bottom panels depict individual SNVs and genes within the 2 MB locus window on chromosome 20. The middle Y-axis displays the strength of the association as -log10($P$) with a dashed red line marking the threshold for genome-wide significance ($P < 5 \times 10^{-8}$).

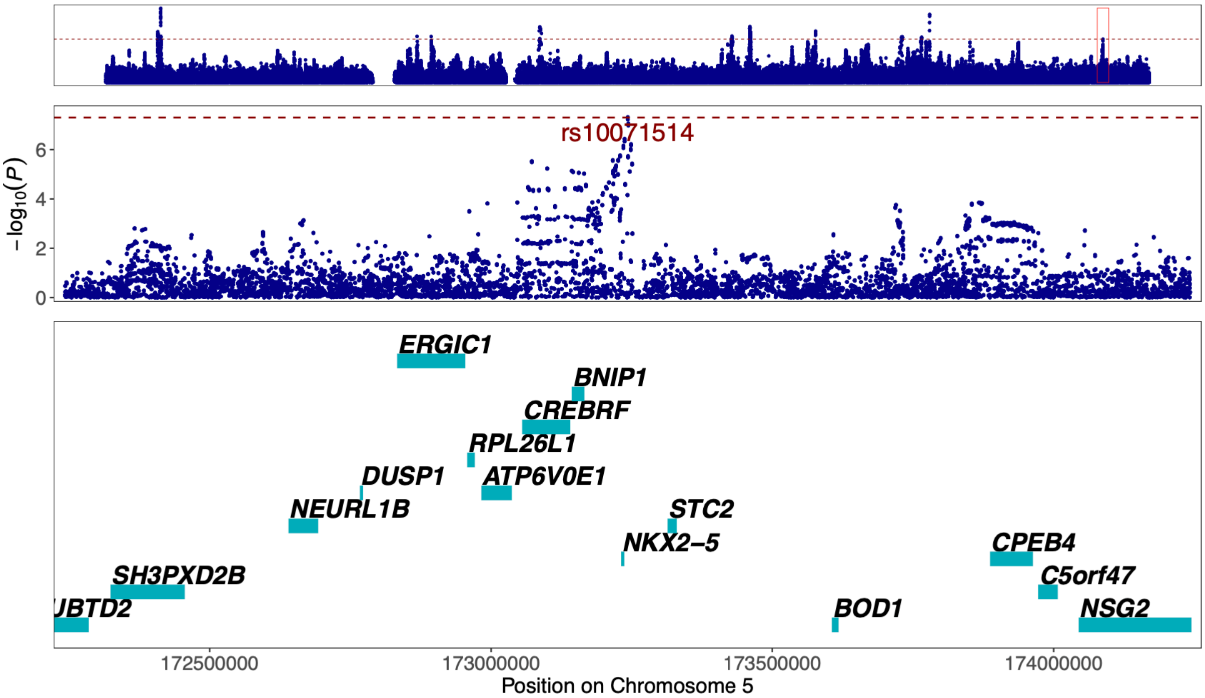


Figure 22. **Regional association plot of previously unreported CAD signal at 5q35.1 around the GWAS lead variant rs10071514.** The most likely causal gene explaining the association at the locus was predicted to be *NKX2-5* (*NK2 Homeobox 5*). The top panel shows an overview of the genomic position, the middle and bottom panels depict individual SNVs and genes within the 2 MB locus window on chromosome 5. The middle Y-axis displays the strength of the association as -log10($P$) with a dashed red line marking the threshold for genome-wide significance ($P < 5 \times 10^{-8}$).

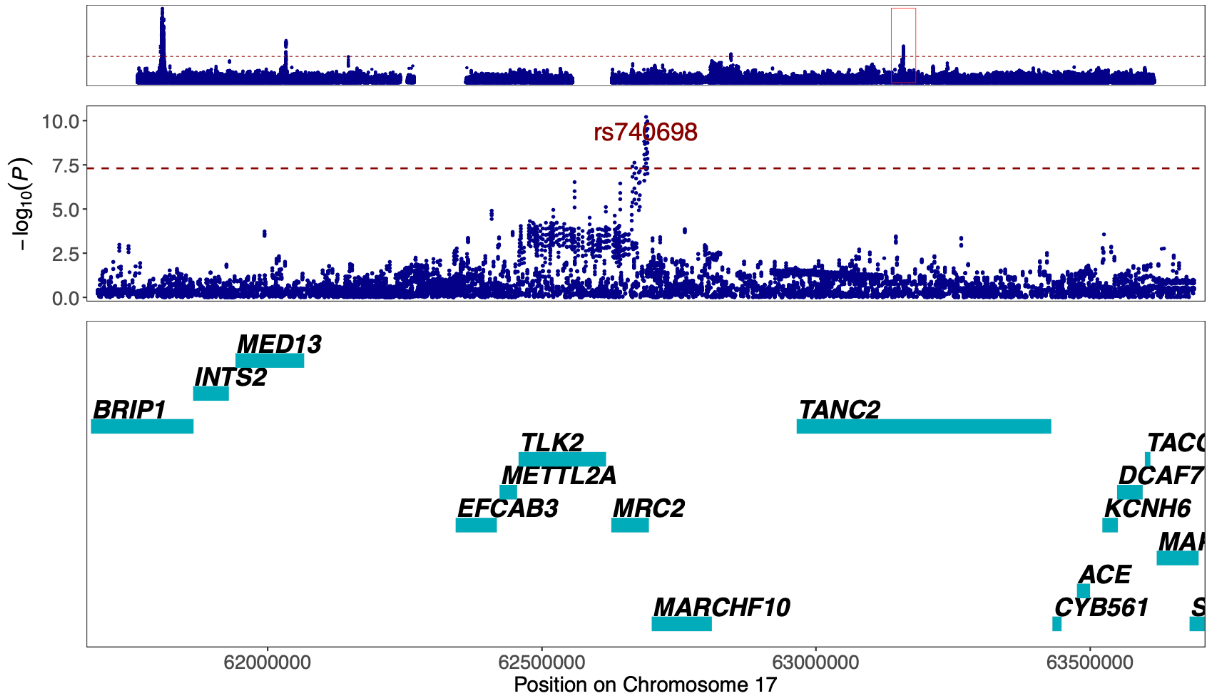


Figure 23. **Regional association plot of previously unreported TAA signal at 17q23.2 around the GWAS lead variant rs740698.** The most likely causal gene explaining the association at the locus was predicted to be *MRC2* (*Mannose Receptor C-Type 2*). The top panel shows an overview of the genomic position, the middle and bottom panels depict individual SNVs and genes within the 2 MB locus window on chromosome 17. The middle Y-axis displays the strength of the association as -log$_{10}$(*P*) with a dashed red line marking the threshold for genome-wide significance ($P < 5 \times 10^{-8}$).

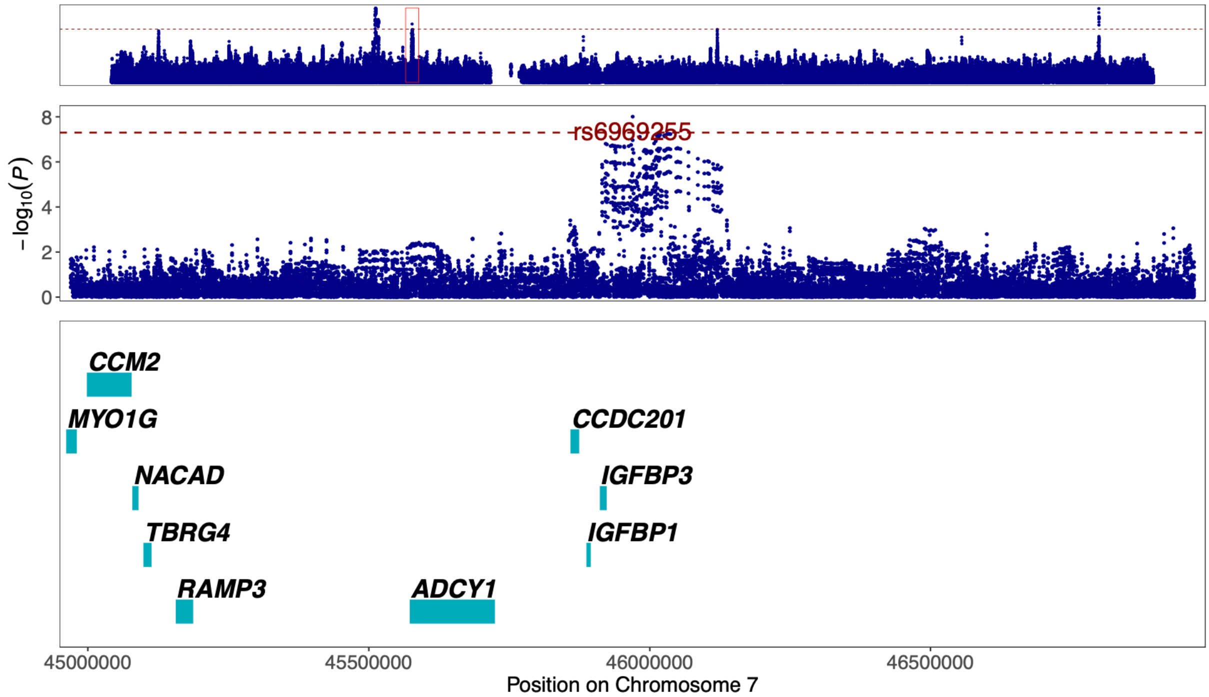


Figure 24. **Regional association plot of previously unreported AAA signal at 7p12.3 around the GWAS lead variant rs6969255.** The most likely causal gene explaining the association at the locus was predicted to be *IGFBP3* (*Insulin Like Growth Factor Binding Protein 3*). The top panel shows an overview of the genomic position, the middle and bottom panels depict individual SNVs and genes within the 2 MB locus window on chromosome 7. The middle Y-axis displays the strength of the association as -log$_{10}$(*P*) with a dashed red line marking the threshold for genome-wide significance ($P < 5 \times 10^{-8}$).

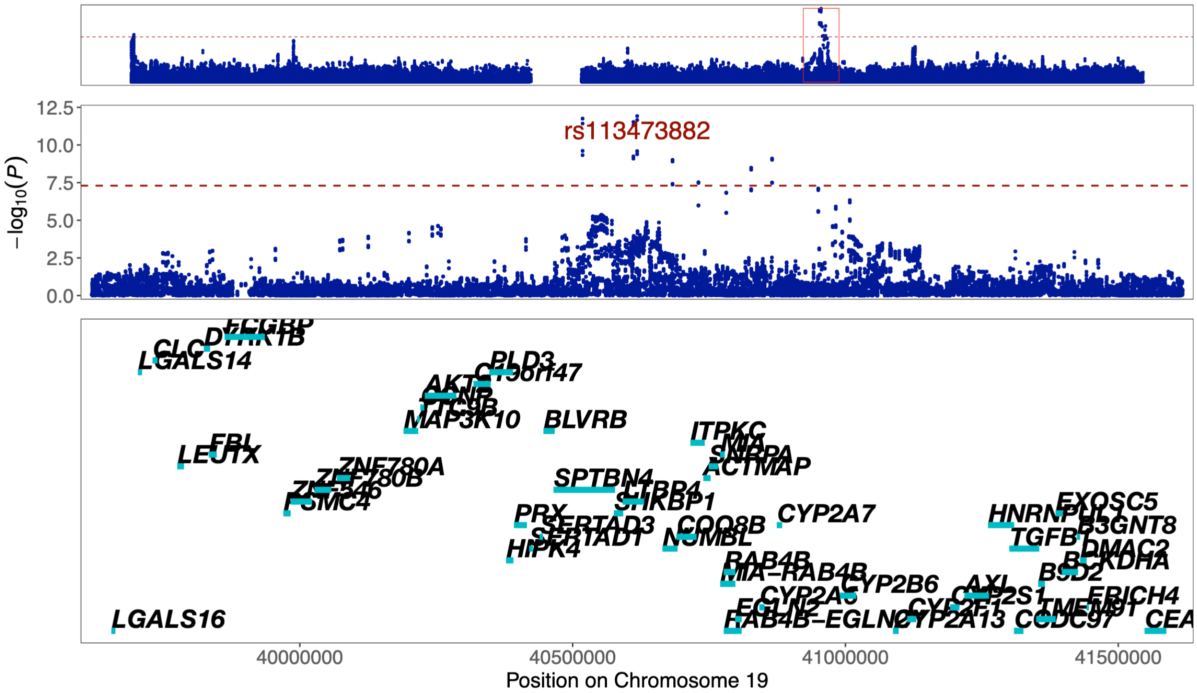


Figure 25. **Regional association plot of previously unreported TAA signal at 19q13.2 around the GWAS lead variant rs113473882.** The most likely causal gene explaining the association at the locus was predicted to be *BLVRB* (*Biliverdin Reductase B*). The top panel shows an overview of the genomic position, the middle and bottom panels depict individual SNVs and genes within the 2 MB locus window on chromosome 19. The middle Y-axis displays the strength of the association as -log$_{10}$($P$) with a dashed red line marking the threshold for genome-wide significance ($P < 5 \times 10^{-8}$).

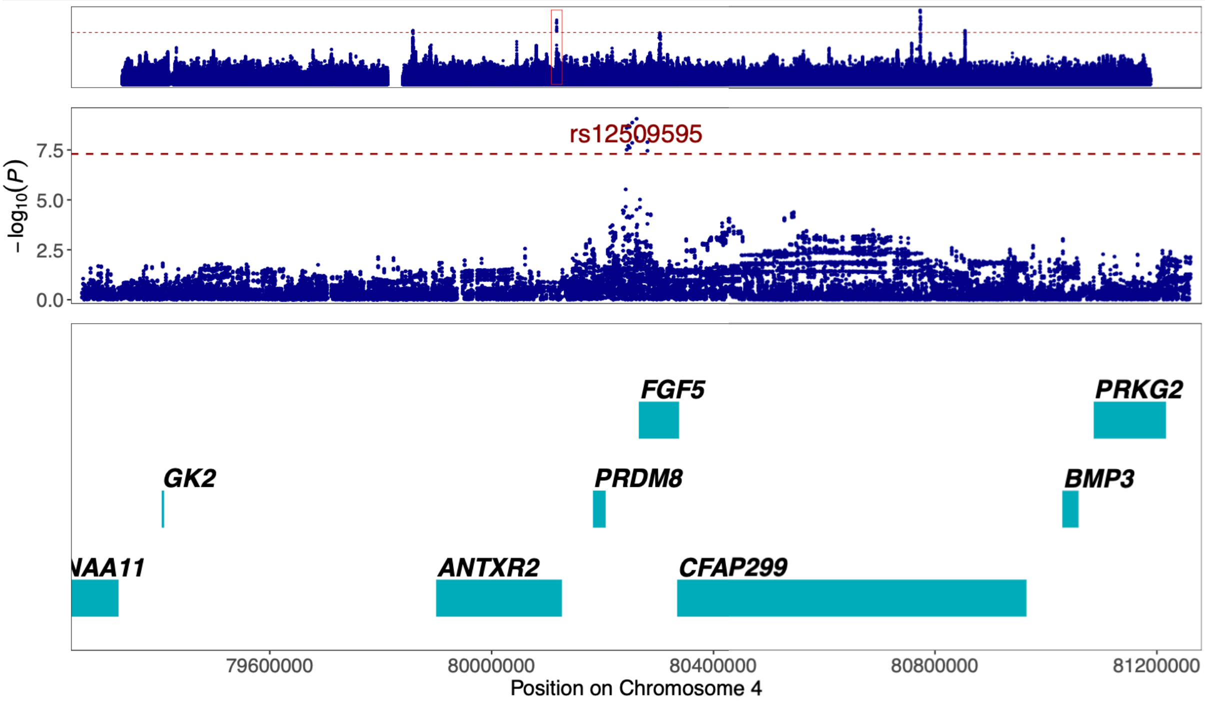


Figure 26. **Regional association plot of previously unreported AAA signal at 4q21.21 around the GWAS lead variant rs12509595.** The most likely causal gene explaining the association at the locus was predicted to be *FGF5* (*Fibroblast Growth Factor 5*). The top panel shows an overview of the genomic position, the middle and bottom panels depict individual SNVs and genes within the 2 MB locus window on chromosome 4. The middle Y-axis displays the strength of the association as -log$_{10}$($P$) with a dashed red line marking the threshold for genome-wide significance ($P < 5 \times 10^{-8}$).

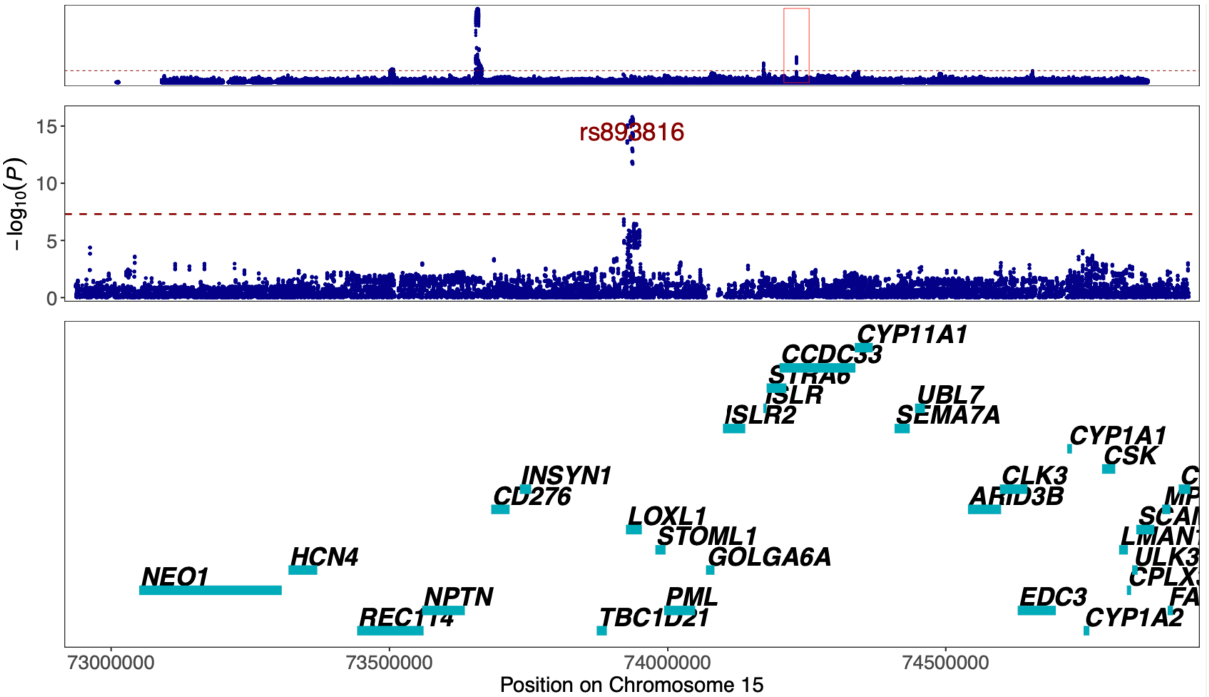


Figure 27. **Regional association plot of previously unreported TAA signal at 15q24.1 around the GWAS lead variant rs893816.** The most likely causal gene explaining the association at the locus was predicted to be *LOXL1* (*Lysyl Oxidase Like 1*). The top panel shows an overview of the genomic position, the middle and bottom panels depict individual SNVs and genes within the 2 MB locus window on chromosome 15. The middle Y-axis displays the strength of the association as -log10($P$) with a dashed red line marking the threshold for genome-wide significance ($P < 5 \times 10^{-8}$).

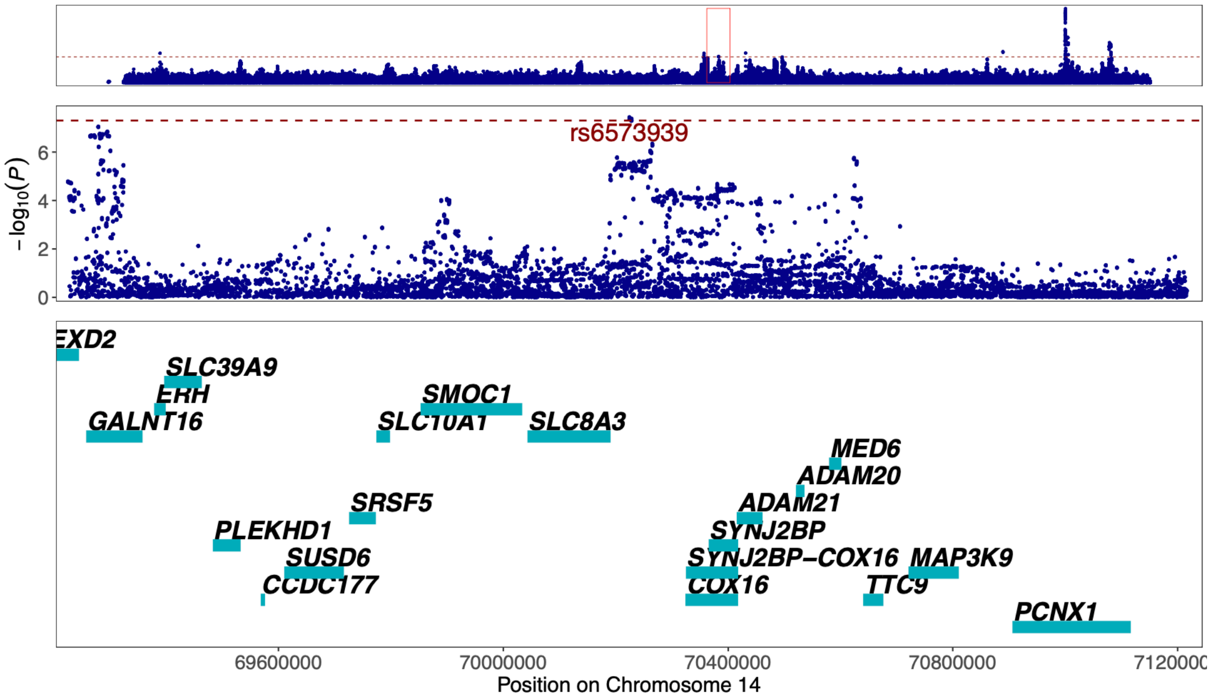


Figure 28. **Regional association plot of previously unreported CAD signal at 14q24.2 around the GWAS lead variant rs6573939.** The most likely causal gene explaining the association at the locus was predicted to be *COX16* (*Cytochrome C Oxidase Assembly Factor COX16*). The top panel shows an overview of the genomic position, the middle and bottom panels depict individual SNVs and genes within the 2 MB locus window on chromosome 14. The middle Y-axis displays the strength of the association as -log10($P$) with a dashed red line marking the threshold for genome-wide significance ($P < 5 \times 10^{-8}$).

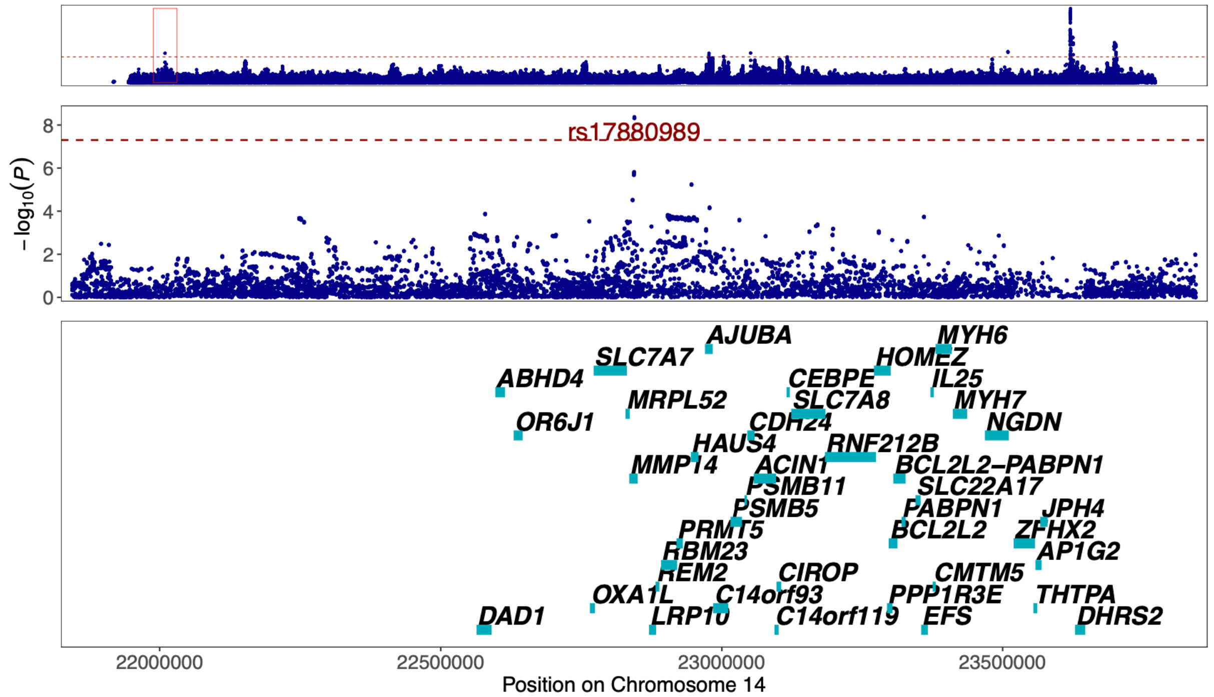


Figure 29. **Regional association plot of previously unreported CAD signal at 14q11.2 around the GWAS lead variant rs17880989.** The most likely causal gene explaining the association at the locus was predicted to be *MMP14* (*Matrix Metallopeptidase 14*). The top panel shows an overview of the genomic position, the middle and bottom panels depict individual SNVs and genes within the 2 MB locus window on chromosome 14. The middle Y-axis displays the strength of the association as -log10($P$) with a dashed red line marking the threshold for genome-wide significance ($P < 5 \times 10^{-8}$).

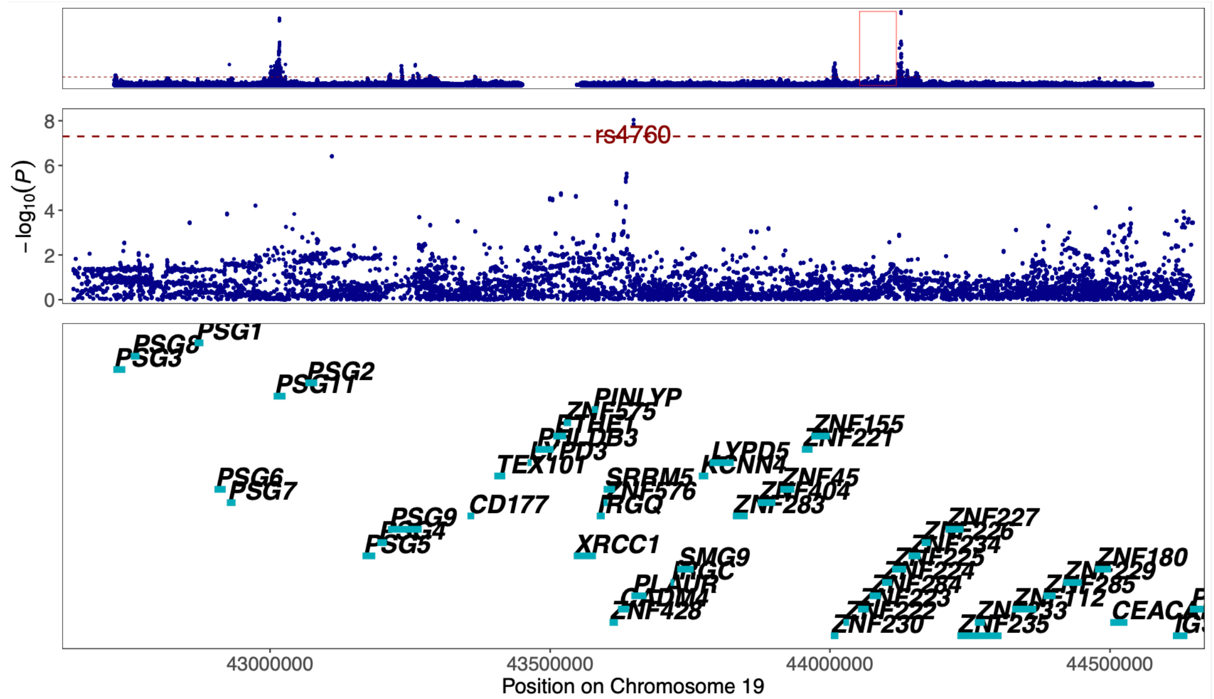


Figure 30. **Regional association plot of previously unreported CAD signal at 19q13.31 around the GWAS lead variant rs4760.** The most likely causal gene explaining the association at the locus was predicted to be *PLAUR* (*Plasminogen Activator, Urokinase Receptor*). The top panel shows an overview of the genomic position, the middle and bottom panels depict individual SNVs and genes within the 2 MB locus window on chromosome 19. The middle Y-axis displays the strength of the association as -log10($P$) with a dashed red line marking the threshold for genome-wide significance ($P < 5 \times 10^{-8}$).

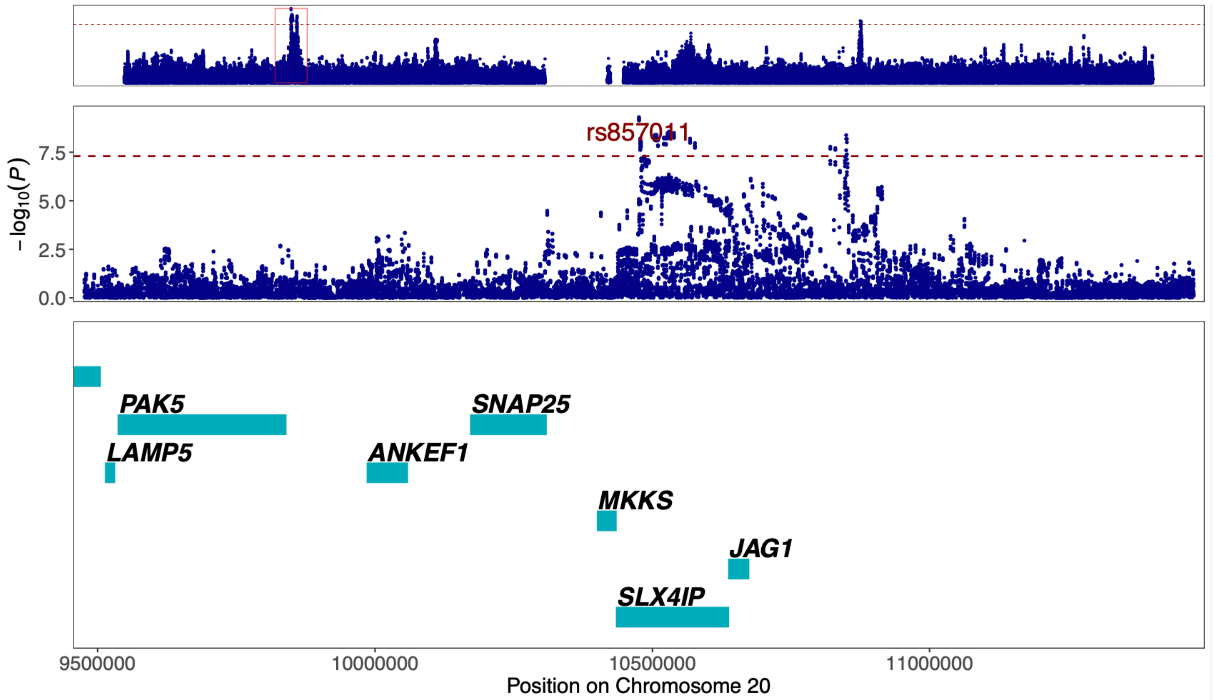


Figure 31. **Regional association plot of previously unreported TAA signal at 20p12.2 around the GWAS lead variant rs857011.** The most likely causal gene explaining the association at the locus was predicted to be *SLX4IP* (*SLX4 Interacting Protein*). The top panel shows an overview of the genomic position, the middle and bottom panels depict individual SNVs and genes within the 2 MB locus window on chromosome 20. The middle Y-axis displays the strength of the association as -$\log_{10}$(*P*) with a dashed red line marking the threshold for genome-wide significance ($P < 5 \times 10^{-8}$).

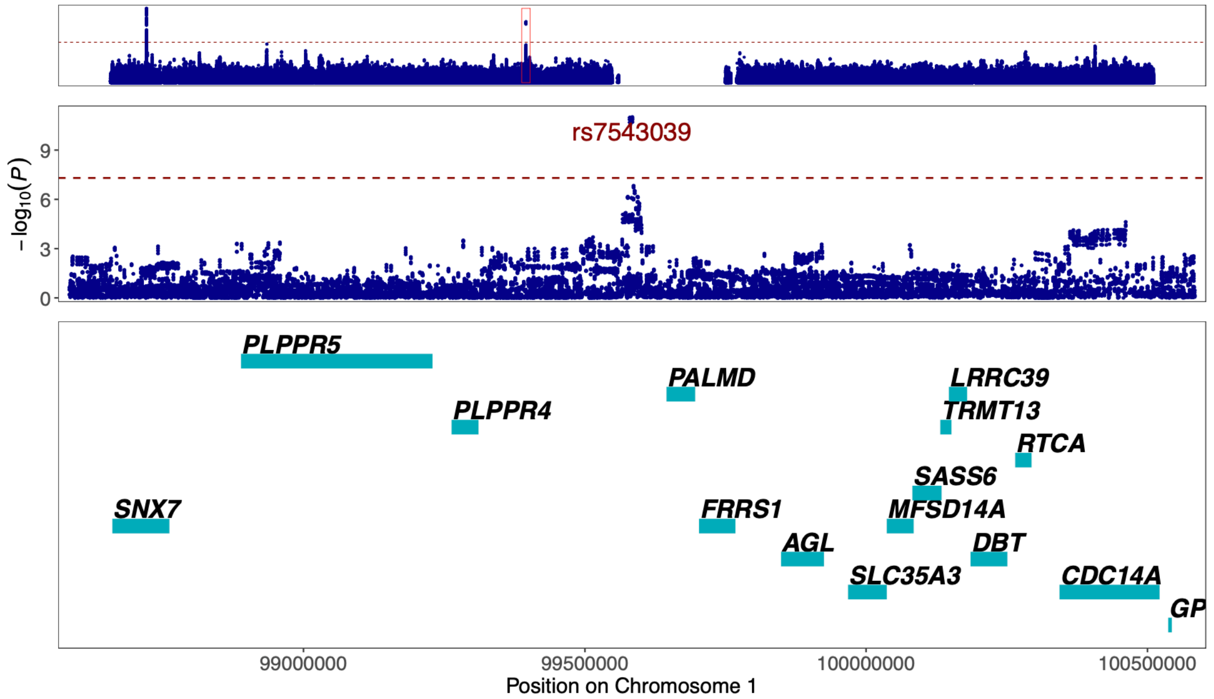


Figure 32. **Regional association plot of previously unreported TAA signal at 1p21.2 around the GWAS lead variant rs7543039.** The most likely causal gene explaining the association at the locus was predicted to be *PALMD* (*Palmdelphin*). The top panel shows an overview of the genomic position, the middle and bottom panels depict individual SNVs and genes within the 2 MB locus window on chromosome 1. The middle Y-axis displays the strength of the association as -$\log_{10}$(*P*) with a dashed red line marking the threshold for genome-wide significance ($P < 5 \times 10^{-8}$).

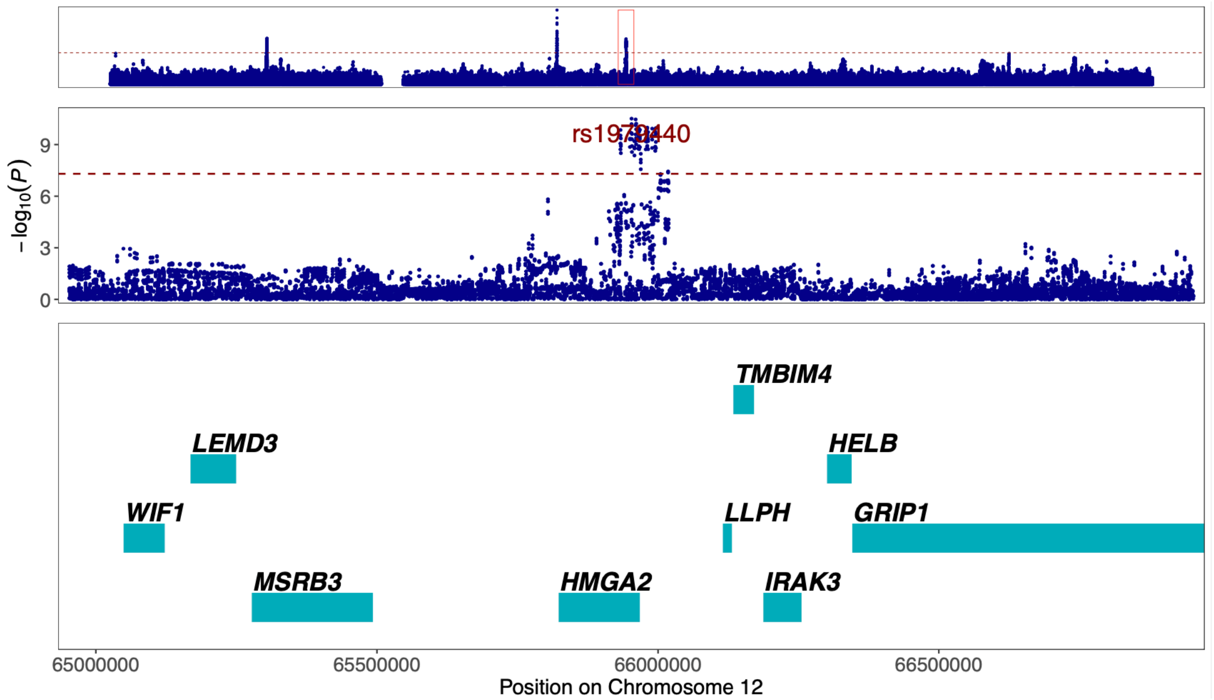


Figure 33. **Regional association plot of previously unreported TAA signal at 12q14.3 around the GWAS lead variant rs1979440.** The most likely causal gene explaining the association at the locus was predicted to be *HMGA2* (*High Mobility Group AT-Hook 2*). The top panel shows an overview of the genomic position, the middle and bottom panels depict individual SNVs and genes within the 2 MB locus window on chromosome 12. The middle Y-axis displays the strength of the association as -$\log_{10}$(*P*) with a dashed red line marking the threshold for genome-wide significance ($P < 5 \times 10^{-8}$).

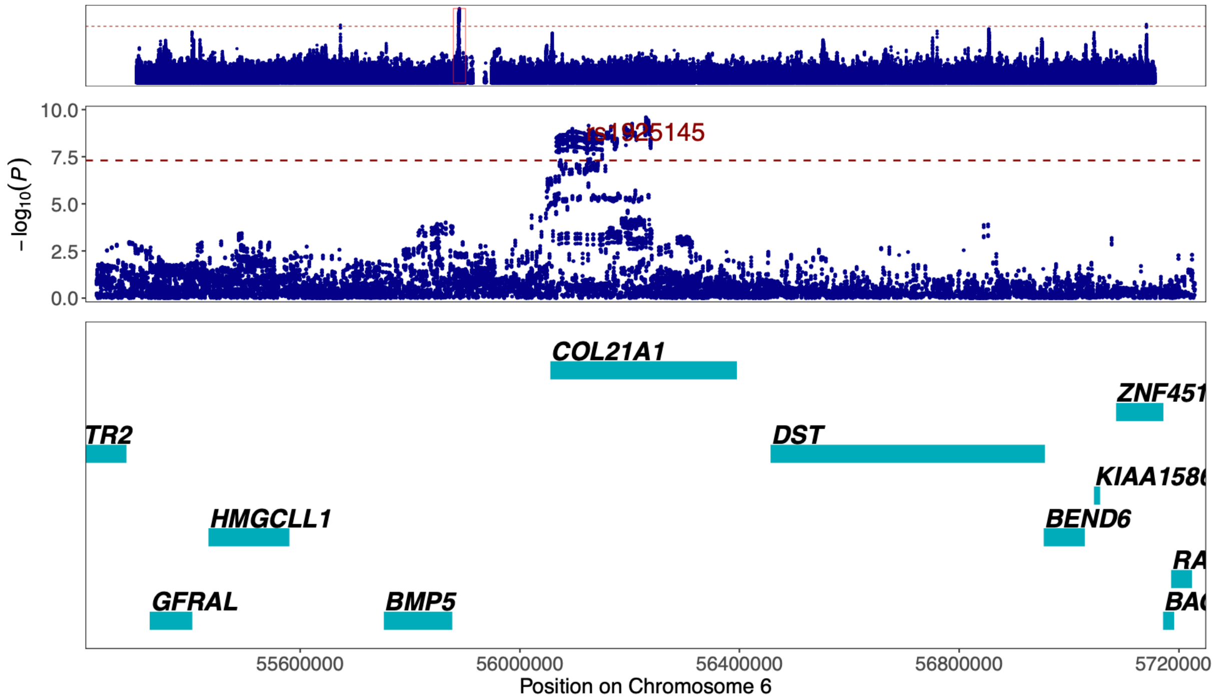


Figure 34. **Regional association plot of previously unreported TAA signal at 6p12.1 around the GWAS lead variant rs1925145.** The most likely causal gene explaining the association at the locus was predicted to be *COL21A1* (*Collagen Type XXI Alpha 1 Chain*). The top panel shows an overview of the genomic position, the middle and bottom panels depict individual SNVs and genes within the 2 MB locus window on chromosome 6. The middle Y-axis displays the strength of the association as -$\log_{10}$(*P*) with a dashed red line marking the threshold for genome-wide significance ($P < 5 \times 10^{-8}$).

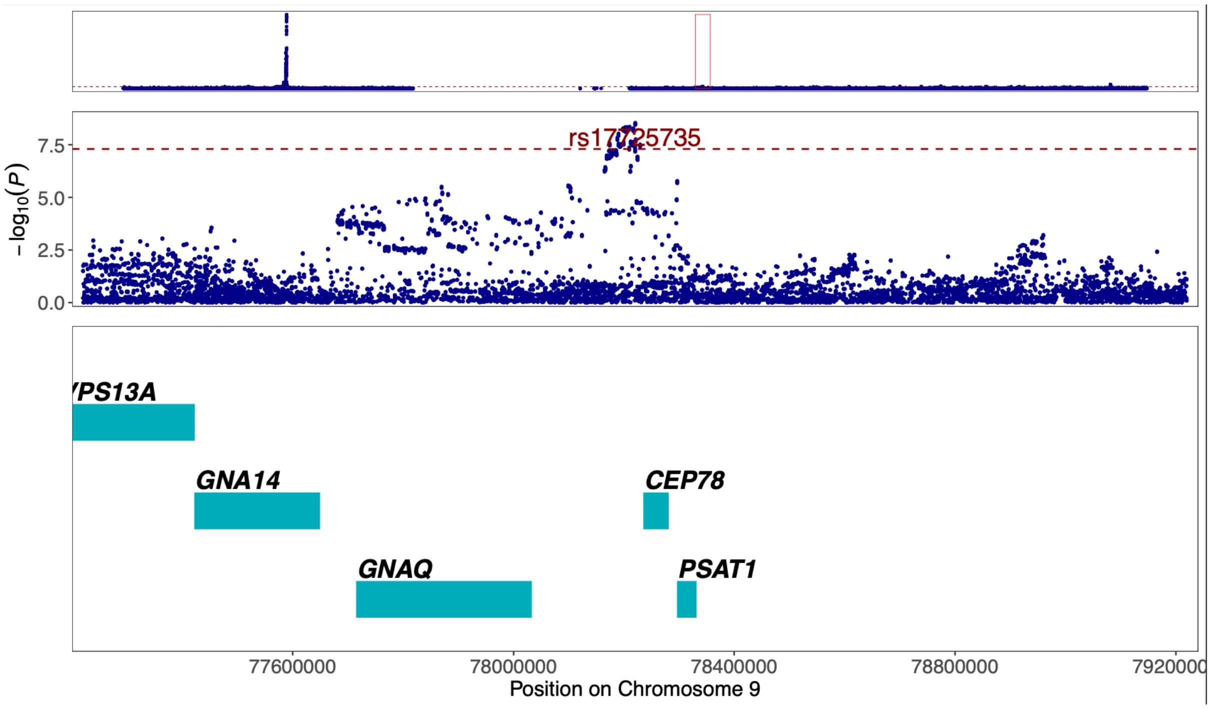


Figure 35. **Regional association plot of previously unreported CAD signal at 9q21.2 around the GWAS lead variant rs17725735.** The most likely causal gene explaining the association at the locus was predicted to be *GNAQ* (*G Protein Subunit Alpha Q*). The top panel shows an overview of the genomic position, the middle and bottom panels depict individual SNVs and genes within the 2 MB locus window on . The middle Y-axis displays the strength of the association as -$\log_{10}(P)$ with a dashed red line marking the threshold for genome-wide significance ($P < 5 \times 10^{-8}$).

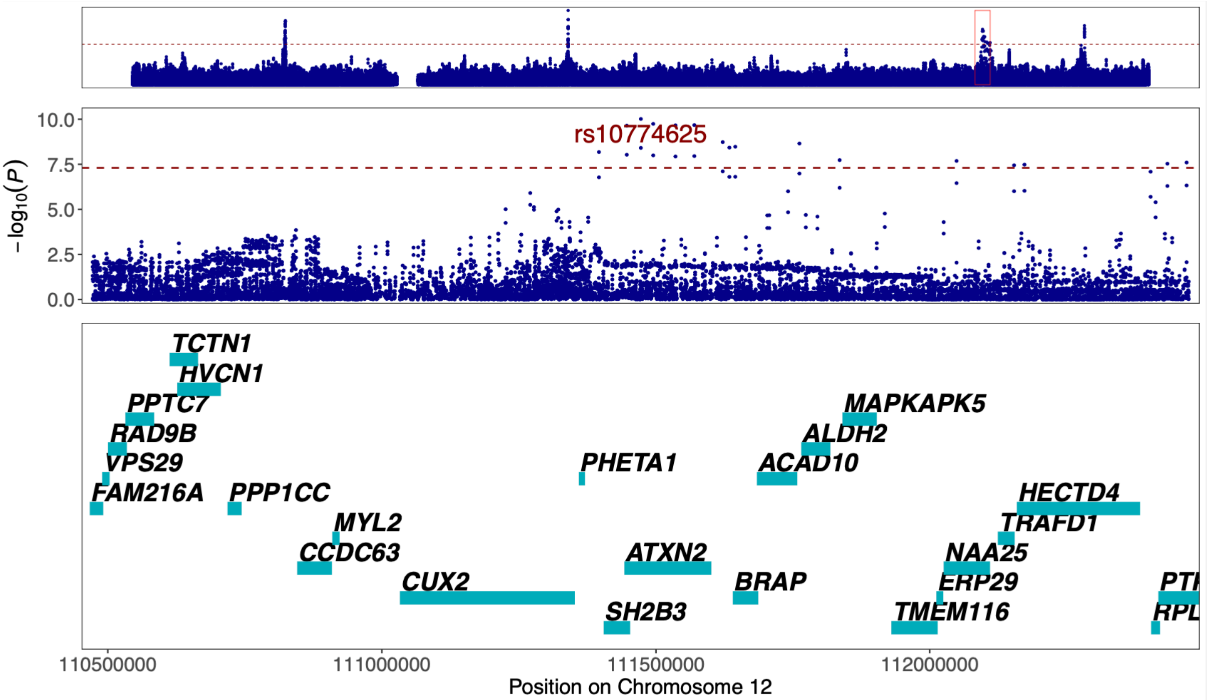


Figure 36. **Regional association plot of previously unreported AAA signal at 12q24.12 around the GWAS lead variant rs10774625.** The most likely causal gene explaining the association at the locus was predicted to be *SH2B3* (*SH2B Adaptor Protein 3*). The top panel shows an overview of the genomic position, the middle and bottom panels depict individual SNVs and genes within the 2 MB locus window on chromosome 12. The middle Y-axis displays the strength of the association as -$\log_{10}(P)$ with a dashed red line marking the threshold for genome-wide significance ($P < 5 \times 10^{-8}$).

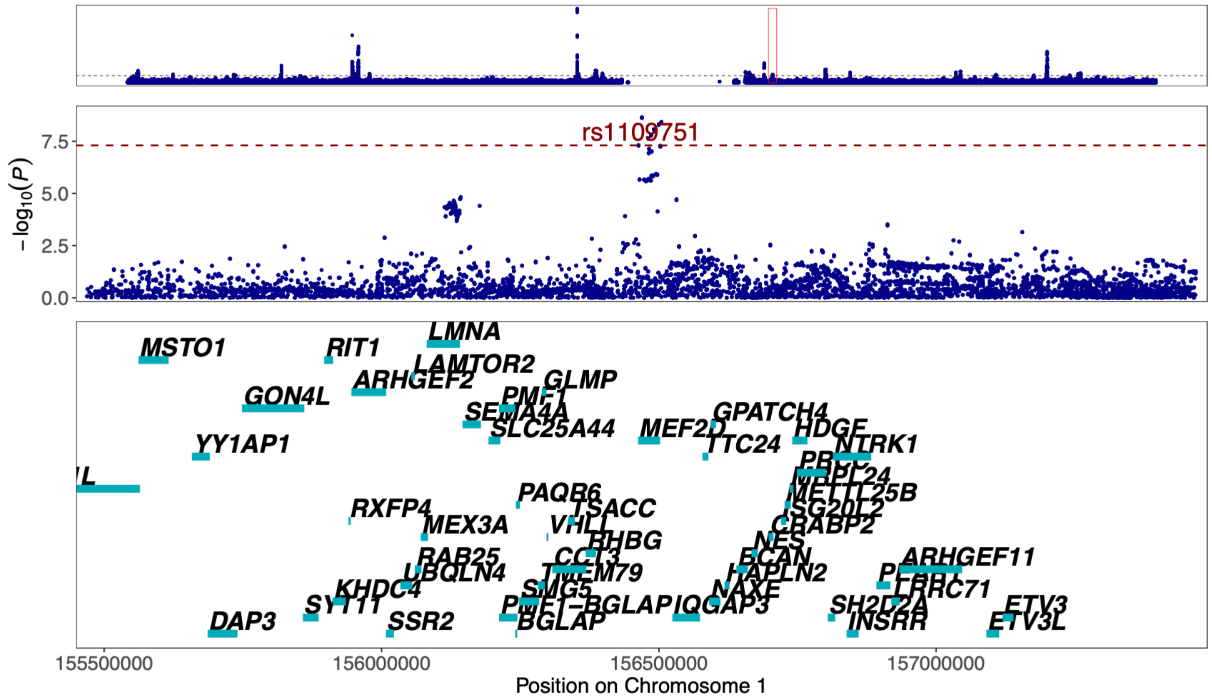


Figure 37. **Regional association plot of previously unreported CAD signal at 1q22 around the GWAS lead variant rs1109751.** The most likely causal gene explaining the association at the locus was predicted to be *MEF2D* (*Myocyte Enhancer Factor 2D*). The top panel shows an overview of the genomic position, the middle and bottom panels depict individual SNVs and genes within the 2 MB locus window on chromosome 1. The middle Y-axis displays the strength of the association as -log10(*P*) with a dashed red line marking the threshold for genome-wide significance ($P < 5 \times 10^{-8}$).

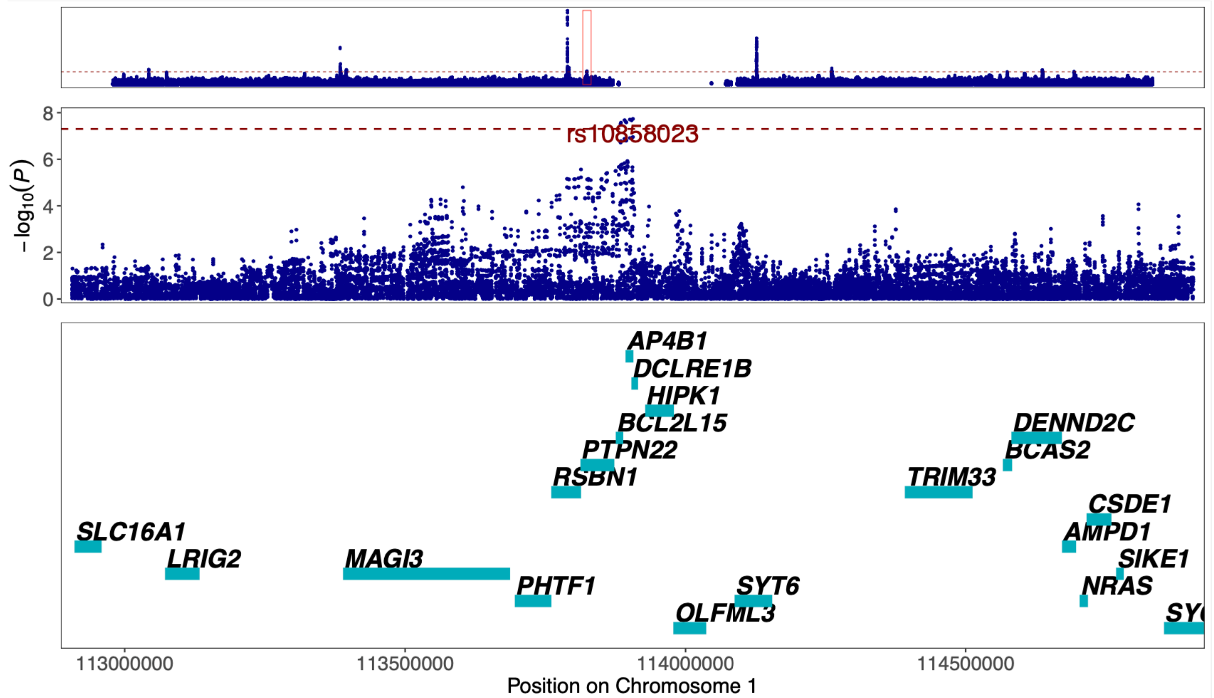


Figure 38. **Regional association plot of previously unreported AAA signal at 1p13.2 around the GWAS lead variant rs10858023.** The most likely causal gene explaining the association at the locus was predicted to be *DCLRE1B* (*DNA Cross-Link Repair 1B*). The top panel shows an overview of the genomic position, the middle and bottom panels depict individual SNVs and genes within the 2 MB locus window on chromosome 1. The middle Y-axis displays the strength of the association as -log10(*P*) with a dashed red line marking the threshold for genome-wide significance ($P < 5 \times 10^{-8}$).

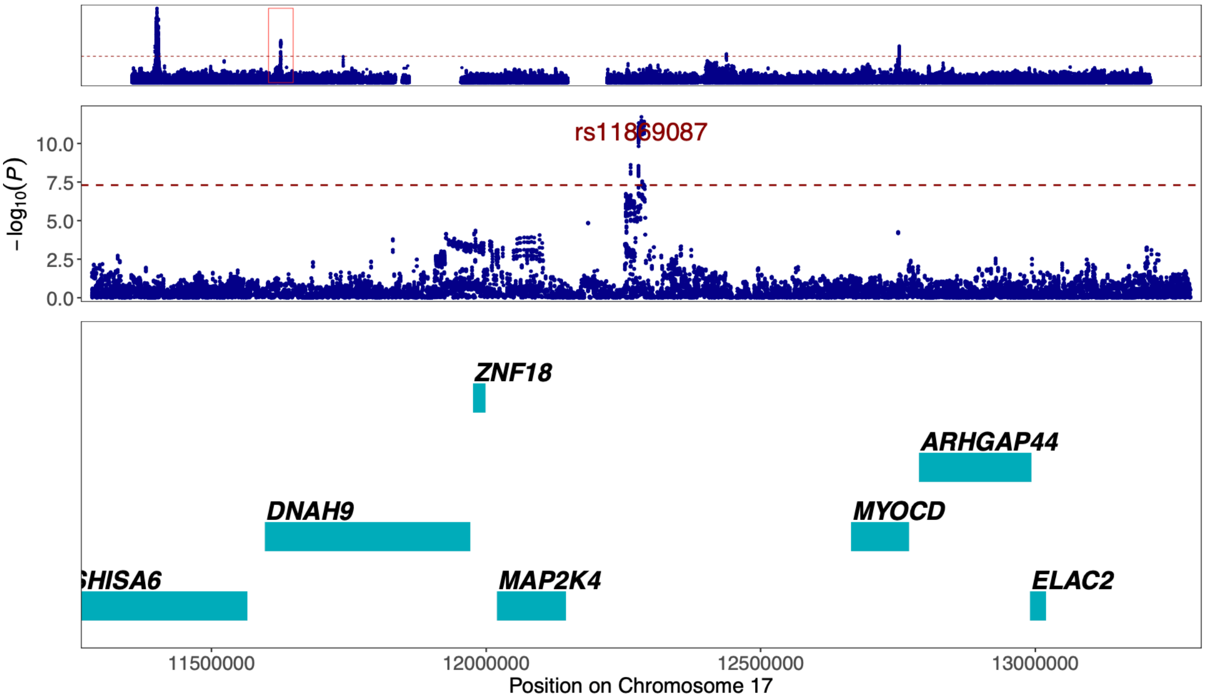


Figure 39. **Regional association plot of previously unreported TAA signal at 17p12 around the GWAS lead variant rs11869087.** The most likely causal gene explaining the association at the locus was predicted to be *MAP2K4* (*Mitogen-Activated Protein Kinase Kinase 4*). The top panel shows an overview of the genomic position, the middle and bottom panels depict individual SNVs and genes within the 2 MB locus window on chromosome 17. The middle Y-axis displays the strength of the association as -log10(*P*) with a dashed red line marking the threshold for genome-wide significance ($P < 5 \times 10^{-8}$).

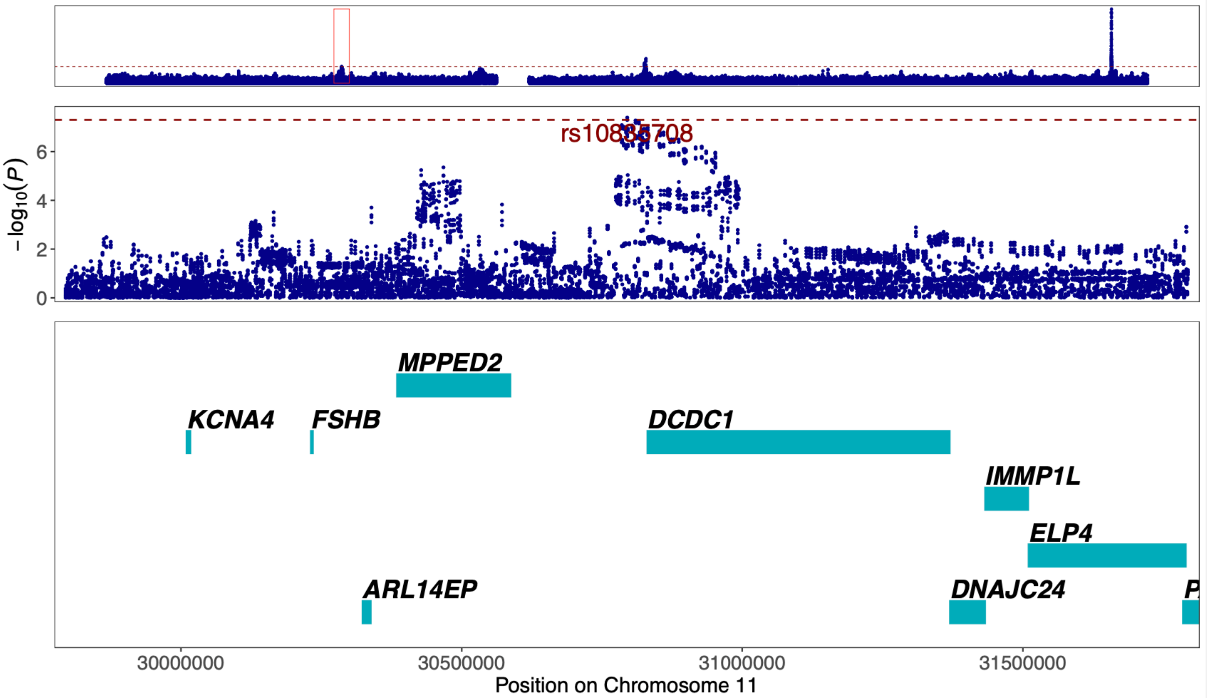


Figure 40. **Regional association plot of previously unreported TAA signal at 11p14.1 around the GWAS lead variant rs10835708.** The most likely causal gene explaining the association at the locus was predicted to be *MPPED2* (*Metallophosphoesterase Domain Containing 2*). The top panel shows an overview of the genomic position, the middle and bottom panels depict individual SNVs and genes within the 2 MB locus window on chromosome 11. The middle Y-axis displays the strength of the association as -log10(*P*) with a dashed red line marking the threshold for genome-wide significance ($P < 5 \times 10^{-8}$).

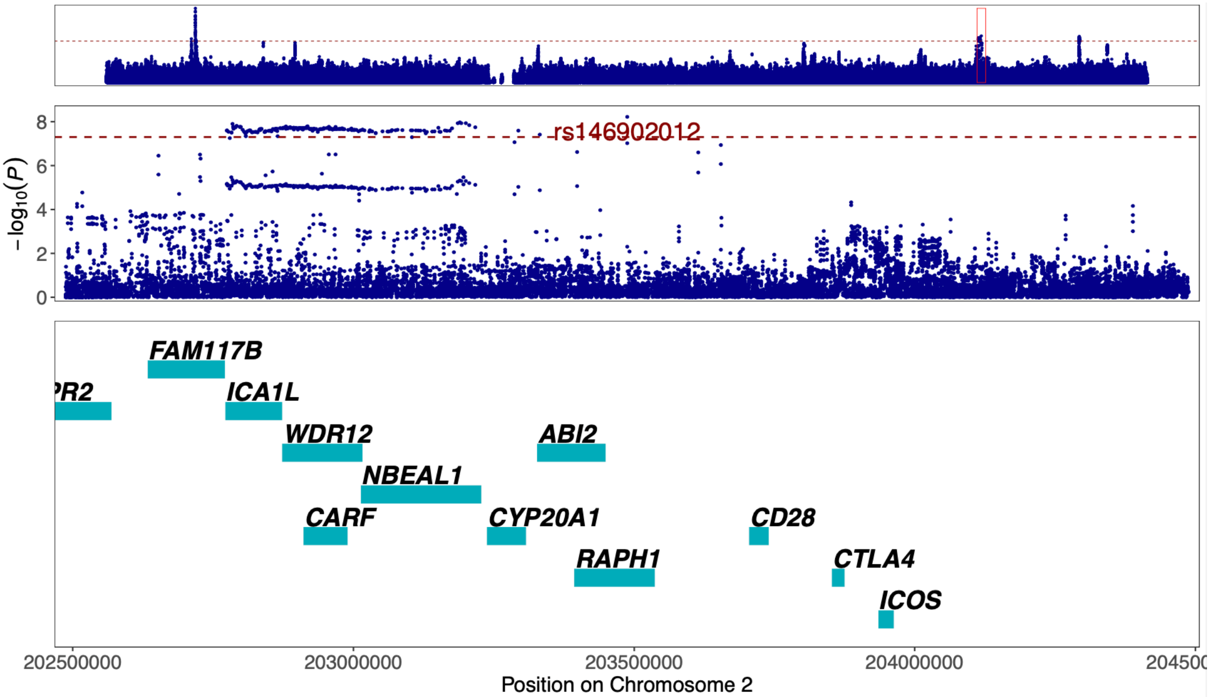


Figure 41. **Regional association plot of previously unreported AAA signal at 2q33.2 around the GWAS lead variant rs146902012.** The most likely causal gene explaining the association at the locus was predicted to be *FAM117B* (*Family With Sequence Similarity 117 Member B*). The top panel shows an overview of the genomic position, the middle and bottom panels depict individual SNVs and genes within the 2 MB locus window on chromosome 2. The middle Y-axis displays the strength of the association as -$\log_{10}$($P$) with a dashed red line marking the threshold for genome-wide significance ($P < 5 \times 10^{-8}$).

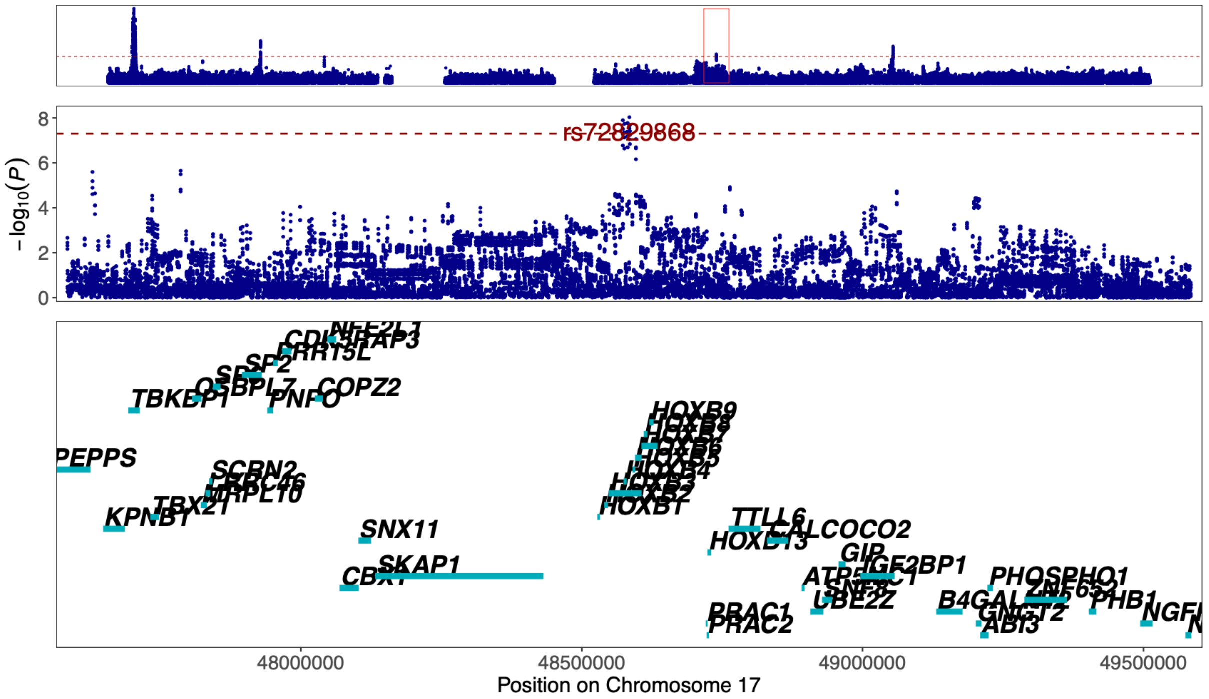


Figure 42. **Regional association plot of previously unreported TAA signal at 17q21.32 around the GWAS lead variant rs72829868.** The most likely causal gene explaining the association at the locus was predicted to be *HOXB3* (*Homeobox B3*). The top panel shows an overview of the genomic position, the middle and bottom panels depict individual SNVs and genes within the 2 MB locus window on chromosome 17. The middle Y-axis displays the strength of the association as -$\log_{10}$($P$) with a dashed red line marking the threshold for genome-wide significance ($P < 5 \times 10^{-8}$).

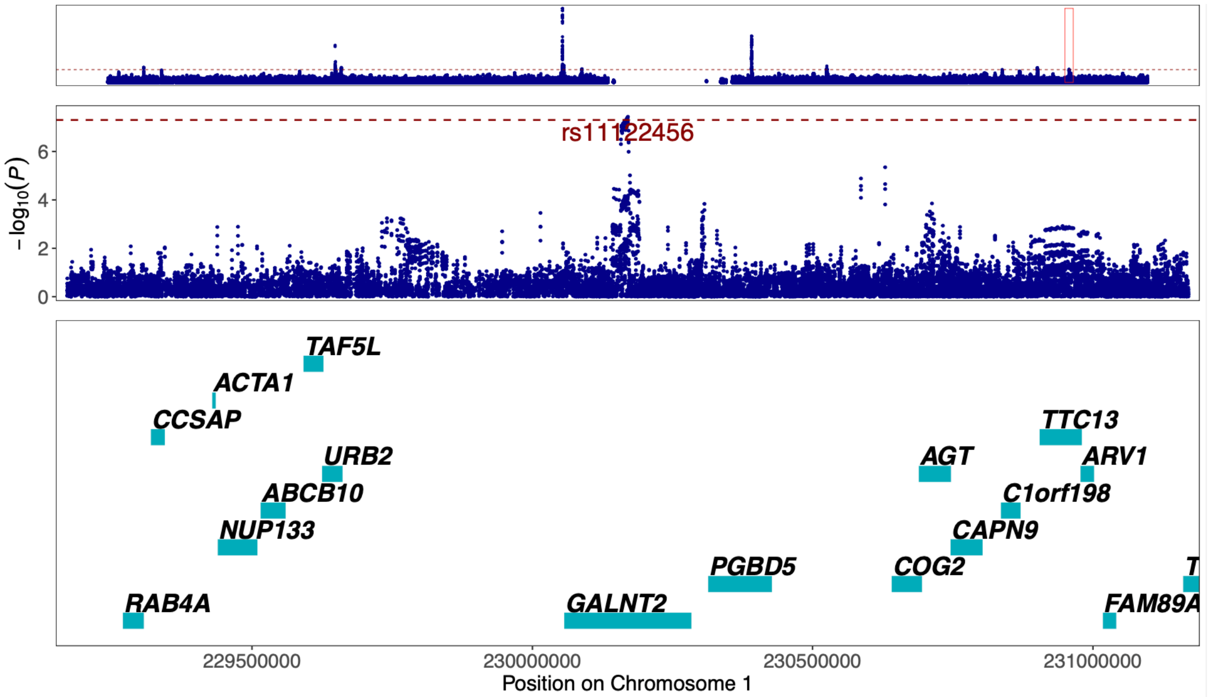


Figure 43. **Regional association plot of previously unreported AAA signal at 1q42.13 around the GWAS lead variant rs11122456.** The most likely causal gene explaining the association at the locus was predicted to be *GALNT2* (*Polypeptide N-Acetylgalactosaminyltransferase 2*). The top panel shows an overview of the genomic position, the middle and bottom panels depict individual SNVs and genes within the 2 MB locus window on chromosome 1. The middle Y-axis displays the strength of the association as -log10(*P*) with a dashed red line marking the threshold for genome-wide significance ($P < 5 \times 10^{-8}$).

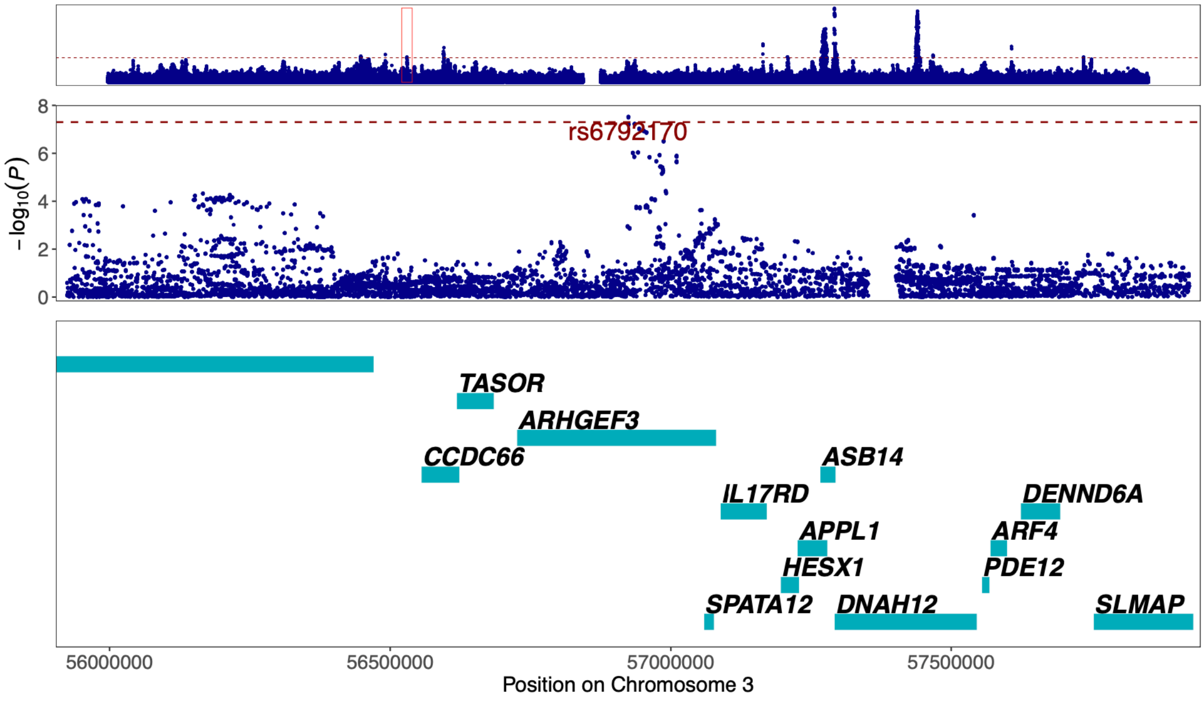


Figure 44. **Regional association plot of previously unreported CAD signal at 3p14.3 around the GWAS lead variant rs6792170.** The most likely causal gene explaining the association at the locus was predicted to be *ARHGEF3* (*Rho Guanine Nucleotide Exchange Factor 3*). The top panel shows an overview of the genomic position, the middle and bottom panels depict individual SNVs and genes within the 2 MB locus window on chromosome 3. The middle Y-axis displays the strength of the association as -log10(*P*) with a dashed red line marking the threshold for genome-wide significance ($P < 5 \times 10^{-8}$).

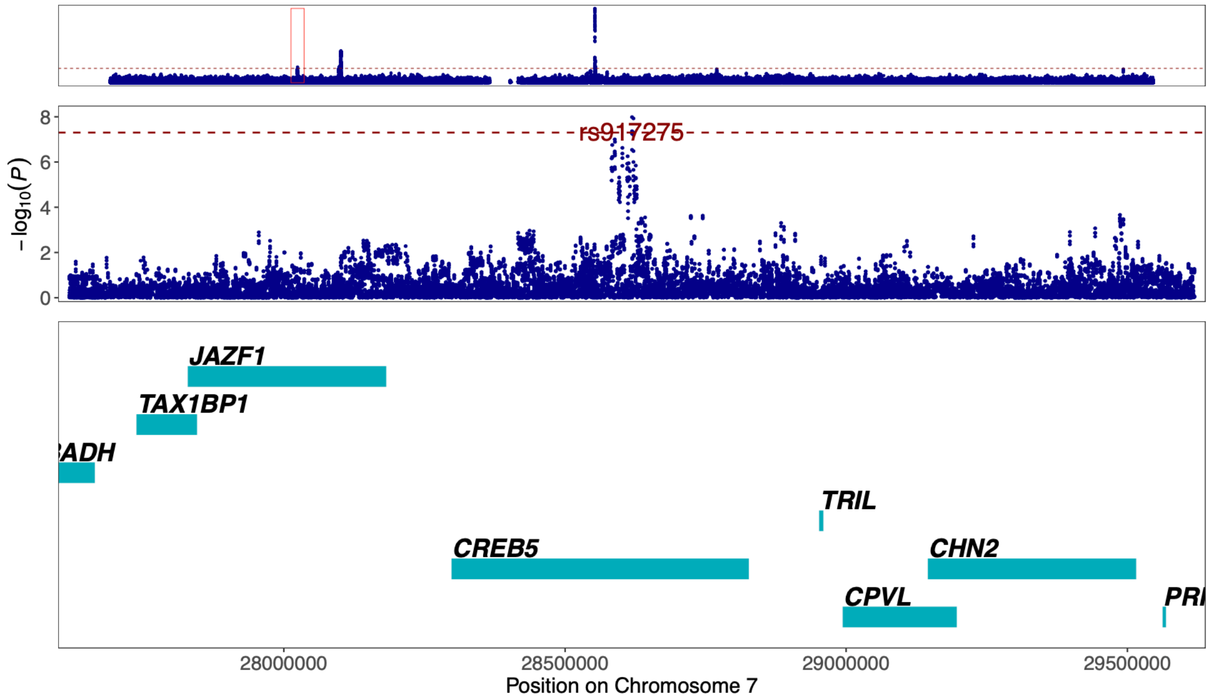


Figure 45. **Regional association plot of previously unreported TAA signal at 7p15.1 around the GWAS lead variant rs917275.** The most likely causal gene explaining the association at the locus was predicted to be *CREB5* ( *CAMP Responsive Element Binding Protein 5*). The top panel shows an overview of the genomic position, the middle and bottom panels depict individual SNVs and genes within the 2 MB locus window on chromosome 7. The middle Y-axis displays the strength of the association as -$\log_{10}$($P$) with a dashed red line marking the threshold for genome-wide significance ($P < 5 \times 10^{-8}$).

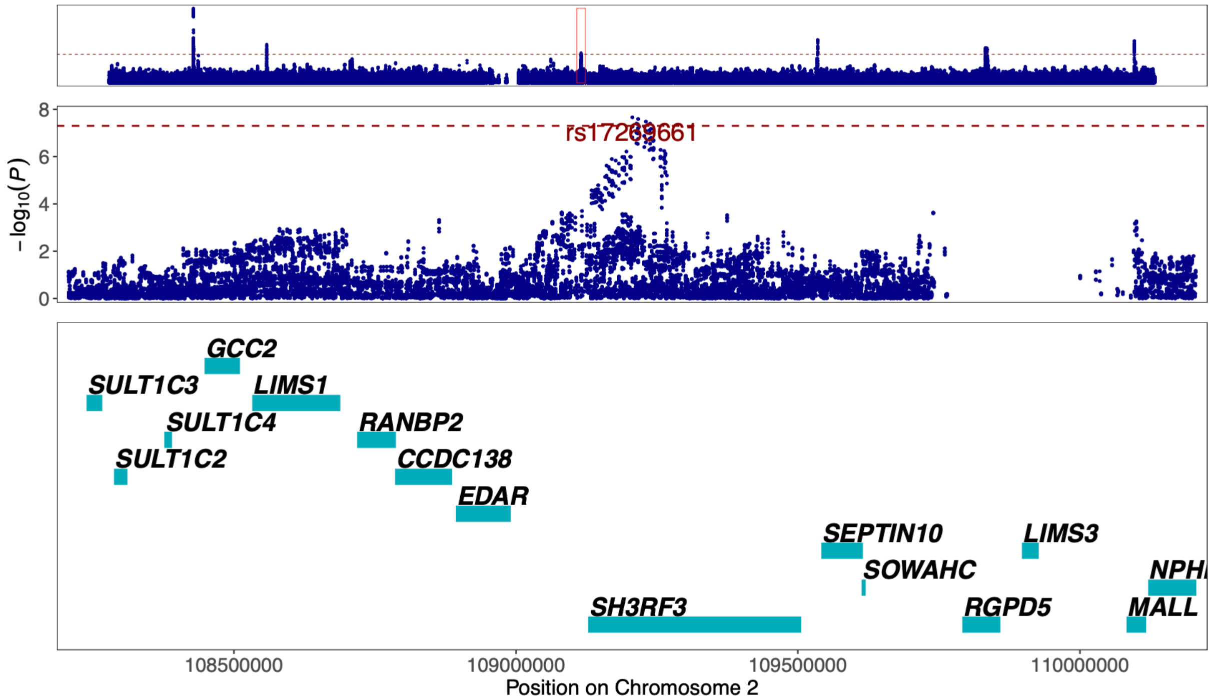


Figure 46. **Regional association plot of previously unreported TAA signal at 2q13 around the GWAS lead variant rs17269661.** The most likely causal gene explaining the association at the locus was predicted to be *SH3RF3* (*SH3 Domain Containing Ring Finger 3*). The top panel shows an overview of the genomic position, the middle and bottom panels depict individual SNVs and genes within the 2 MB locus window on chromosome 2. The middle Y-axis displays the strength of the association as -$\log_{10}$($P$) with a dashed red line marking the threshold for genome-wide significance ($P < 5 \times 10^{-8}$).

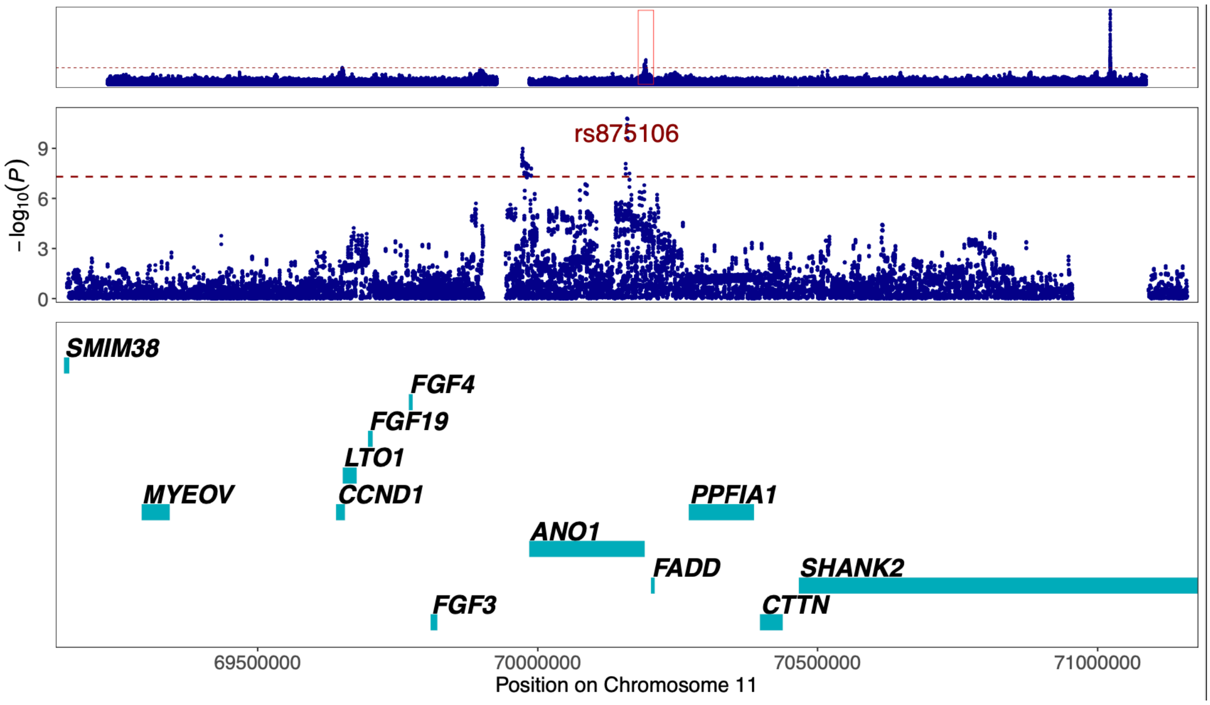


Figure 47. **Regional association plot of previously unreported TAA signal at 11q13.3 around the GWAS lead variant rs875106.** The most likely causal gene explaining the association at the locus was predicted to be *ANO1* (*Anoctamin 1*). The top panel shows an overview of the genomic position, the middle and bottom panels depict individual SNVs and genes within the 2 MB locus window on chromosome 11. The middle Y-axis displays the strength of the association as $-\log_{10}(P)$ with a dashed red line marking the threshold for genome-wide significance ($P < 5 \times 10^{-8}$).

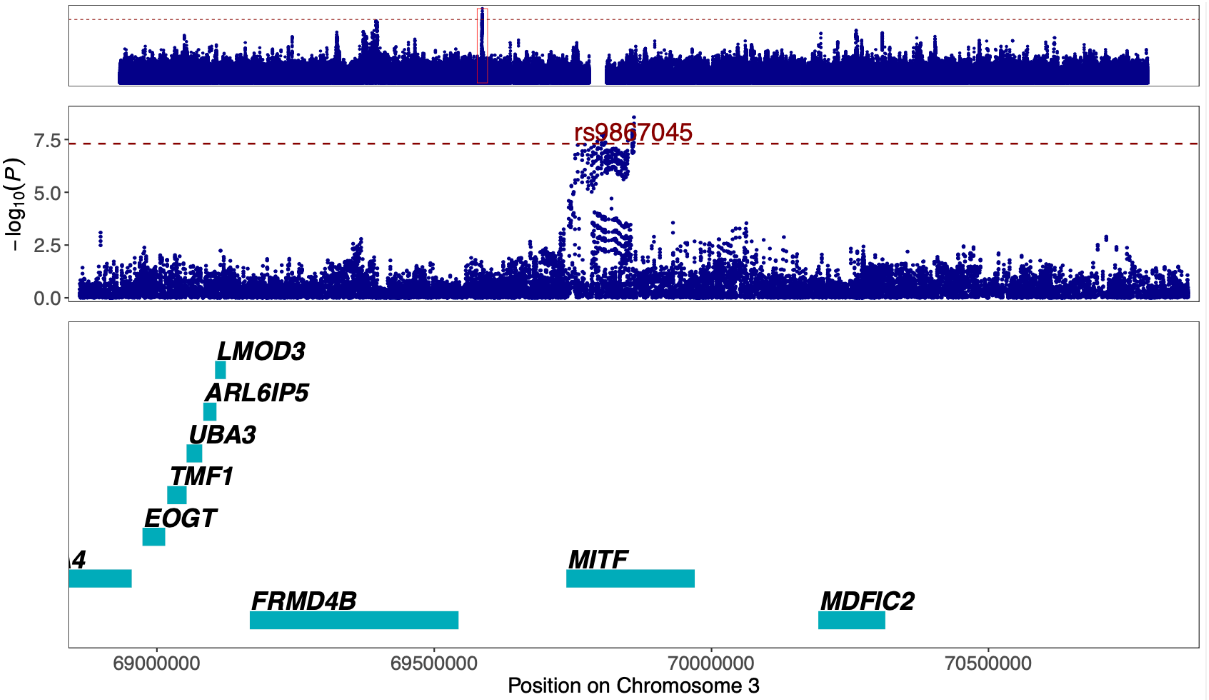


Figure 48. **Regional association plot of previously unreported AAA signal at 3p14.1 around the GWAS lead variant rs9867045.** The most likely causal gene explaining the association at the locus was predicted to be *MITF* (*Melanocyte Inducing Transcription Factor*). The top panel shows an overview of the genomic position, the middle and bottom panels depict individual SNVs and genes within the 2 MB locus window on chromosome 3. The middle Y-axis displays the strength of the association as $-\log_{10}(P)$ with a dashed red line marking the threshold for genome-wide significance ($P < 5 \times 10^{-8}$).

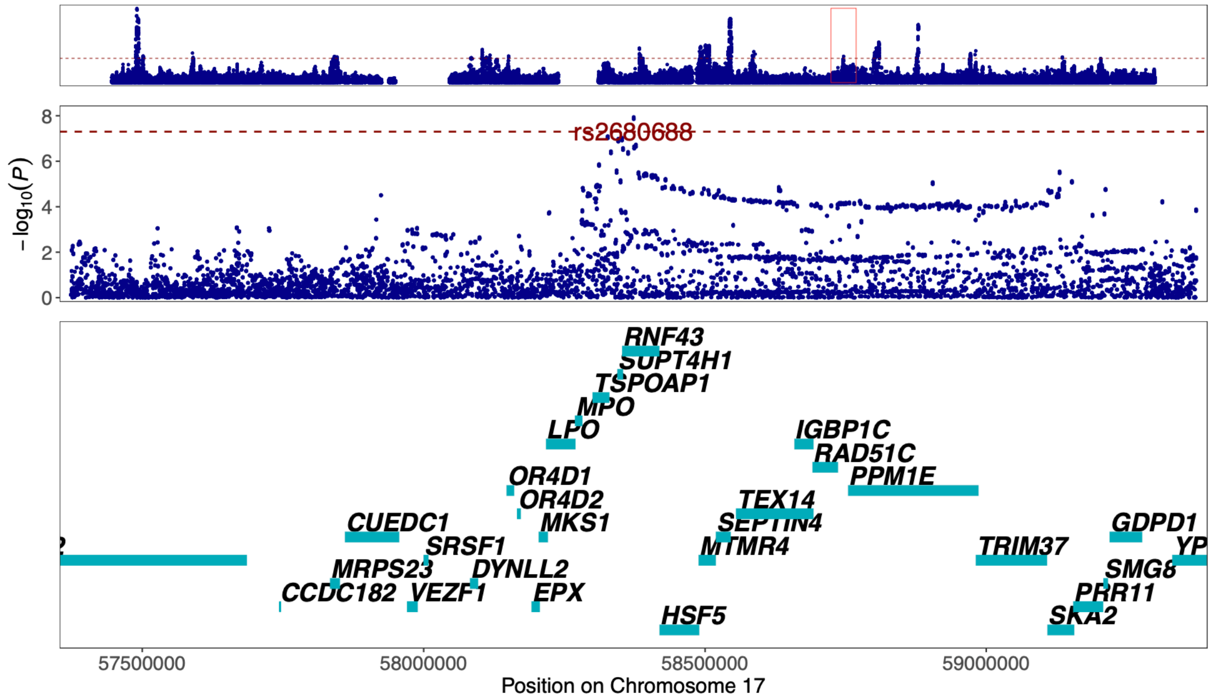


Figure 49. **Regional association plot of previously unreported CAD signal at 17q22 around the GWAS lead variant rs2680688.** The most likely causal gene explaining the association at the locus was predicted to be *MPO* (*Myeloperoxidase*). The top panel shows an overview of the genomic position, the middle and bottom panels depict individual SNVs and genes within the 2 MB locus window on chromosome 17. The middle Y-axis displays the strength of the association as -log10(*P*) with a dashed red line marking the threshold for genome-wide significance ($P < 5 \times 10^{-8}$).

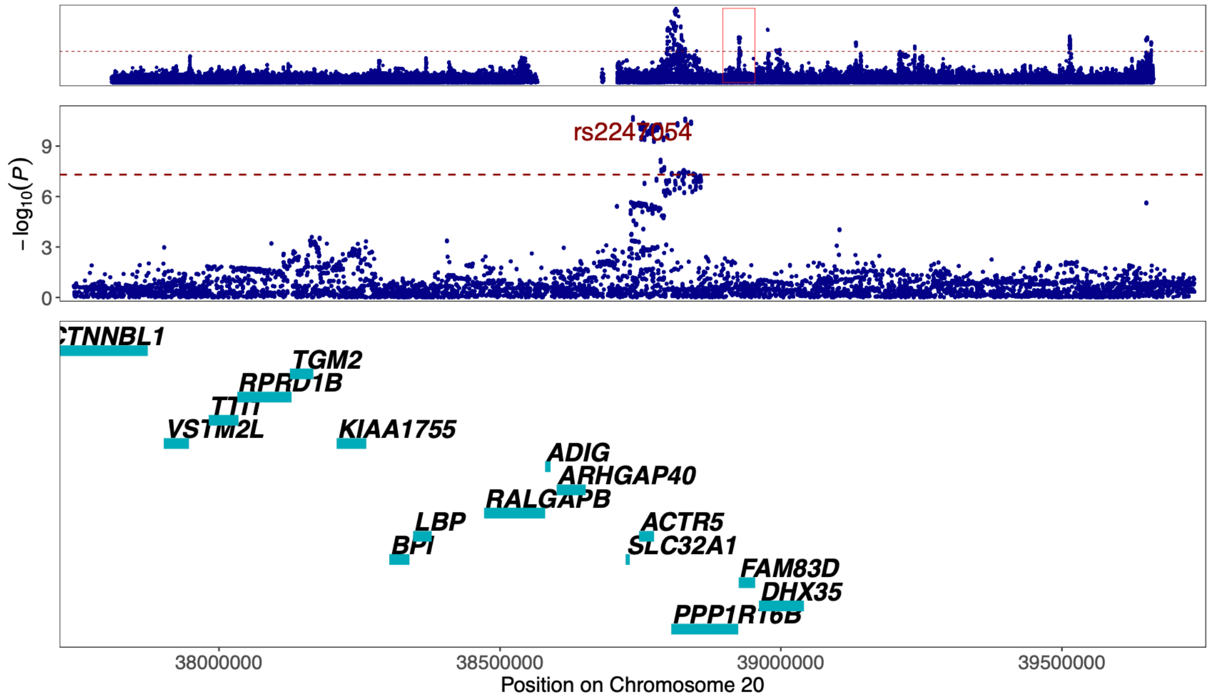


Figure 50. **Regional association plot of previously unreported CAD signal at 20q11.23 around the GWAS lead variant rs2247054.** The most likely causal gene explaining the association at the locus was predicted to be *PPP1R16B* (*Protein Phosphatase 1 Regulatory Subunit 16B*). The top panel shows an overview of the genomic position, the middle and bottom panels depict individual SNVs and genes within the 2 MB locus window on chromosome 20. The middle Y-axis displays the strength of the association as -log10(*P*) with a dashed red line marking the threshold for genome-wide significance ($P < 5 \times 10^{-8}$).

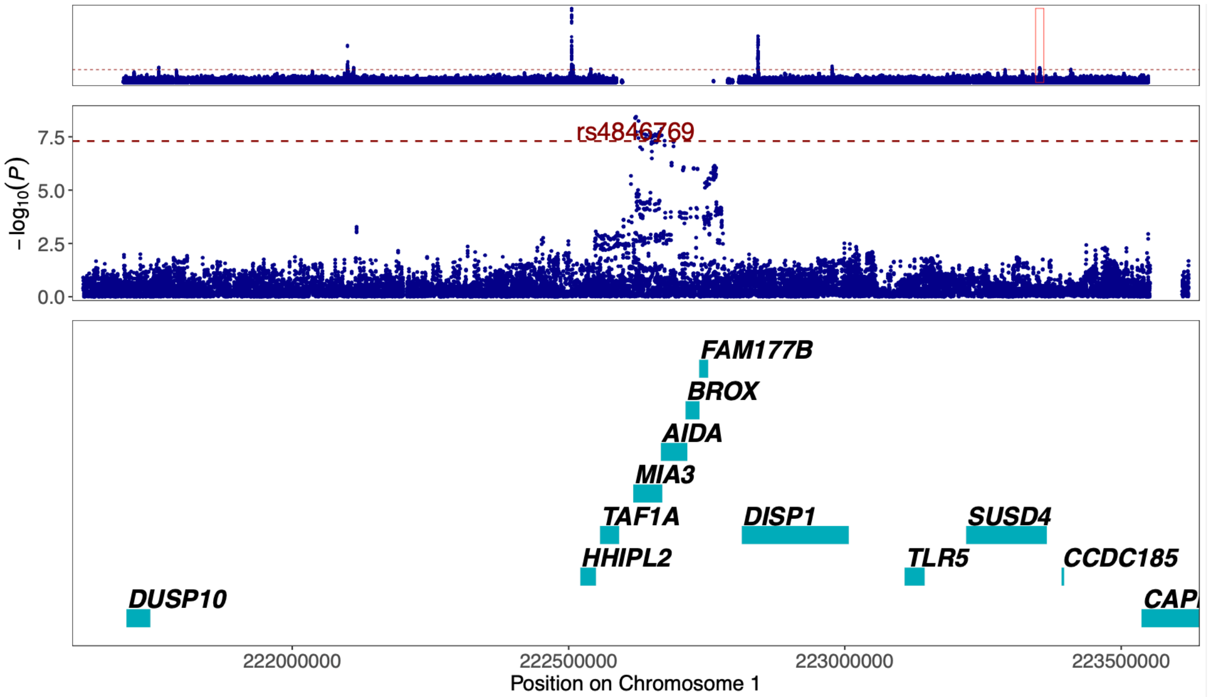


Figure 51. **Regional association plot of previously unreported AAA signal at 1q41 around the GWAS lead variant rs4846769.** The most likely causal gene explaining the association at the locus was predicted to be *MIA3* (*MIA SH3 Domain ER Export Factor 3*). The top panel shows an overview of the genomic position, the middle and bottom panels depict individual SNVs and genes within the 2 MB locus window on chromosome 1. The middle Y-axis displays the strength of the association as -log$_{10}$(*P*) with a dashed red line marking the threshold for genome-wide significance ($P < 5 \times 10^{-8}$).

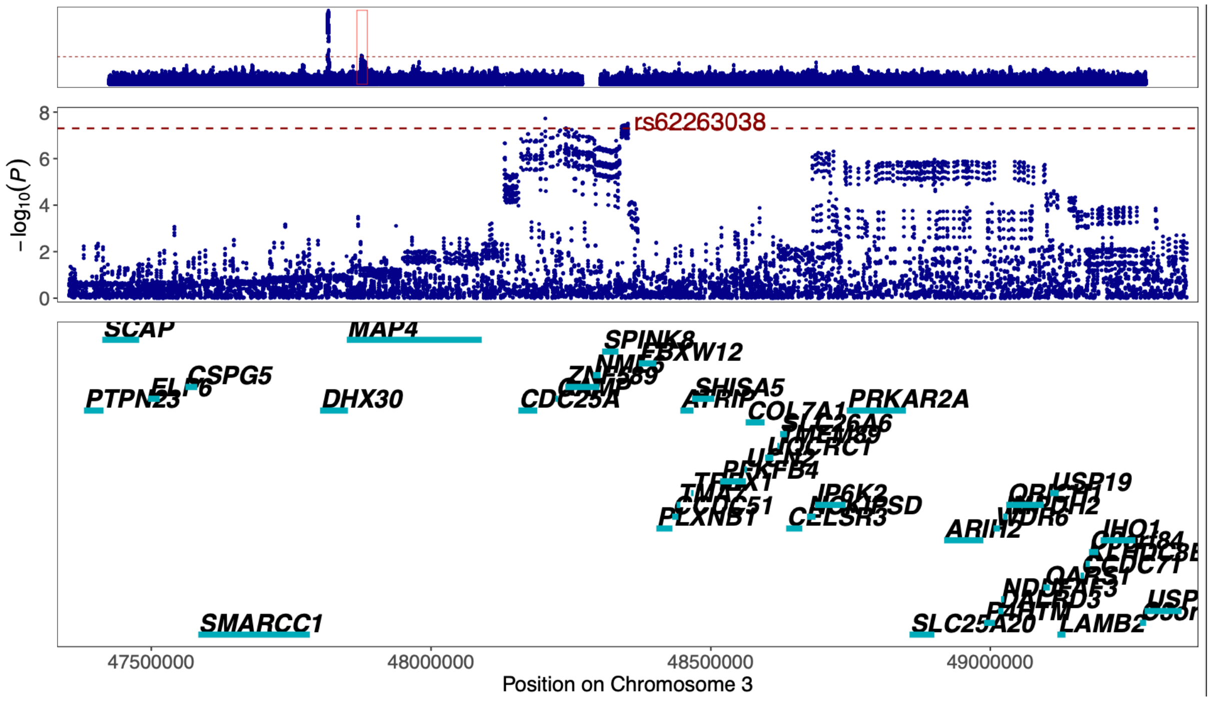


Figure 52. **Regional association plot of previously unreported TAA signal at 3p21.31 around the GWAS lead variant rs62263038.** The most likely causal gene explaining the association at the locus was predicted to be *ZNF589* (*Zinc Finger Protein 589*). The top panel shows an overview of the genomic position, the middle and bottom panels depict individual SNVs and genes within the 2 MB locus window on chromosome 3. The middle Y-axis displays the strength of the association as -log$_{10}$(*P*) with a dashed red line marking the threshold for genome-wide significance ($P < 5 \times 10^{-8}$).

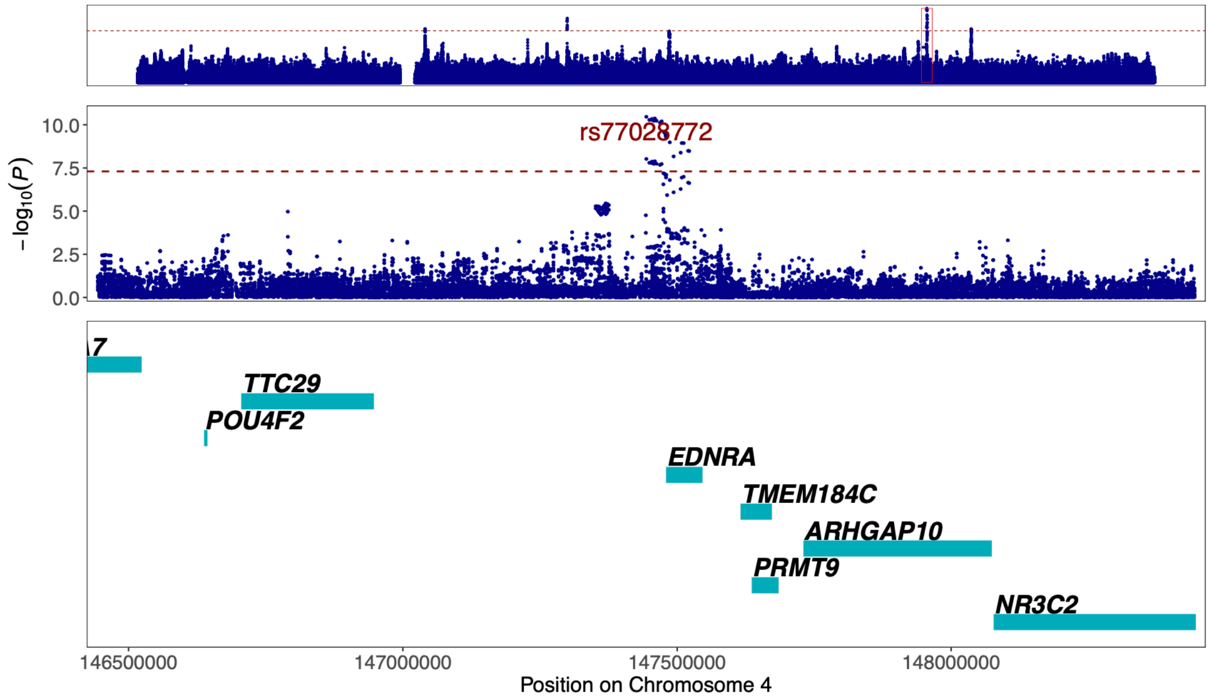


Figure 53. **Regional association plot of previously unreported AAA signal at 4q31.22 around the GWAS lead variant rs77028772.** The most likely causal gene explaining the association at the locus was predicted to be *EDNRA* (*Endothelin Receptor Type A*). The top panel shows an overview of the genomic position, the middle and bottom panels depict individual SNVs and genes within the 2 MB locus window on chromosome 4. The middle Y-axis displays the strength of the association as -log$_{10}$(*P*) with a dashed red line marking the threshold for genome-wide significance ($P < 5 \times 10^{-8}$).

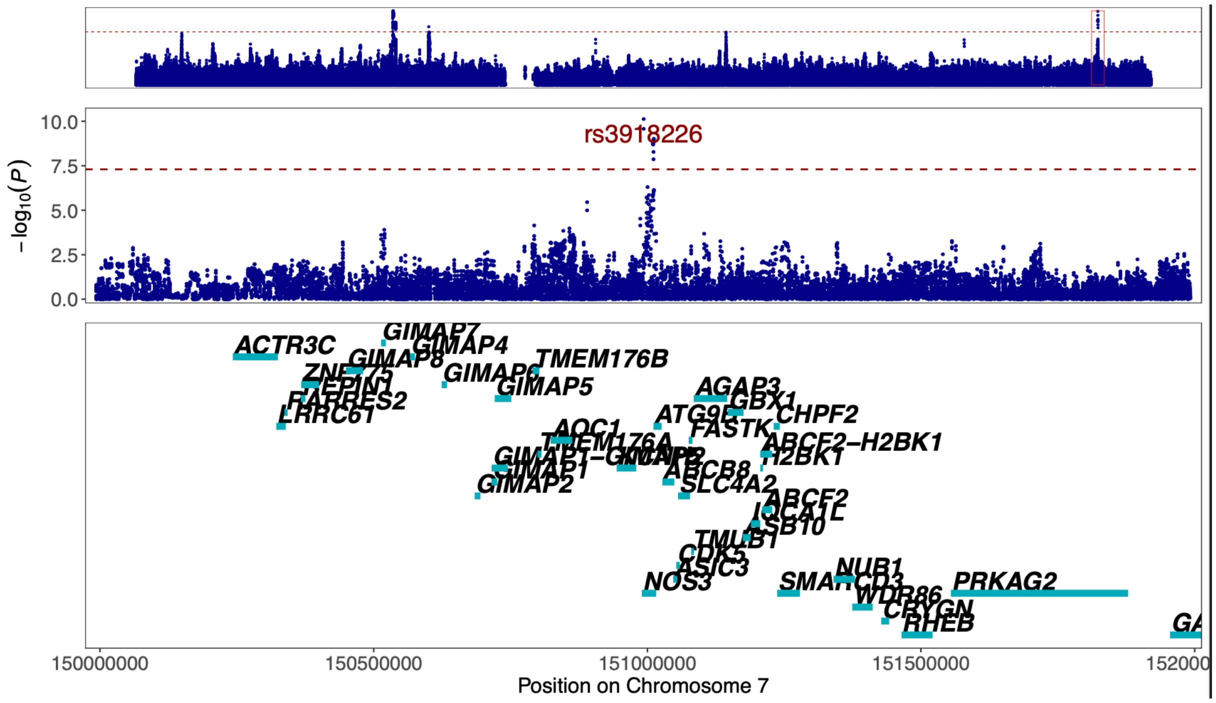


Figure 54. **Regional association plot of previously unreported AAA signal at 7q36.1 around the GWAS lead variant rs3918226.** The most likely causal gene explaining the association at the locus was predicted to be *NOS3* (*Nitric Oxide Synthase 3*). The top panel shows an overview of the genomic position, the middle and bottom panels depict individual SNVs and genes within the 2 MB locus window on chromosome 7. The middle Y-axis displays the strength of the association as -log$_{10}$(*P*) with a dashed red line marking the threshold for genome-wide significance ($P < 5 \times 10^{-8}$).

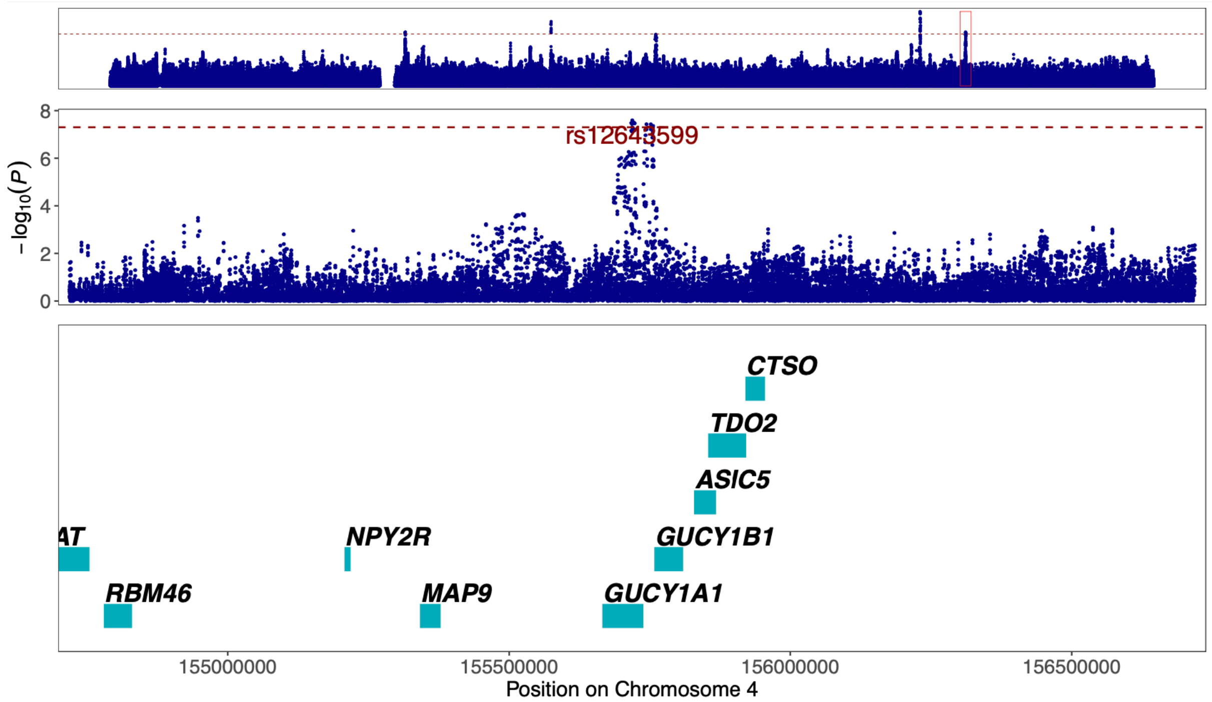


Figure 55. **Regional association plot of previously unreported AAA signal at 4q32.1 around the GWAS lead variant rs12643599.** The most likely causal gene explaining the association at the locus was predicted to be *GUCY1A1* (*Guanylate Cyclase 1 Soluble Subunit Alpha 1*). The top panel shows an overview of the genomic position, the middle and bottom panels depict individual SNVs and genes within the 2 MB locus window on chromosome 4. The middle Y-axis displays the strength of the association as -$\log_{10}(P)$ with a dashed red line marking the threshold for genome-wide significance ($P < 5 \times 10^{-8}$).

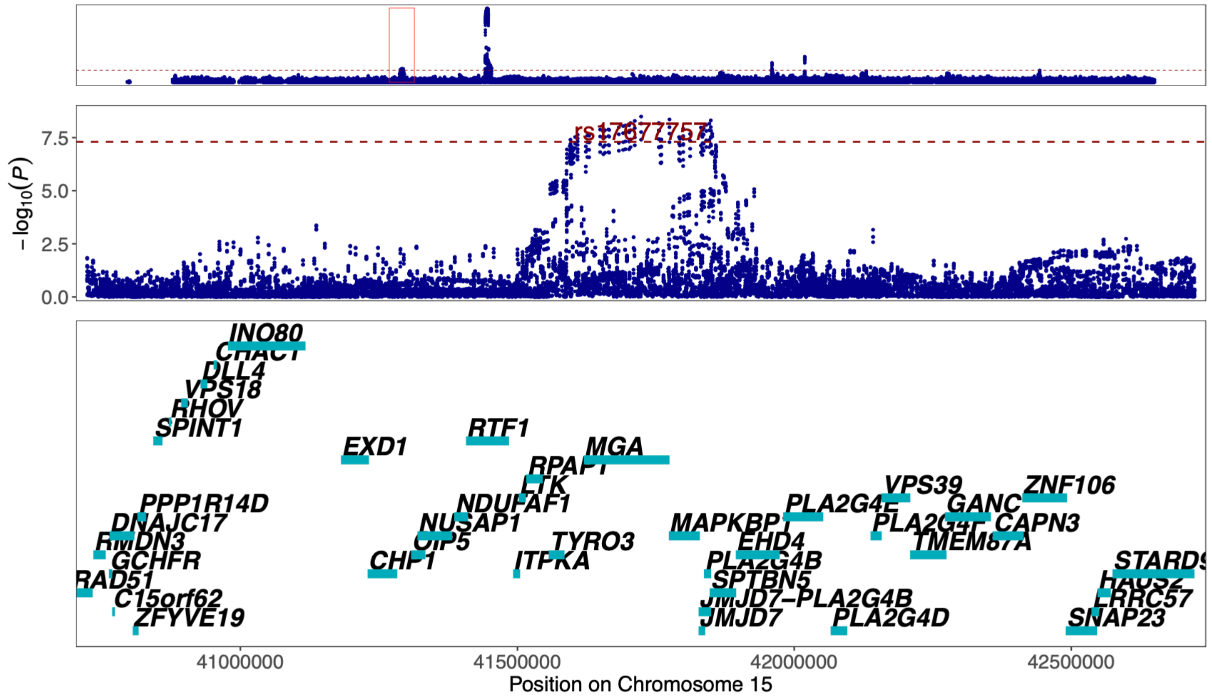


Figure 56. **Regional association plot of previously unreported TAA signal at 15q15.1 around the GWAS lead variant rs17677757.** The most likely causal gene explaining the association at the locus was predicted to be *PLA2G4B* (*Phospholipase A2 Group IVB*). The top panel shows an overview of the genomic position, the middle and bottom panels depict individual SNVs and genes within the 2 MB locus window on chromosome 15. The middle Y-axis displays the strength of the association as -$\log_{10}(P)$ with a dashed red line marking the threshold for genome-wide significance ($P < 5 \times 10^{-8}$).

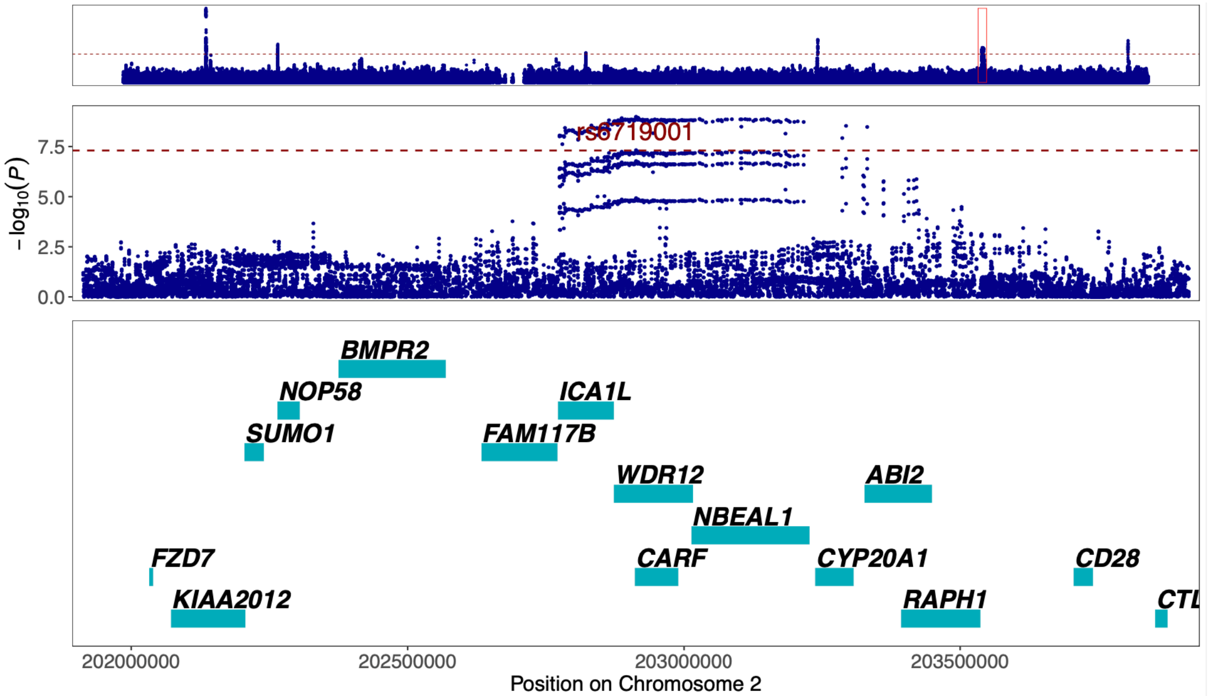


Figure 57. **Regional association plot of previously unreported TAA signal at 2q33.2 around the GWAS lead variant rs6719001.** The most likely causal gene explaining the association at the locus was predicted to be *CARF* (*Calcium Responsive Transcription Factor*). The top panel shows an overview of the genomic position, the middle and bottom panels depict individual SNVs and genes within the 2 MB locus window on chromosome 2. The middle Y-axis displays the strength of the association as -log$_{10}$($P$) with a dashed red line marking the threshold for genome-wide significance ($P < 5$ x $10^{-8}$).

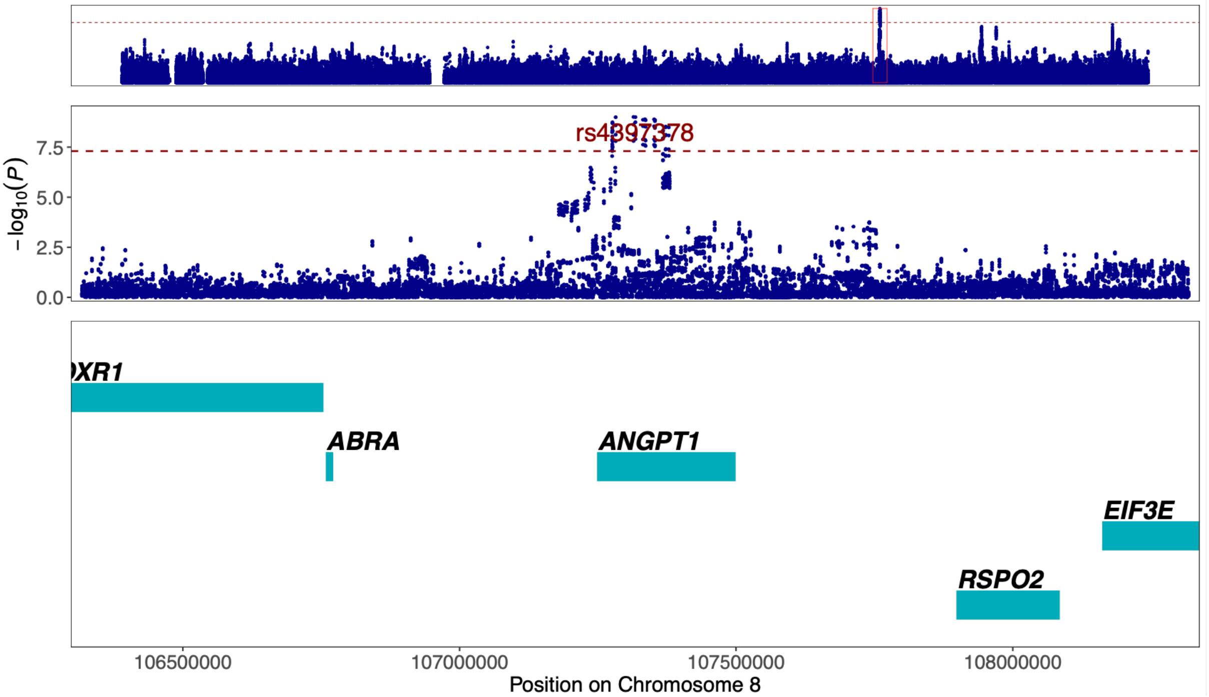


Figure 58. **Regional association plot of previously unreported TAA signal at 8q23.1 around the GWAS lead variant rs4397378.** The most likely causal gene explaining the association at the locus was predicted to be *ANGPT1* (*Angiopoietin 1*). The top panel shows an overview of the genomic position, the middle and bottom panels depict individual SNVs and genes within the 2 MB locus window on chromosome 8. The middle Y-axis displays the strength of the association as -log$_{10}$($P$) with a dashed red line marking the threshold for genome-wide significance ($P < 5$ x $10^{-8}$).

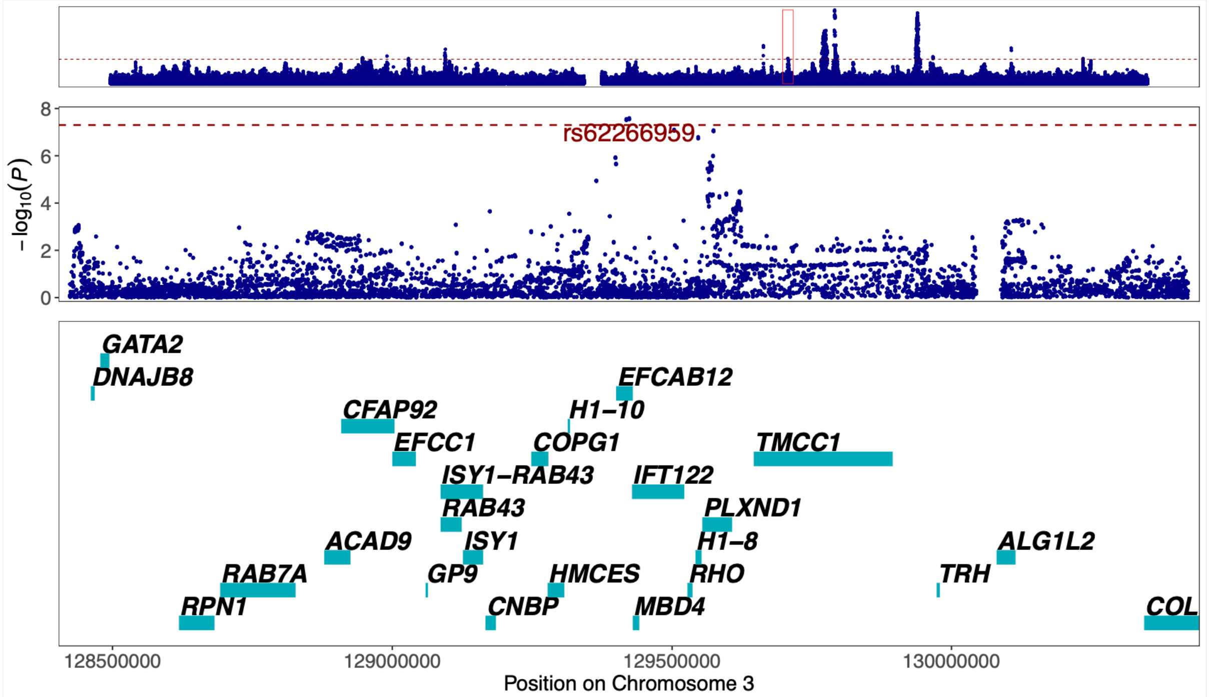


Figure 59. **Regional association plot of previously unreported CAD signal at 3q21.3 around the GWAS lead variant rs62266959.** The most likely causal gene explaining the association at the locus was predicted to be *IFT122* (*Intraflagellar Transport 122*). The top panel shows an overview of the genomic position, the middle and bottom panels depict individual SNVs and genes within the 2 MB locus window on chromosome 3. The middle Y-axis displays the strength of the association as -log$_{10}$($P$) with a dashed red line marking the threshold for genome-wide significance ($P < 5 \times 10^{-8}$).

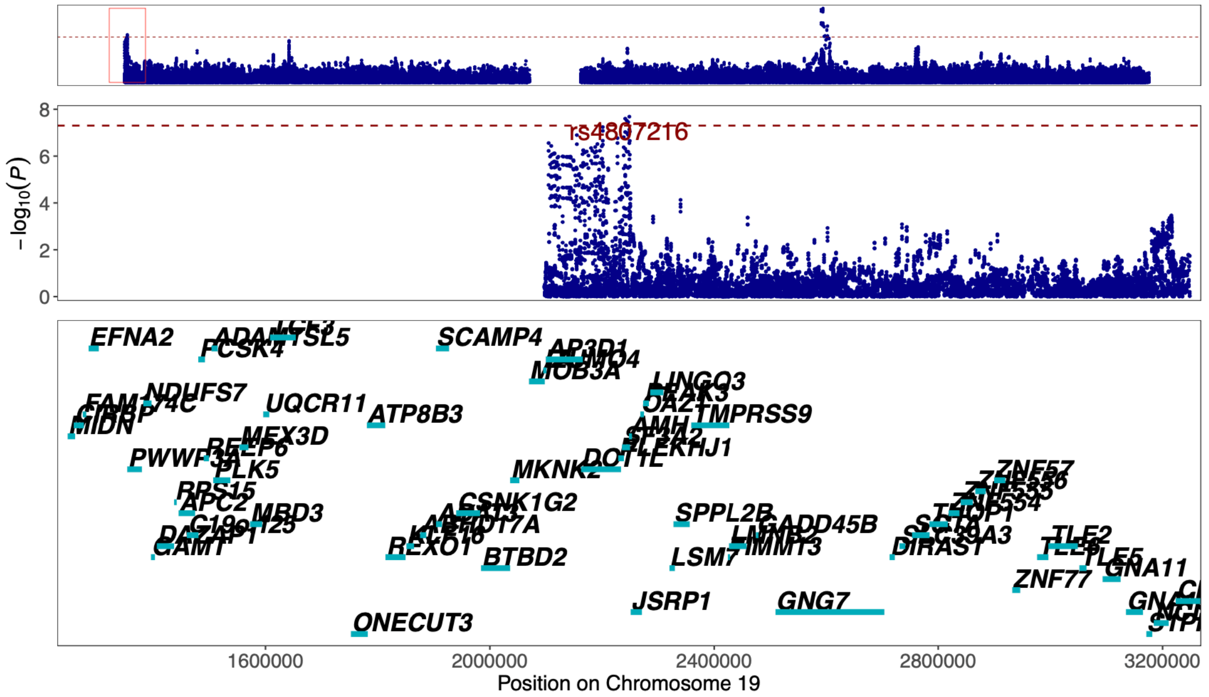


Figure 60. **Regional association plot of previously unreported TAA signal at 19p13.3 around the GWAS lead variant rs4807216.** The most likely causal gene explaining the association at the locus was predicted to be *AMH* (*Anti-Mullerian Hormone*). The top panel shows an overview of the genomic position, the middle and bottom panels depict individual SNVs and genes within the 2 MB locus window on chromosome 19. The middle Y-axis displays the strength of the association as -log$_{10}$($P$) with a dashed red line marking the threshold for genome-wide significance ($P < 5 \times 10^{-8}$).

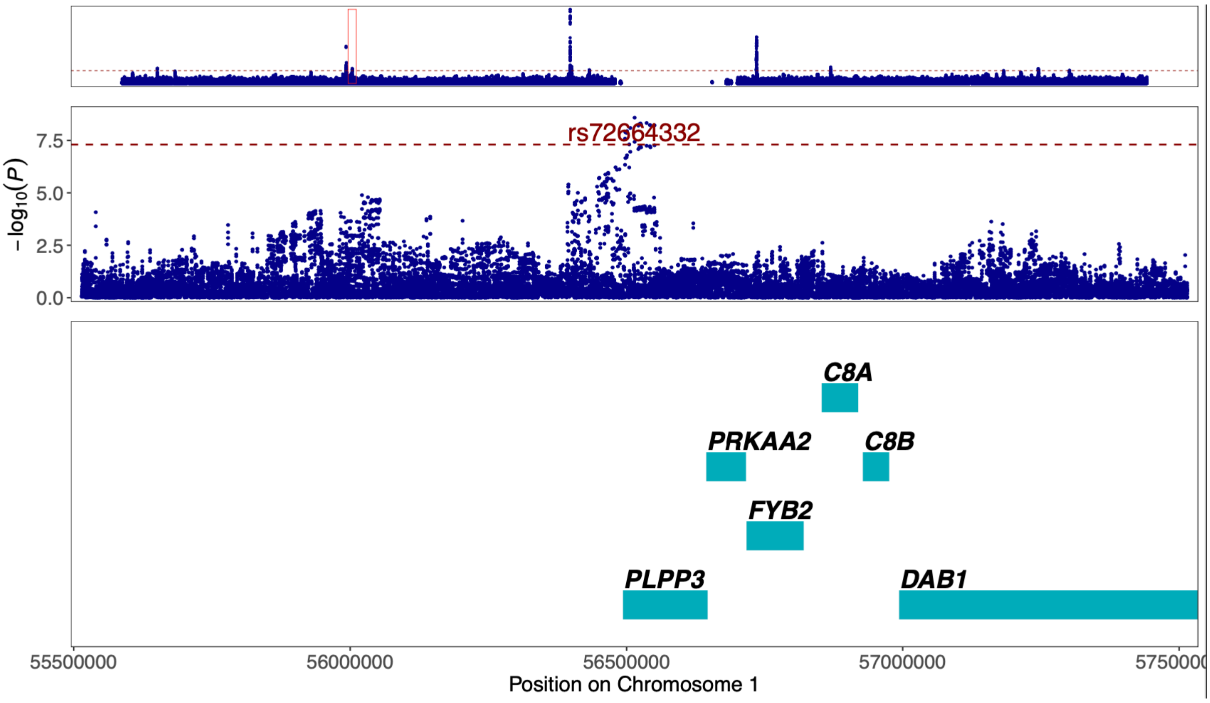


Figure 61. **Regional association plot of previously unreported AAA signal at 1p32.2 around the GWAS lead variant rs72664332.** The most likely causal gene explaining the association at the locus was predicted to be *PLPP3* (*Phospholipid Phosphatase 3*). The top panel shows an overview of the genomic position, the middle and bottom panels depict individual SNVs and genes within the 2 MB locus window on chromosome 1. The middle Y-axis displays the strength of the association as -$\log_{10}(P)$ with a dashed red line marking the threshold for genome-wide significance ($P < 5 \times 10^{-8}$).

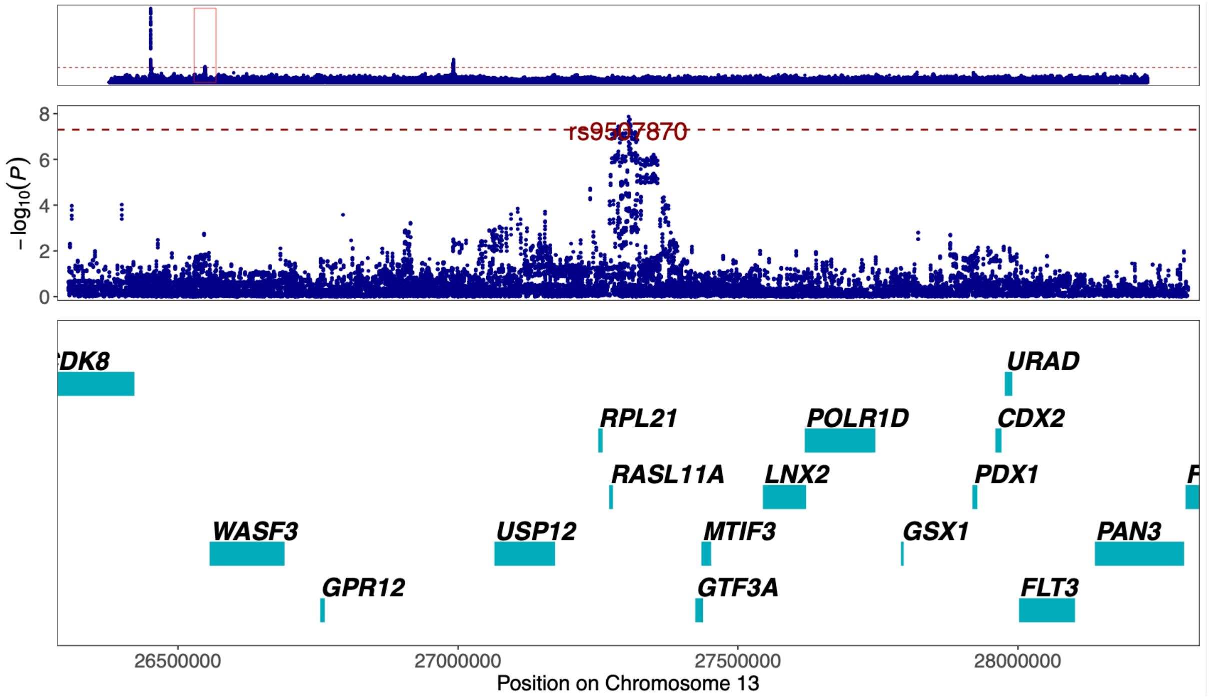


Figure 62. **Regional association plot of previously unreported TAA signal at 13q12.2 around the GWAS lead variant rs9507870.** The most likely causal gene explaining the association at the locus was predicted to be *RASL11A* (*RAS Like Family 11 Member A*). The top panel shows an overview of the genomic position, the middle and bottom panels depict individual SNVs and genes within the 2 MB locus window on chromosome 13. The middle Y-axis displays the strength of the association as -$\log_{10}(P)$ with a dashed red line marking the threshold for genome-wide significance ($P < 5 \times 10^{-8}$).

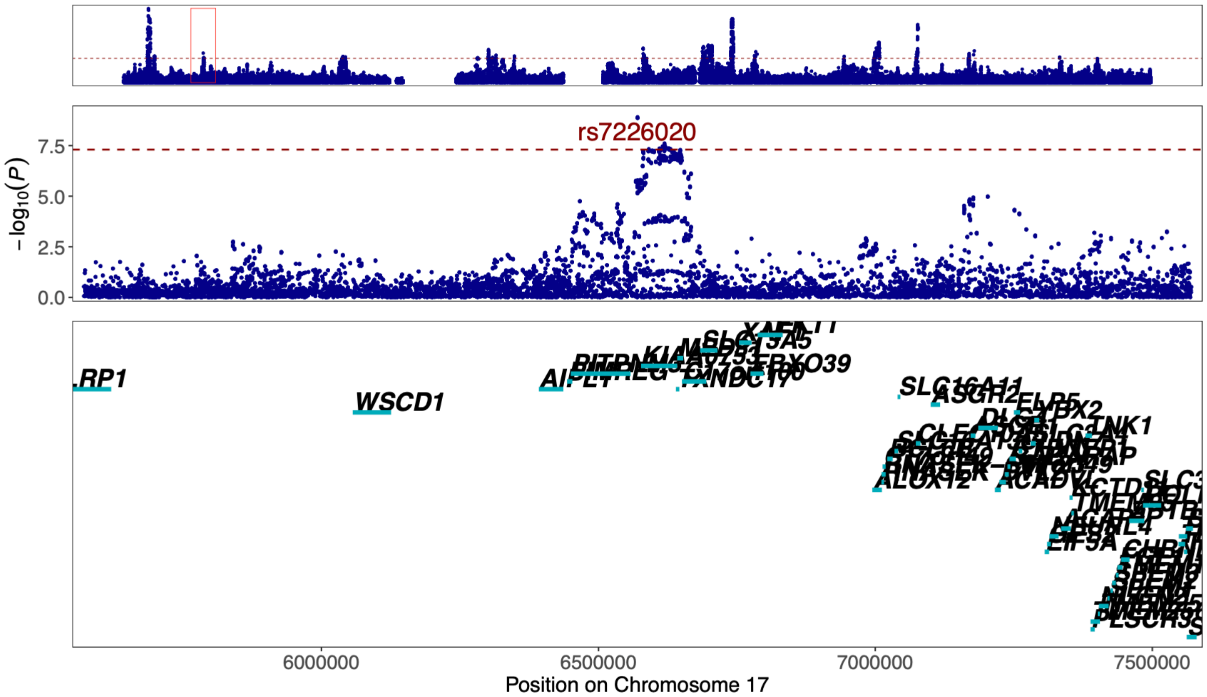


Figure 63. **Regional association plot of previously unreported CAD signal at 17p13.1 around the GWAS lead variant rs7226020.** The most likely causal gene explaining the association at the locus was predicted to be *KIAA0753* (*KIAA0753*). The top panel shows an overview of the genomic position, the middle and bottom panels depict individual SNVs and genes within the 2 MB locus window on chromosome 17. The middle Y-axis displays the strength of the association as -$\log_{10}$(*P*) with a dashed red line marking the threshold for genome-wide significance ($P < 5 \times 10^{-8}$).

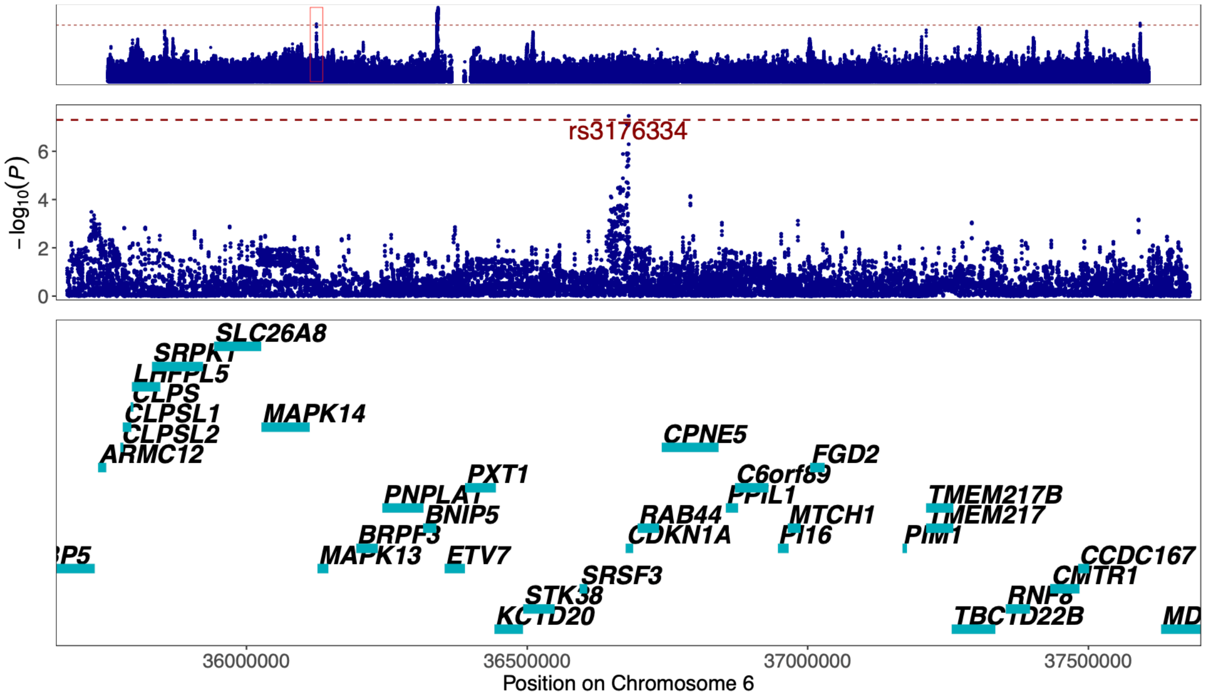


Figure 64. **Regional association plot of previously unreported TAA signal at 6p21.2 around the GWAS lead variant rs3176334.** The most likely causal gene explaining the association at the locus was predicted to be *CDKN1A* (*Cyclin Dependent Kinase Inhibitor 1A*). The top panel shows an overview of the genomic position, the middle and bottom panels depict individual SNVs and genes within the 2 MB locus window on chromosome 6. The middle Y-axis displays the strength of the association as -$\log_{10}$(*P*) with a dashed red line marking the threshold for genome-wide significance ($P < 5 \times 10^{-8}$).

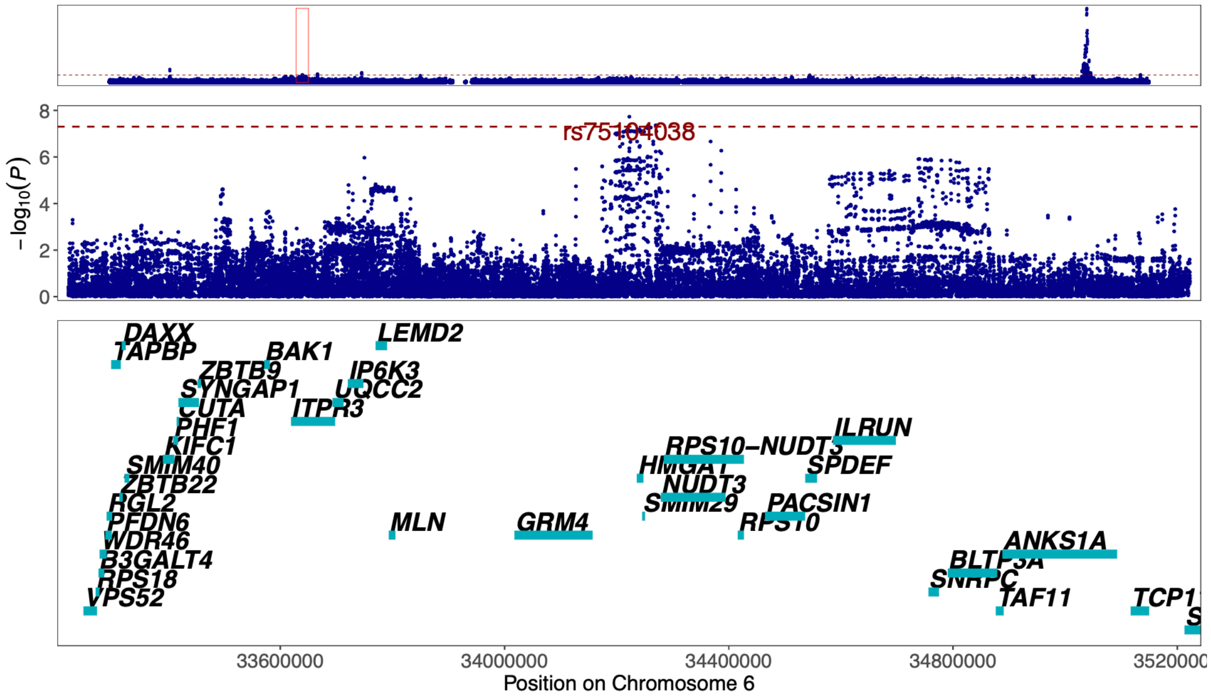


Figure 65. **Regional association plot of previously unreported AAA signal at 6p21.31 around the GWAS lead variant rs75104038.** The most likely causal gene explaining the association at the locus was predicted to be *HMGA1* (*High Mobility Group AT-Hook 1*). The top panel shows an overview of the genomic position, the middle and bottom panels depict individual SNVs and genes within the 2 MB locus window on chromosome 6. The middle Y-axis displays the strength of the association as -log10($P$) with a dashed red line marking the threshold for genome-wide significance ($P < 5 \times 10^{-8}$).

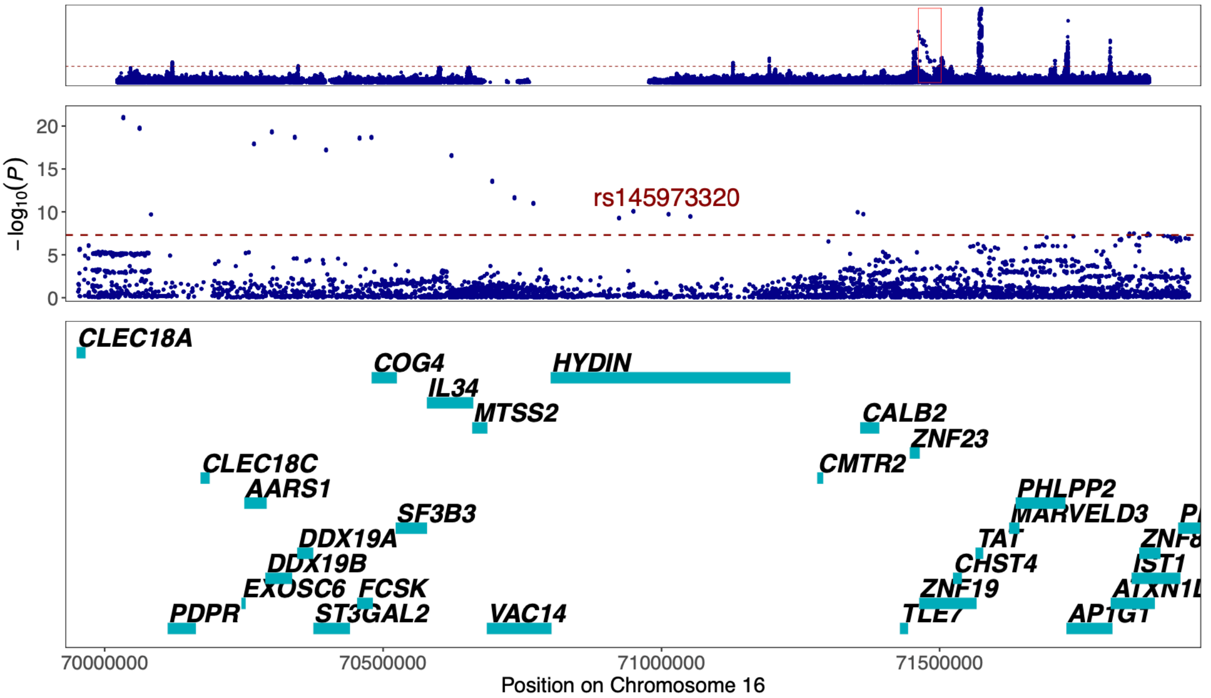


Figure 66. **Regional association plot of previously unreported CAD signal at 16q22.2 around the GWAS lead variant rs145973320.** The most likely causal gene explaining the association at the locus was predicted to be *CLEC18A* (*C-Type Lectin Domain Family 18 Member A*). The top panel shows an overview of the genomic position, the middle and bottom panels depict individual SNVs and genes within the 2 MB locus window on chromosome 16. The middle Y-axis displays the strength of the association as -log10($P$) with a dashed red line marking the threshold for genome-wide significance ($P < 5 \times 10^{-8}$).

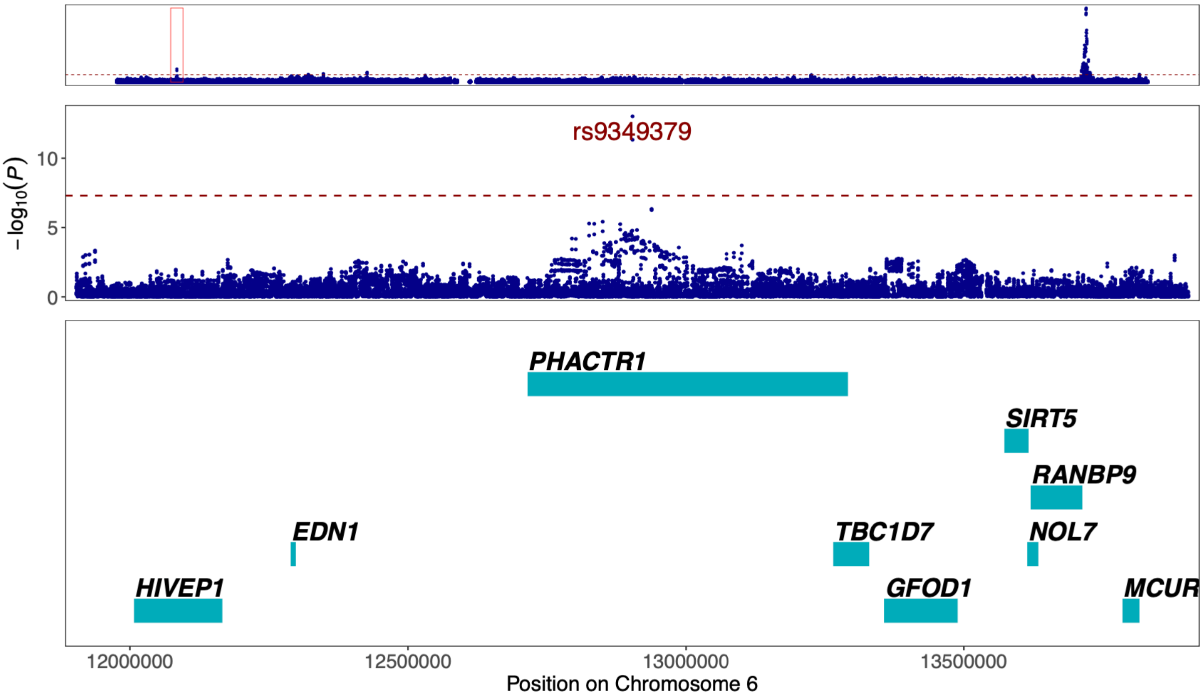


Figure 67. **Regional association plot of previously unreported AAA signal at 6p24.1 around the GWAS lead variant rs9349379.** The most likely causal gene explaining the association at the locus was predicted to be *PHACTR1* (*Phosphatase And Actin Regulator 1*). The top panel shows an overview of the genomic position, the middle and bottom panels depict individual SNVs and genes within the 2 MB locus window on chromosome 6. The middle Y-axis displays the strength of the association as -$\log_{10}(P)$ with a dashed red line marking the threshold for genome-wide significance ($P < 5 \times 10^{-8}$).

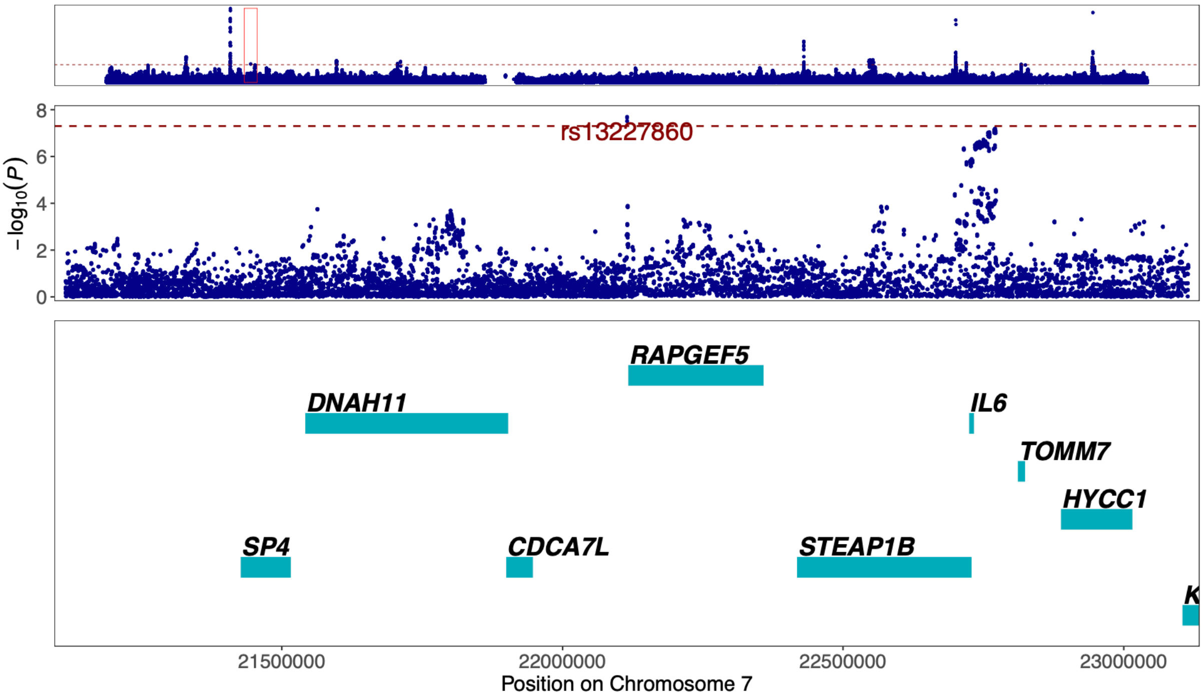


Figure 68. **Regional association plot of previously unreported CAD signal at 7p15.3 around the GWAS lead variant rs13227860.** The most likely causal gene explaining the association at the locus was predicted to be *RAPGEF5* (*Rap Guanine Nucleotide Exchange Factor 5*). The top panel shows an overview of the genomic position, the middle and bottom panels depict individual SNVs and genes within the 2 MB locus window on chromosome 7. The middle Y-axis displays the strength of the association as -$\log_{10}(P)$ with a dashed red line marking the threshold for genome-wide significance ($P < 5 \times 10^{-8}$).

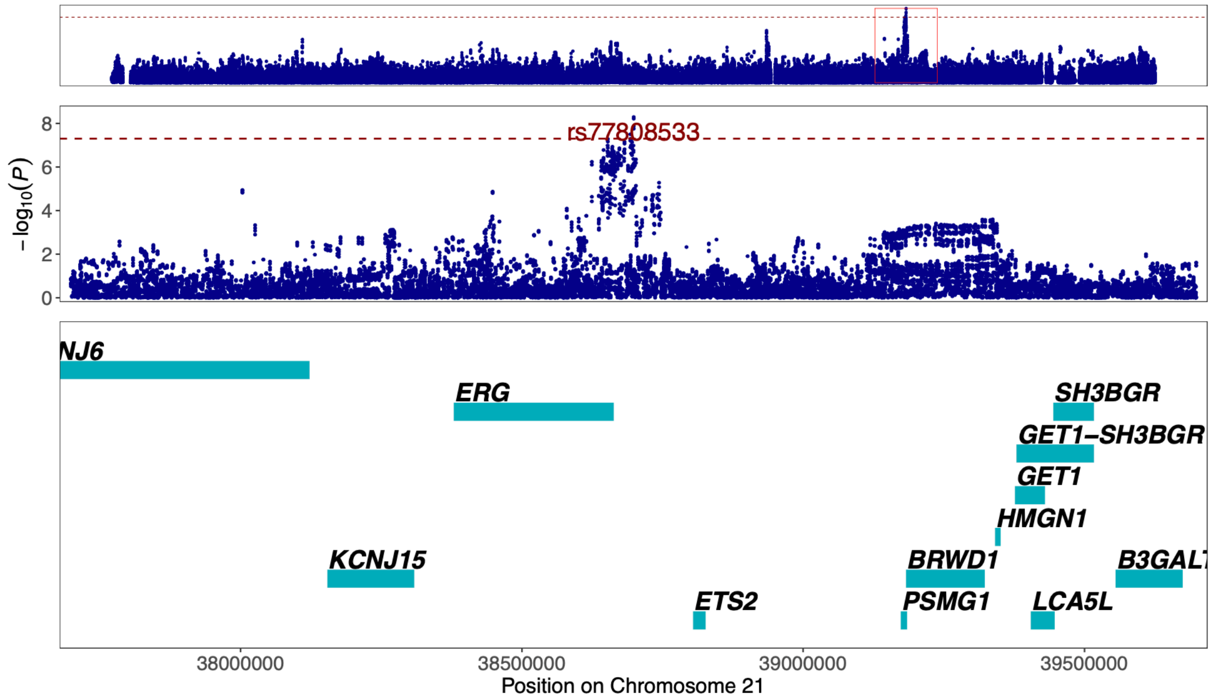


Figure 69. **Regional association plot of previously unreported TAA signal at 21q22.2 around the GWAS lead variant rs77808533.** The most likely causal gene explaining the association at the locus was predicted to be *ERG* (*ETS Transcription Factor ERG*). The top panel shows an overview of the genomic position, the middle and bottom panels depict individual SNVs and genes within the 2 MB locus window on chromosome 21. The middle Y-axis displays the strength of the association as -log$_{10}$(*P*) with a dashed red line marking the threshold for genome-wide significance ($P < 5 \times 10^{-8}$).

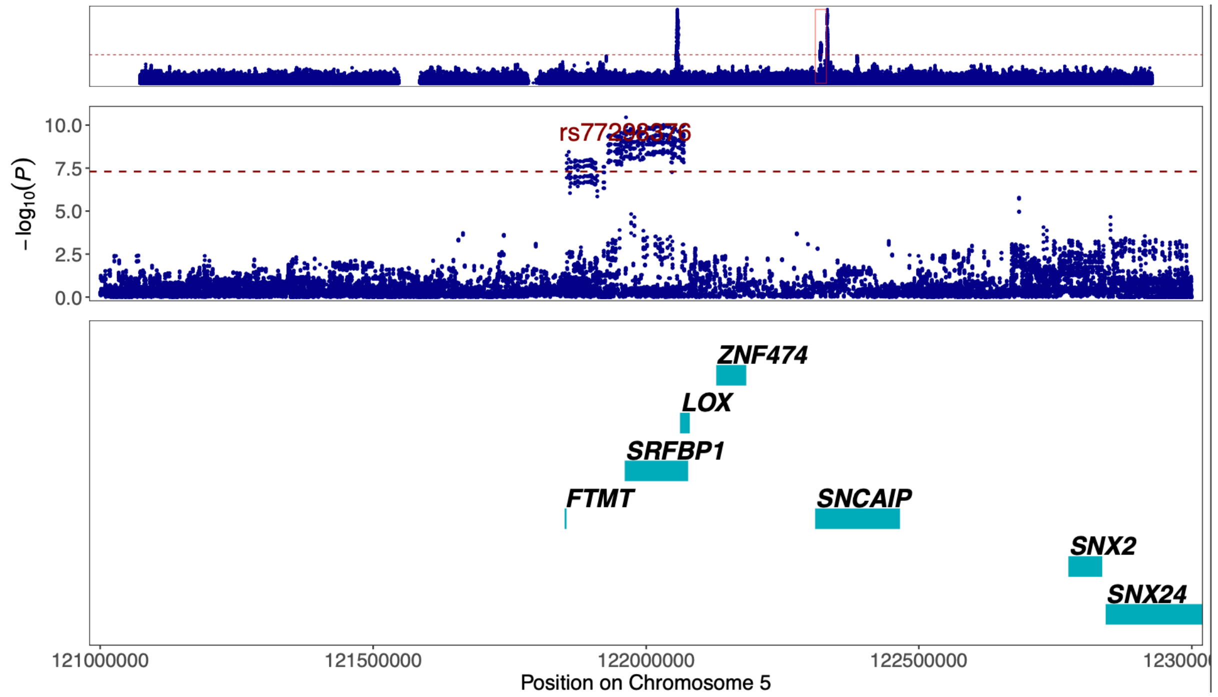


Figure 70. **Regional association plot of previously unreported TAA signal at 5q23.1 around the GWAS lead variant rs77298376.** The most likely causal gene explaining the association at the locus was predicted to be *SRFBP1* (*Serum Response Factor Binding Protein 1*). The top panel shows an overview of the genomic position, the middle and bottom panels depict individual SNVs and genes within the 2 MB locus window on chromosome 5. The middle Y-axis displays the strength of the association as -log$_{10}$(*P*) with a dashed red line marking the threshold for genome-wide significance ($P < 5 \times 10^{-8}$).

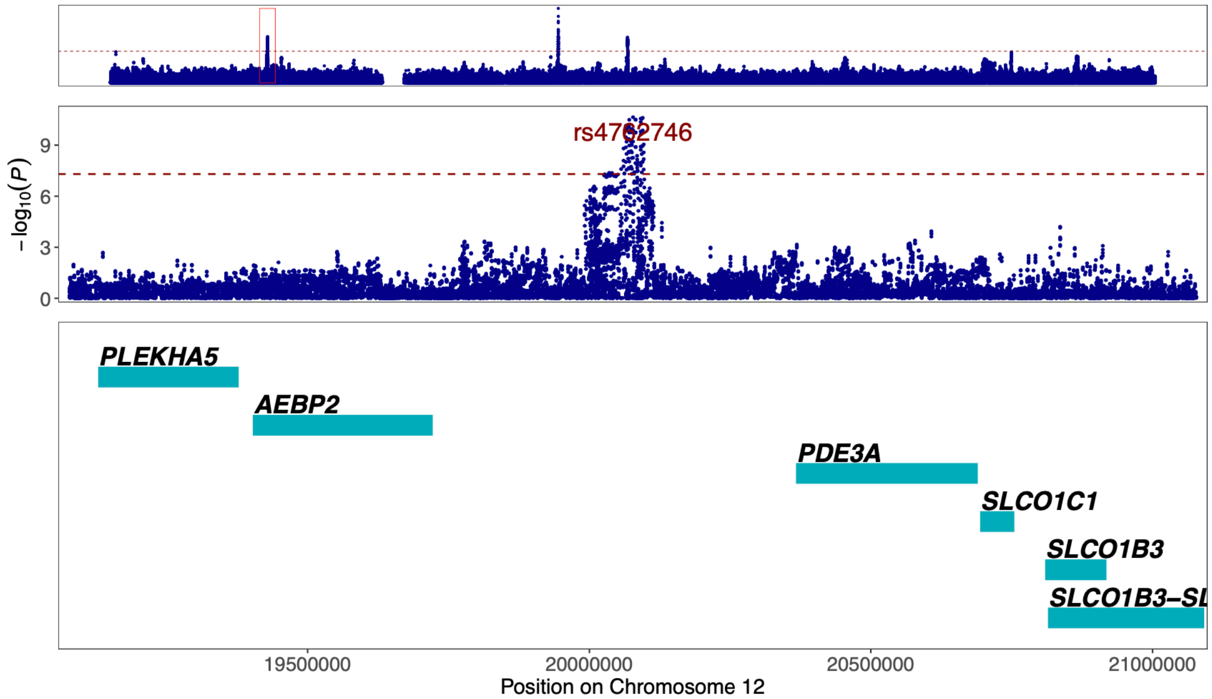


Figure 71. **Regional association plot of previously unreported TAA signal at 12p12.2 around the GWAS lead variant rs4762746.** The most likely causal gene explaining the association at the locus was predicted to be *PDE3A* (*Phosphodiesterase 3A*). The top panel shows an overview of the genomic position, the middle and bottom panels depict individual SNVs and genes within the 2 MB locus window on chromosome 12. The middle Y-axis displays the strength of the association as -$\log_{10}(P)$ with a dashed red line marking the threshold for genome-wide significance ($P < 5 \times 10^{-8}$).

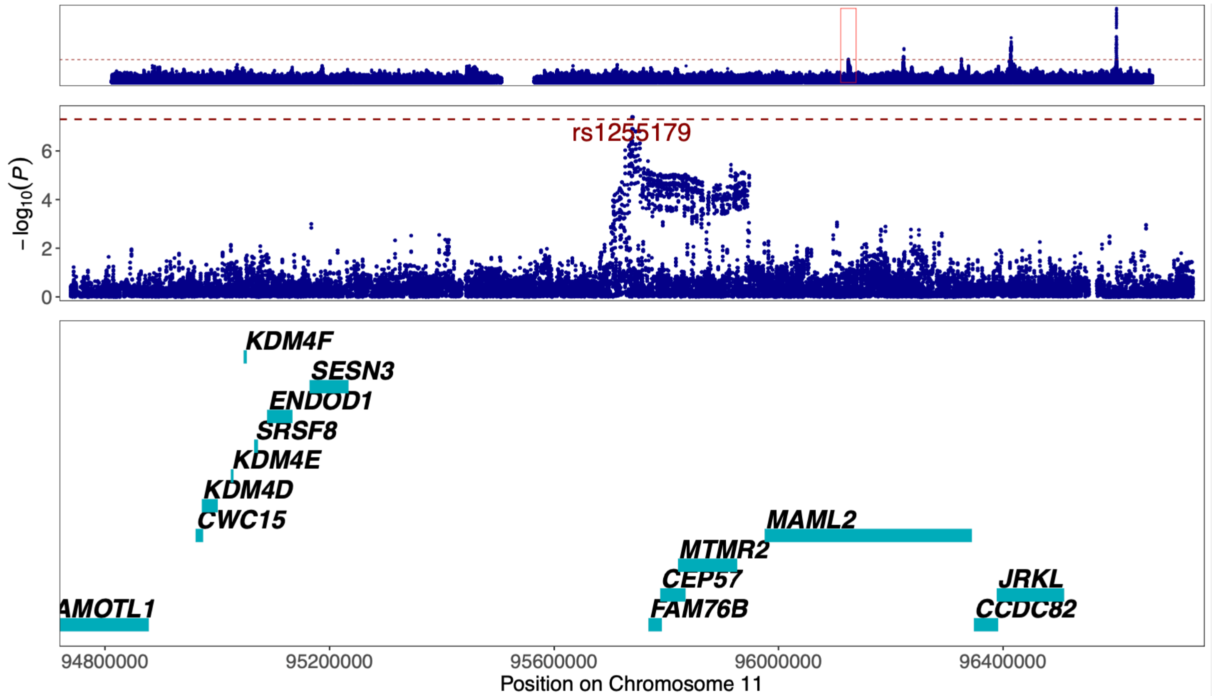


Figure 72. **Regional association plot of previously unreported AAA signal at 11q21 around the GWAS lead variant rs1255179.** The most likely causal gene explaining the association at the locus was predicted to be *CEP57* (*Centrosomal Protein 57*). The top panel shows an overview of the genomic position, the middle and bottom panels depict individual SNVs and genes within the 2 MB locus window on chromosome 11. The middle Y-axis displays the strength of the association as -$\log_{10}(P)$ with a dashed red line marking the threshold for genome-wide significance ($P < 5 \times 10^{-8}$).

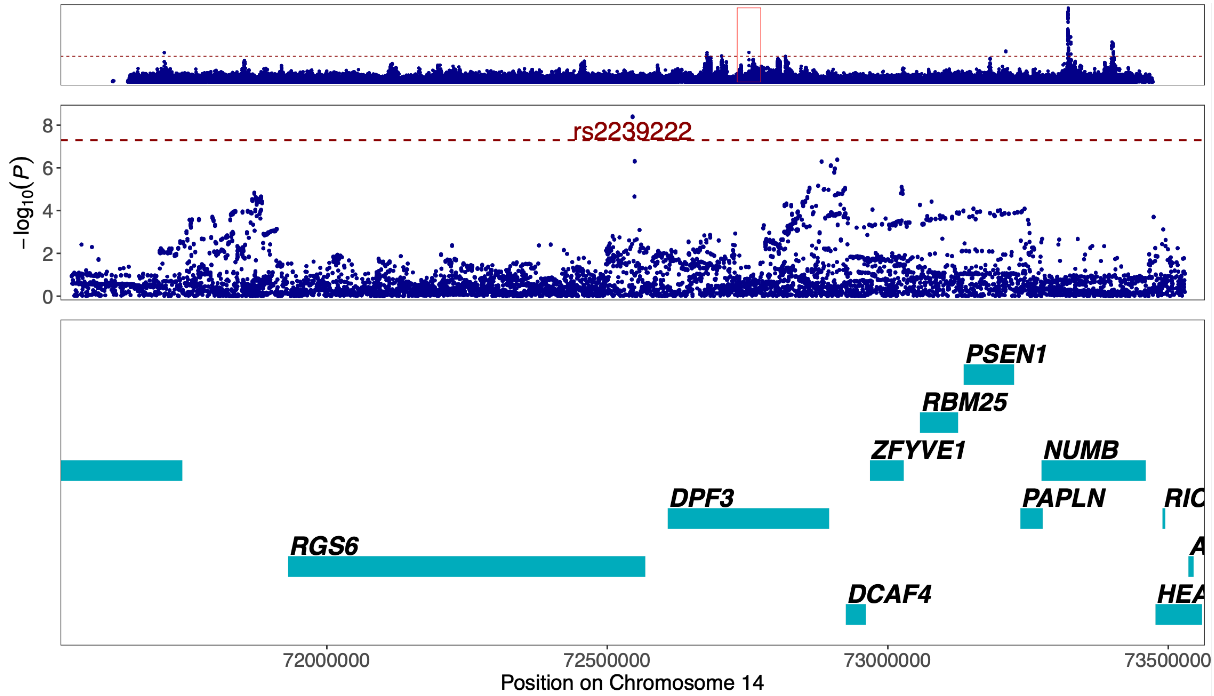


Figure 73. **Regional association plot of previously unreported CAD signal at 14q24.2 around the GWAS lead variant rs2239222.** The most likely causal gene explaining the association at the locus was predicted to be *DPF3* (*Double PHD Fingers 3)*. The top panel shows an overview of the genomic position, the middle and bottom panels depict individual SNVs and genes within the 2 MB locus window on chromosome 14. The middle Y-axis displays the strength of the association as -$\log_{10}(P)$ with a dashed red line marking the threshold for genome-wide significance ($P < 5 \times 10^{-8}$).
